# Effects of depression genetic risk and household socioeconomic status on emotional behavior and brain development in early adolescence

**DOI:** 10.1101/2025.05.16.25327790

**Authors:** Claire E. Campbell, W. James Gauderman, Megan M. Herting

**Author notes:** Corresponding Author: Correspondence to Megan M. Herting. University of Southern California, 1845 N. Soto Rm 225N, Los Angeles, California, 90089.

## Abstract

The World Health Organization (WHO) ranks depression as the number one non-fatal contributor to the global burden of disease. Previous work finds that early intervention prior to onset leads to preferred outcomes. To examine depression prior to onset, risks are assessed in relation to depressive prodromal behavior and brain biomarkers. Research shows environmental and genetic risks both separately influence depression onset, but little research has examined their interaction. Thus, we examined whether the socioeconomic predictor of family income-to-needs ratio (INR) may have independent and/or interactive effects with an individual’s depression polygenic risk score (D-PRS) on youth behavior and brain structure and function. We leveraged the U.S. based ABCD Study® - an existing longitudinal dataset; this work examined ∼8,000 subjects, 9-10 year-of-age at baseline to 11-12 year-of-age at 2-year follow-up. Historically, genetic analysis was conducted in European-like samples. Therefore, our work first assesses effects in youth with European-like genetic ancestry. Next, to improve generalizability, the same analyses were conducted in youths who are not European-like. Overall, D-PRS was associated with behavior and brain within the European-like youths, but less so in the not European-like sample. A potential moderating effect of INR on D-PRS was also seen for brain network connectivity, with further main effects of INR in youths from lower socioeconomic statuses. The findings from this project suggests behavior-brain biomarkers associated with socioeconomic status and depression risk vary by genetic ancestry, further highlighting the importance of precision-based medicine research in hopes of improving early detection and treatment of depression.

## **A.** Introduction

Half of all mental health disorders arise by age 14 (1), with most cases going untreated. In particular, Major Depressive Disorder (MDD) appearing before age 30, which is classified as early onset depression (2), is of increasing concern, with current rates around 16% of U.S. adolescents (3). During adolescence, emotional and cognitive control brain regions continue to develop, streamlining into efficient circuits, which leaves the brain vulnerable to environmental factors that could moderate an individual’s genetic predisposition to depression (4). Therefore, identifying brain-behavior biomarkers of depression risk, particularly prodromal markers emerging before disorder onset, is essential for effective intervention (5).

Two transdiagnostic prodromal behavioral markers of depression risk are decreased positive affect (6) and increased severity of depressive/withdrawn symptoms (7). In the U.S., MDD is diagnosed clinically when an individual endorses five or more symptoms for at least two weeks, complicating early identification of at-risk individuals before meeting diagnostic criteria (8). Focusing on prodromal markers allows for a dimensional assessment approach, emphasizing symptoms over formal diagnosis. Regarding brain biomarkers, previous studies have aimed to identify brain phenotypes related to depressive symptoms in healthy individuals and those already diagnosed. In doing so, depression symptomatology in adolescents (9) and young adults (10) has been found to inversely correlate with intra-network functional connectivity of the Salience Network (SN), where increased connectivity links to lower positive affect (9,10). Other studies have noted decreased functional connectivity between the SN and Frontoparietal Network (FPN) with the amygdala and hippocampus in depressed young adults (11). Cross-sectionally, elevated intra-network Default Mode Network (DMN) connectivity has also been observed in depressed adolescents (10), while decreased FPN connectivity was reported in adults with heightened depressive symptoms (12). Depression has also been associated with structural brain changes. Reduced cortical thickness cross-sectionally in adolescents (13) and surface area longitudinally in adults (14) are associated with increased depressive symptoms. The amygdala and hippocampus (15) also show structural and functional alterations in children and young adults with depression, including smaller hippocampal volumes in children displaying increased withdrawal symptoms (16). Yet, questions remain as to how genetic and environmental factors may exert unique and/or interactive effects to influence these brain-behavior biomarkers of depression risk during adolescent development.

The World Health Organization emphasizes that mental health is shaped partly by an individual’s environment, including socioeconomic factors (17). Systematic reviews show pervasive negative associations between common mental disorders and childhood socioeconomic status (SES) (18). Lower household income is linked to prodromal depression markers, including increased depressive symptoms and decreased positive affect in youth. Given ongoing brain maturation during childhood and adolescence, SES may influence depression risk by altering neurodevelopmental trajectories. Lower household SES has been related to decreases in intra-network connectivity of the SN in childhood (19), whereas relationships between SES and intra-network DMN connectivity during childhood have been mixed (21, 22). Lower household SES is also related to decreased surface area (20) and reduced amygdala (21) and hippocampal volumes (22). These findings underscore SES as a crucial factor when exploring brain biomarkers related to early onset depression.

Beyond the environment, genetic factors also play a significant role in risk for depression. Early onset depression is more influenced by genetic makeup compared to late onset (23). Assessing genetic risk through polygenic risk scores (PRS) can aid in developing personalized care pathways for youth at risk for early onset depression (24). However, few longitudinal studies have examined the effect of depression-related PRS on early-onset depression (24,25). Moreover, most studies investigating neurobiological markers of depression have relied on small cross-sectional samples, with limited attention to longitudinal changes or the racial and ethnic composition of cohorts. Furthermore, little work has examined the combined effects of genetics and SES on longitudinal changes in brain function, structure, and behavior related to depression in diverse youth cohorts.

The current research aims to fill these gaps by investigating the independent and interactive effects of household socioeconomic status and depression genetic risk on longitudinal changes in prodromal behavior markers of depression and emotional brain networks using data from the nationwide, diverse Adolescent Brain Cognitive Development (ABCD) Study (26). Ultimately, studying how genetics and SES jointly influence brain-behavior development in diverse youth cohorts can help clarify their independent and interactive effects on depression risk, enabling earlier detection and more targeted interventions.

## **B.** Methods

This research utilized genetic, behavioral, neuroimaging, and demographic data from the ABCD Study® - a longitudinal single cohort study (26). Harmonized data was collected from 21 U.S. sites, allowing for a diverse cohort of 11,873 children (48% female, baseline race/ethnicity characteristics: 50% non-Hispanic white, 22% Hispanic, 15% non-Hispanic Black, 2% Asian, and 11% mixed/other) (26). The ABCD Study is intended to represent a population-based, non-clinical sample; based on study design, we have longitudinal data collected at differing timepoints (**Figure 1**).

**Figure 1:**
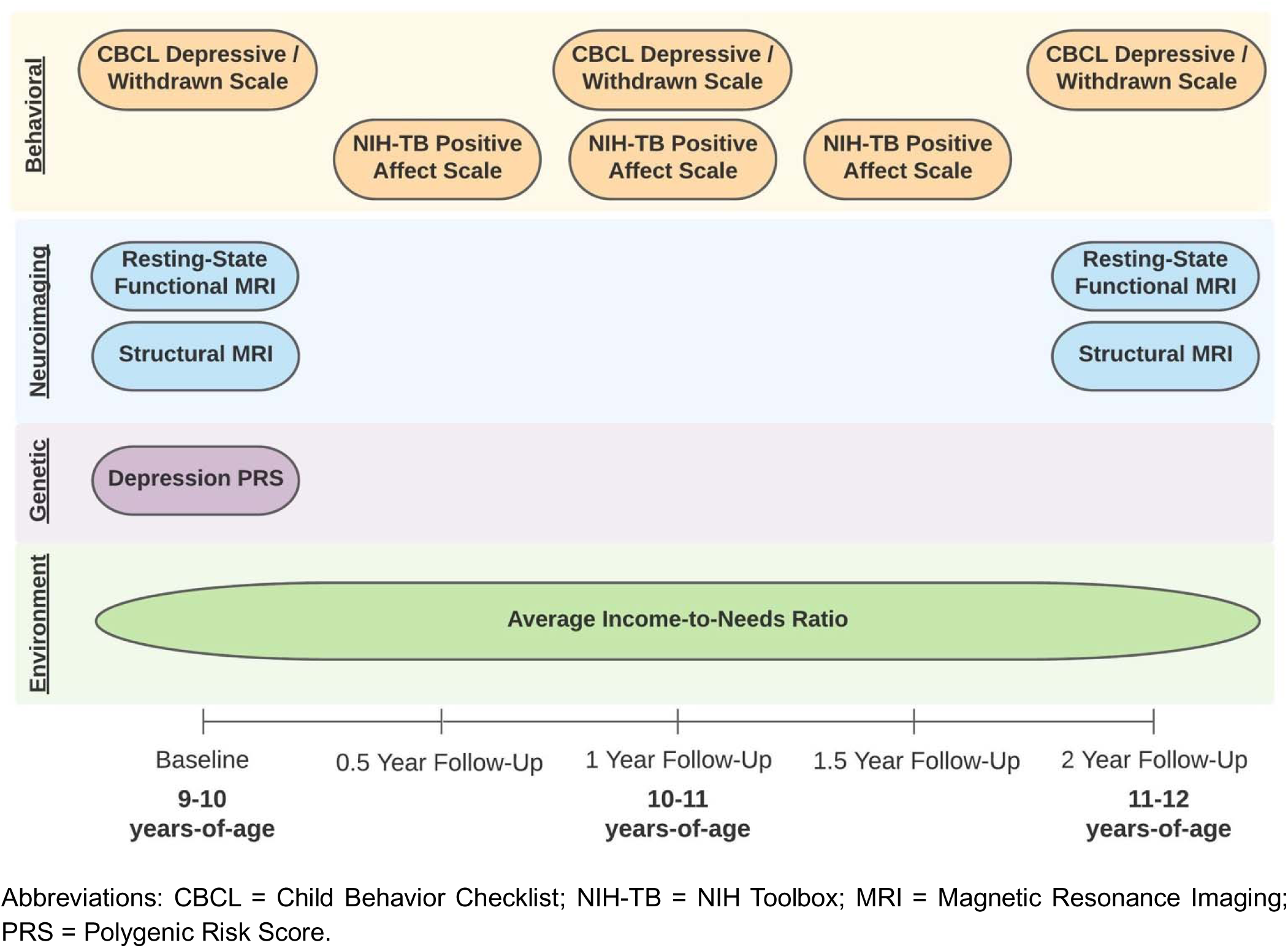
Collection timeline of data for emotional behavior and brain analyses. Abbreviations: CBCL = Child Behavior Checklist; NIH-TB = NIH Toolbox; MRI = Magnetic Resonance Imaging; PRS = Polygenic Risk Score.

### B.1 Exclusionary Criteria

#### B.1.1 General Exclusionary Criteria for the Complete Sample

From the larger ABCD cohort, additional exclusions were applied (**Supplemental Section A.1, Supplemental Figure 1**). Participants were required to have genetic data and at least one sociodemographic or behavioral measure. To satisfy model assumptions regarding independent observations, one subject per family was randomly selected. For neuroimaging, those without handedness scores or with poor MRI data were excluded (**Supplemental Section A.2**). Since most genome wide association studies (GWAS), including this study, use European-like discovery samples (27), analyses was first conducted on subjects with ≥80% European-like ancestry (28) based on the 1000 Genome reference panel phase 3 release (29), followed by tests in those who are not European-like (<80% European-like ancestry) to assess generalizability. Due to collinearity between the depression polygenic risk score and genetic principal components (PCs), not European-like individuals were split into two groups using the first population stratification PC (see **Section B.2.3, Supplemental Figures 2–4**).

### B.2 Data Collection and Preparation

#### B.2.1 Behavioral Data

Two behavioral questionnaires were utilized for the proposed research study: the Child Behavior Checklist (CBCL) (30) and the National Institute of Health (NIH) Toolbox Emotion Battery – Positive Affect (31). The CBCL is a 113-item parent-completed questionnaire (e.g., “Show little interest in things around him/her”) assessing the child’s behavior over six months, rated on a 3-point scale: not true (0 points), somewhat or sometimes true (1 point), very often or always true (2 points). Syndrome scales are calculated continuously, with this study focusing on the withdrawn/depressed subscale (8 items, 0–16 points) with higher scores indicating greater severity. The NIH Toolbox Emotion Battery – Positive Affect questionnaire, completed by the child, includes 9 items (e.g., “I felt at ease”) rated on a 3-point scale: not true (1 point), somewhat true (2 points), very true (3 points), with total scores ranging from 9 to 27 points and higher scores indicating greater positive affect.

#### B.2.2 Neuroimaging Data

Anatomical T1-weighted and functional T2*-weighted MRI scans were collected on 3-Tesla scanners (Siemens Prisma, General Electric (GE) 750, and Philips). The ABCD Study® ensured multi-site harmonization in image collection, preprocessing, and quality control (**Supplemental Sections A.2-A.3**) (32). Resting-state fMRI analyses focused on *a priori* intra- and inter-network connectivity within the frontoparietal (FPN), default mode (DMN), and salience (SN) networks (**Supplemental Figure 5, Supplemental Table 1**), plus connectivity between these networks and bilateral hippocampus and amygdala, totaling 18 outcomes. Structural MRI analyses included cortical thickness and surface area of 22 Destrieux atlas regions per hemisphere (88 measurements) and bilateral amygdala and hippocampus volumes (4 measurements).

#### B.2.3 Depression Polygenic Risk Score (D-PRS)

Using ABCD Study genetic data (**Supplemental Section A.4.1**), depression polygenic risk scores (D-PRS) were generated based on 2018 Psychiatric Genomics Consortium GWAS summary statistics (33), with higher scores indicating greater polygenic risk. For details on the D-PRS and population PCs generated in our sample see **Supplemental Section A.4.2**. Since late- and early-onset depression may have different genetic predispositions (23), this D-PRS was chosen for its replication in pediatric samples (24), given the study’s focus on early-onset depression. Due to collinearity between D-PRS and PCs in the not European-like group, the first PC split this group into two, resulting in three analysis samples: European-like, not European-like Group 1, and not European-like Group 2. D-PRS values were mean-scaled within groups to adjust for population stratification. Scree plot analysis confirmed 10 PCs sufficiently captured ancestry variance, so all models included 10 PCs as covariates.

#### B.2.4 Incomes-to-Needs Ratio (INR)

Household size and income data from the ABCD Study were used to compute INR as previously published (20) (see **Supplemental Section A.5**). Given INR showed moderate to good reliability across three timepoints (ICC = [0.74, 0.81]), we used the mean of available values, with higher scores indicating greater SES advantage.

#### B.2.5 Covariates and Confounders

Models adjusted for time-varying covariates (study-site and age) and time-invariant covariates obtained from the baseline study visit (sex-at-birth (3), caregiver-identified race/ethnicity (3), parental education (34), and the first 10 genetic PCs). In the neuroimaging analyses, study-site was replaced with MRI scanner serial number; these variables are highly correlated, but the MRI scanner serial number offers a more refined grouping. Handedness was also included as a time-invariant covariate in the neuroimaging analyses, resting-state fMRI models included MRI motion estimates, while structural MRI models included intracranial volume - estimated by Freesurfer - to account for overall brain size. Here, we conceptualize it as a social construct that classifies individuals into politically driven groups to establish social hierarchies, while ethnicity is another social construct that categorizes diverse populations to inform personal and group identities (35). The combined variable of race and ethnicity in ABCD is used to include individuals who personally identify as Hispanic or Latinx when asked about their race (see **Supplemental Section A.6**). Descriptive demographics by collection date and outcome are available in **Supplemental Tables 2-11**.

### B.3 Data Analysis

Initial descriptive and exploratory analyses checked for errors, outliers, and assessed distributions and correlations (**Supplemental Figure 6-9**). Models were evaluated for assumption violations. D-PRS values were mean-scaled for interpretability, while INR values remained untransformed to maintain the interpretability and generalizability across sample groups. A multilevel modeling approach leveraged the longitudinal dataset and handled missing data by using all available observations. An initial two-level model estimated the intercept (initial value at ages 9–10) and slope (age-related change) for associations with D-PRS, INR, and their interaction. Given the complexity of our initial models, higher-order terms were removed stepwise if nonsignificant (**Supplemental Section A.7**), prioritizing model parsimony (see **Supplemental Table 12**). The withdrawn/depressed score was right-skewed, making a standard linear model inappropriate due to propensity for artificial inflation of coefficient significance (36). Instead, this measure was treated as a count metric and a negative binomial model was used to account for overdispersion (37). The modeling equations were the same, but the outcome, *Y*_*ij*_ , was log adjusted, and the error term, *ε*_*ij*_, was removed. Positive affect was not heavily skewed, so our outlined linear mixed-effects model was applied. Cortical region modeling used a linear mixed-effects model given the even distribution. Preliminary analyses showed similar hemispheric trajectories, so both hemisphere measurements were nested within subjects to account for within-individual similarity and maximize available data. Since D-PRS did not predict depression behavior in not European-like Group 2 (see below and **Figure 2A, Supplemental Table 13**), D-PRS and INR were modeled only in European-like and not European-like Group 1, while only INR effects were examined in not European-like Group 2.

**Figure 2:**
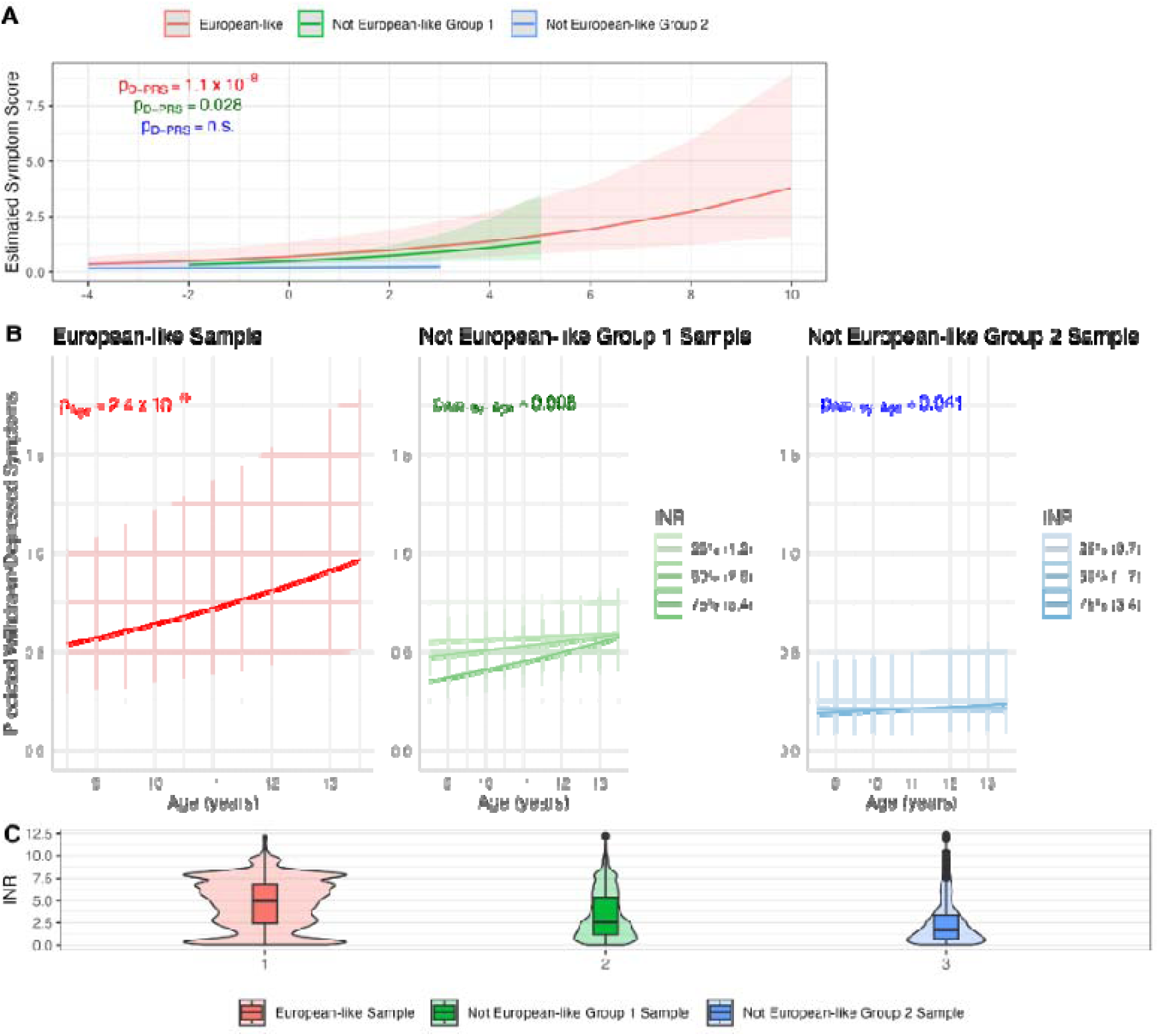
Significant effect of D-PRS, Age, and INR on Withdrawn/Depressed Symptoms. A) Effect of the depression polygenic risk score (D-PRS) on the withdrawn/depressed symptoms in each ancestry group; B) Effect of age in the European-like sample, plus the effect of age modified by income-to-needs ratio (INR) in the not European-like Group 1 and Group 2 samples on the CBCL withdrawn/depressed symptoms, displaying age effects at the 25th, 50th, and 75th percentiles within each sample, with all other variables held constant; C) violin plots showing the distribution of income-to-needs (INR) across each group with width relative to the sample size. Note, CBCL withdrawn/depression symptom scale can range from 0–16 points. Abbreviations: p_D-PRS_ = p-value for effect of D-PRS; p_Age_ = p-value for effect of age; p_INR-by-Age_ = p-value for effect of INR-by-age interaction.

## **C.** Results

In the European-like sample, adolescents were mostly caregiver-identified as non-Hispanic white. For the not European-like Group 1, adolescents were mostly caregiver-identified as Hispanic, while adolescents included in the not European-like Group 2 were mostly caregiver-identified as non-Hispanic Black (**Supplemental Tables 2-11**). At the first timepoint, poverty rates (<1 INR) were 16% for European-like, 20-21% for not European-like Group 1, and 32-34% for not European-like Group 2. After mean-scaling D-PRS, no mean differences appeared between groups, though the European-like sample showed the widest range of polygenetic risk scores (**Supplemental Figure 10**).

### C.1 Behavioral Analyses: Withdrawn/Depressed Symptoms

#### C.1.1 Depression Polygenic Risk Score Effects

In the European-like sample, there was a main effect of D-PRS (IRR=1.184, p=1.1x10⁻ ) (**Figure 2A**, **Supplemental Table 13**). A small main effect was also found in the not European-like Group 1 (IRR=1.223, p=0.028), but this effect was deprecated compared to the European-like sample. No significant D-PRS effects were found for the not European-like Group 2 (**Supplemental Table 13**). The lack of D-PRS by age interaction across groups suggests a consistent genetic effect from ages 9-12 (see **Supplemental Section B.1**).

#### C.1.2 Age Effects and Moderating Effect of Income-to-Needs Ratio

In the European-like sample, age significantly affected withdrawn/depressed symptoms, but INR did not (**Figure 2B; Supplemental Table 13**). In European-like Group 1 (IRR=1.02, p=0.008) and Group 2 (IRR=1.021, p=0.041), a significant interaction between INR and age was observed, with stronger age effects at higher INR; albeit this effect was minor in Group 2. Mean INR differed significantly across groups: 4.6 in the European-like, 3.4 in not European-like Group 1, and 2.4 in not European-like Group 2 (**Figure 2C; Supplemental Table 13**). The higher mean INR in the European-like group may obscure a potential INR-age interaction evident at lower INR values in not European-like groups.

### C.2 Behavioral Analyses: Positive Affect

#### C.2.1 Depression Polygenic Risk Score Effects

To assess the generalizability of D-PRS to other depression-related indicators, we examined positive affect. D-PRS was inversely related to positive affect in individuals with European-like ancestry, while in not European-like samples, the negative trend was nonsignificant (**Figure 3; Supplemental Table 14**). This likely reflects the limited generalizability of our D-PRS score, which was developed in a European-like sample (see details in **Supplemental Section B.2**).

**Figure 3:**
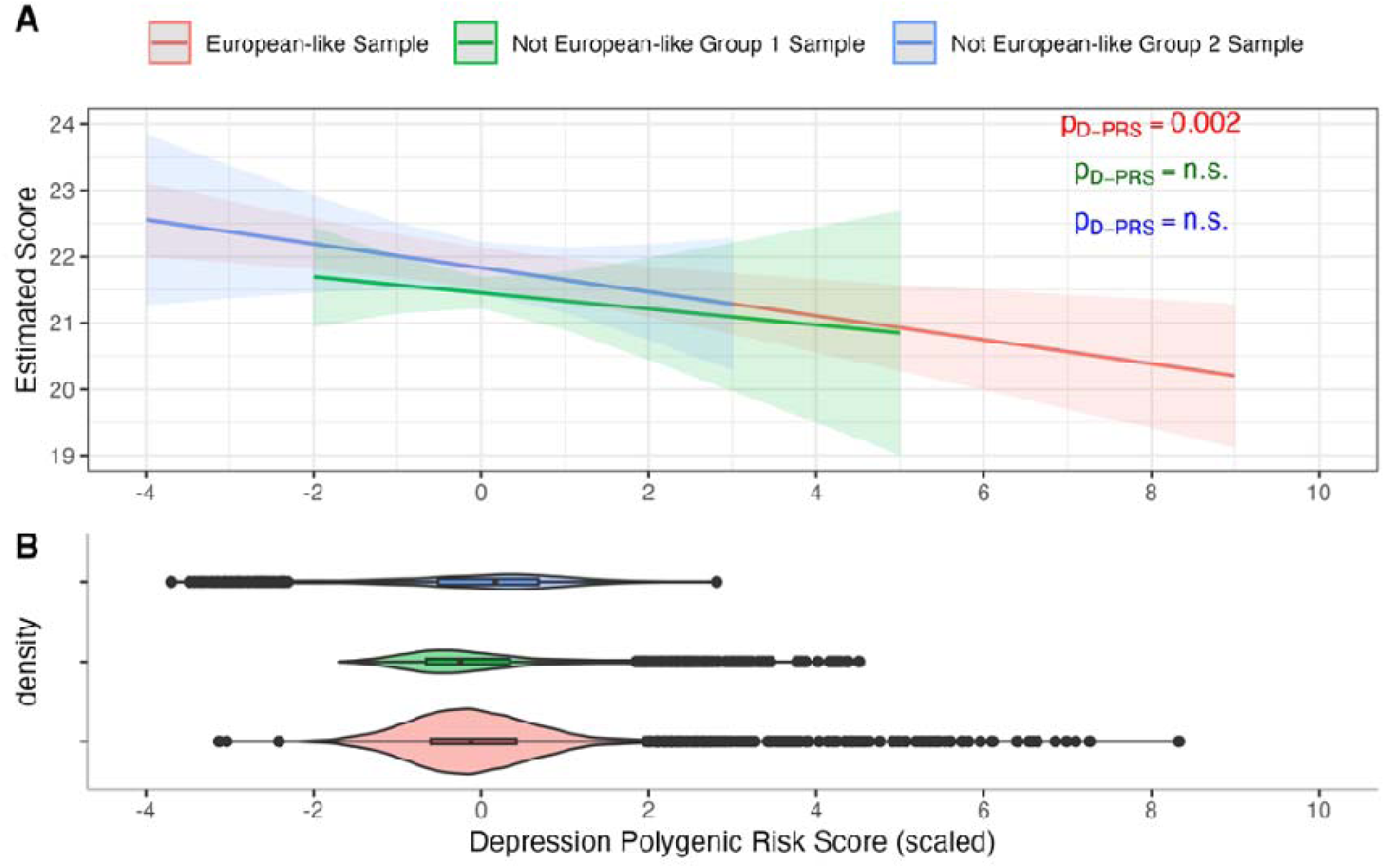
Effect of D-PRS on Positive Affect. A) Effect of the depression polygenic risk score (D-PRS) on the positive affect questionnaire in each group; B) Distribution of D-PRS by analysis group. Note, Positive Affect scale can range from 9–27 points. Abbreviations: p_D-PRS_ = p-value for effect of D-PRS; n.s. = not significant.

##### C.1.2.2 Income-to-Needs Ratio Effects

An INR effect was only observed in adolescents within the not European-like Group 1 (*β*=0.1, p=0.001), with higher INR leading to higher positive affect (**Supplemental Figure 11**; **Supplemental Table 14**). For a 1-unit increase in INR there was a 0.1-point increase in self-reported positive affect, which is minimal given the range of the potential points (9-27 points). INR was not significant in the European-like (*β*=0.017, p=0.238) and not European-like Group 2 (*β*=0, p=0.997) (**Supplemental Figure 11**; **Supplemental Table 14**).

### C.3 Neuroimaging Analyses: Resting-State Functional MRI

#### C.3.1 Income-to-Needs Ratios and Depression Polygenic Risk Scores Interactive Effects

In the European-like sample, trending INR and D-PRS interactions were observed for intra-network connectivity of the DMN (p=0.006) and FPN (p=0.005) after FDR correction, independent of age (**Figure 4B, Supplemental Table 16**). This interaction indicated a positive D-PRS effect on DMN and FPN connectivity in adolescents from higher-INR families, with a small negative or no effect in lower-INR families. Given the trending nature of these interactions, we also removed this interaction and examined the main effects, though no main effects of INR or D-PRS passed FDR correction (**Supplemental Table 15**). In both not European-like groups, no significant INR or D-PRS interactions or main effects were observed after FDR correction (**Supplemental Table 15**).

**Figure 4:**
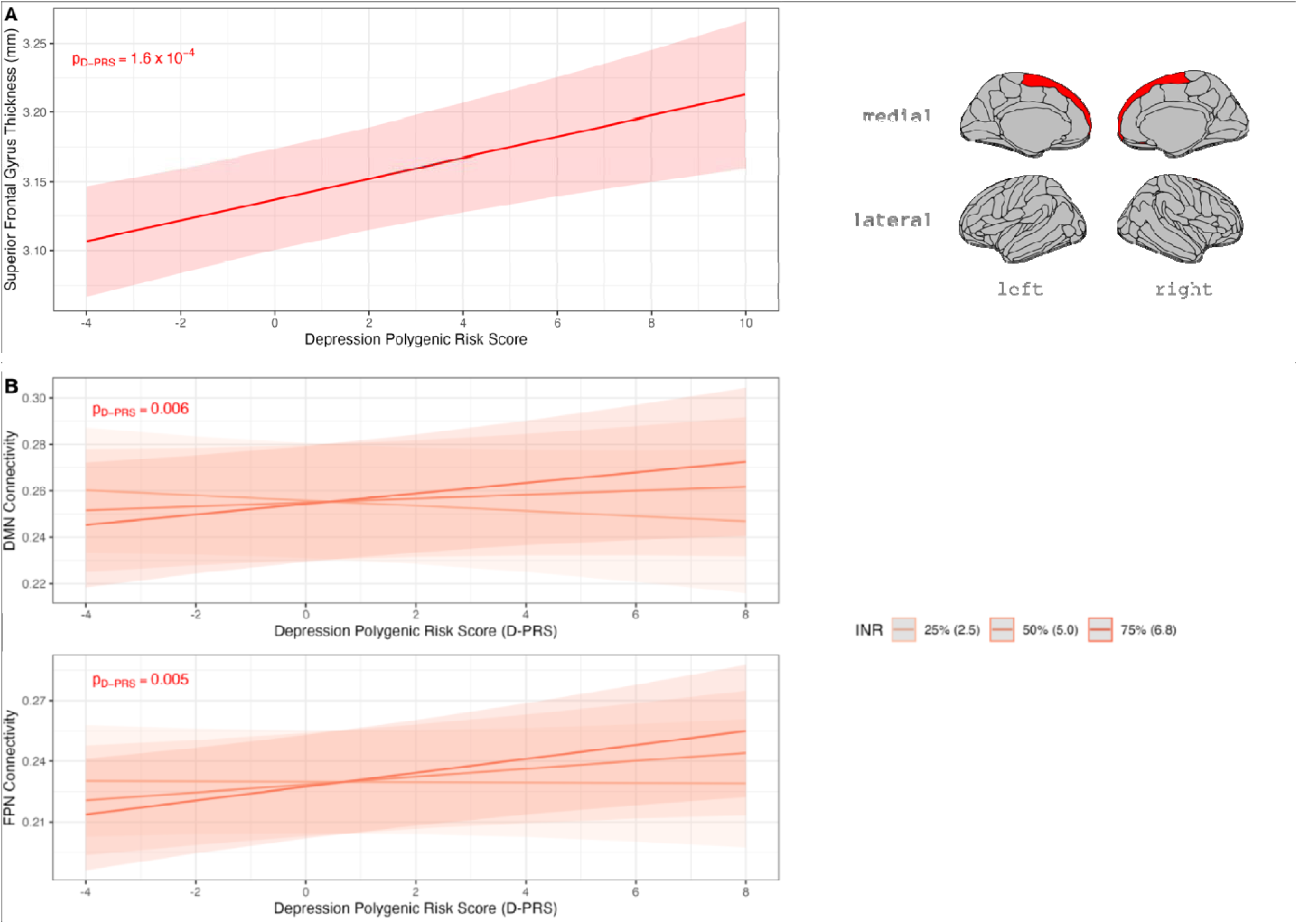
INR and D-PRS findings for the European-like group for intra-network connectivity and structural MRI. A) Effect of depression polygenic risk score (D-PRS) on the thickness of the superior frontal gyrus in the European-like sample (left) with corresponding visual of the superior frontal gyrus location (right); age centered at 10 years; other covariates held constant (caregiver identified race/ethnicity, income-to-needs ratio, MRI scanner serial number, sex-at-birth, highest parents’ education, handedness, and the first 10 genetic PCs). B) Trending interaction between INR and D-PRS of the intra-network connectivity of the Default mode network (DMN) (top) and frontoparietal network (FPN) (bottom) in adolescents in the European-like group; the INR categories are the 25th, 50th, and 75th percentile values of the distribution of INR in the European-like sample; this corresponds to an INR raw value of 2.5, 5.0, and 6.8, respectively; age centered at 10 years; other covariates held constant (caregiver identified race/ethnicity, MRI motion, MRI scanner serial number, sex-at-birth, highest parents’ education, handedness, first 10 genetic PCs). Abbreviations: mm = millimeters; INR = income-to-needs ratio; D-PRS = depression polygenic risk score; p_D-PRS_ = p-value for effect of D-PRS; MRI = magnetic resonance imaging.

### C.4 Neuroimaging Analyses: Structural MRI

#### C.4.1 Depression Polygenic Risk Scores Effects

For the European-like sample, there was a significant effect of D-PRS on the thickness of the superior frontal gyrus (β=0.008, p=1.6x10^-4^; **Supplemental Table 17**) - which is typically associated with both the FPN and DMN (**Supplemental Figure 5**, **Supplemental Table 1**). Specifically, higher D-PRS values were associated with greater thickness regardless of age of participants (**Figure 4A**).

#### C.4.2 Income-to-Needs Ratio Effects

In the European-like sample, a trending effect of INR on the middle temporal gyrus (β=3.381, p=0.004) showed that higher INR was linked to greater surface area in this DMN-associated region (**Supplemental Figure 12**; **Supplemental Table 17**). In not European-like Group 1, no significant or trending effects of D-PRS and INR were observed. In not European-like Group 2, significant INR-by-age interactions were found in several regions, with thickness changes in the fronto-marginal gyrus and sulcus (p=0.006), middle frontal gyrus (p=2.2x10^⁻^), and superior frontal gyrus (p=0.002) (**Figure 5**). Lower INR amplified age-related decreases in thickness between ages 9 and 13 years. Main INR effects showed significant surface area increases in the superior frontal sulcus (β=11.865, p=0.001) and inferior part of the precentral sulcus (β=7.461, p=0.001) in DMN- and FPN-related regions (**Supplemental Figure 13; Supplemental Table 17**). Positive trends were also noted in surface areas of the DMN and FPN-associated fronto-marginal gyrus and sulcus (β=3.781, p=0.016) and middle frontal gyrus (β=12.709, p=0.011), and DMN-associated precuneus (β=7.196, p=0.013) (**Supplemental Figure 14; Supplemental Table 17**), with no significant INR effects on amygdala and hippocampus volumes (p_FDR_>0.05; **Supplemental Table 18**).

**Figure 5:**
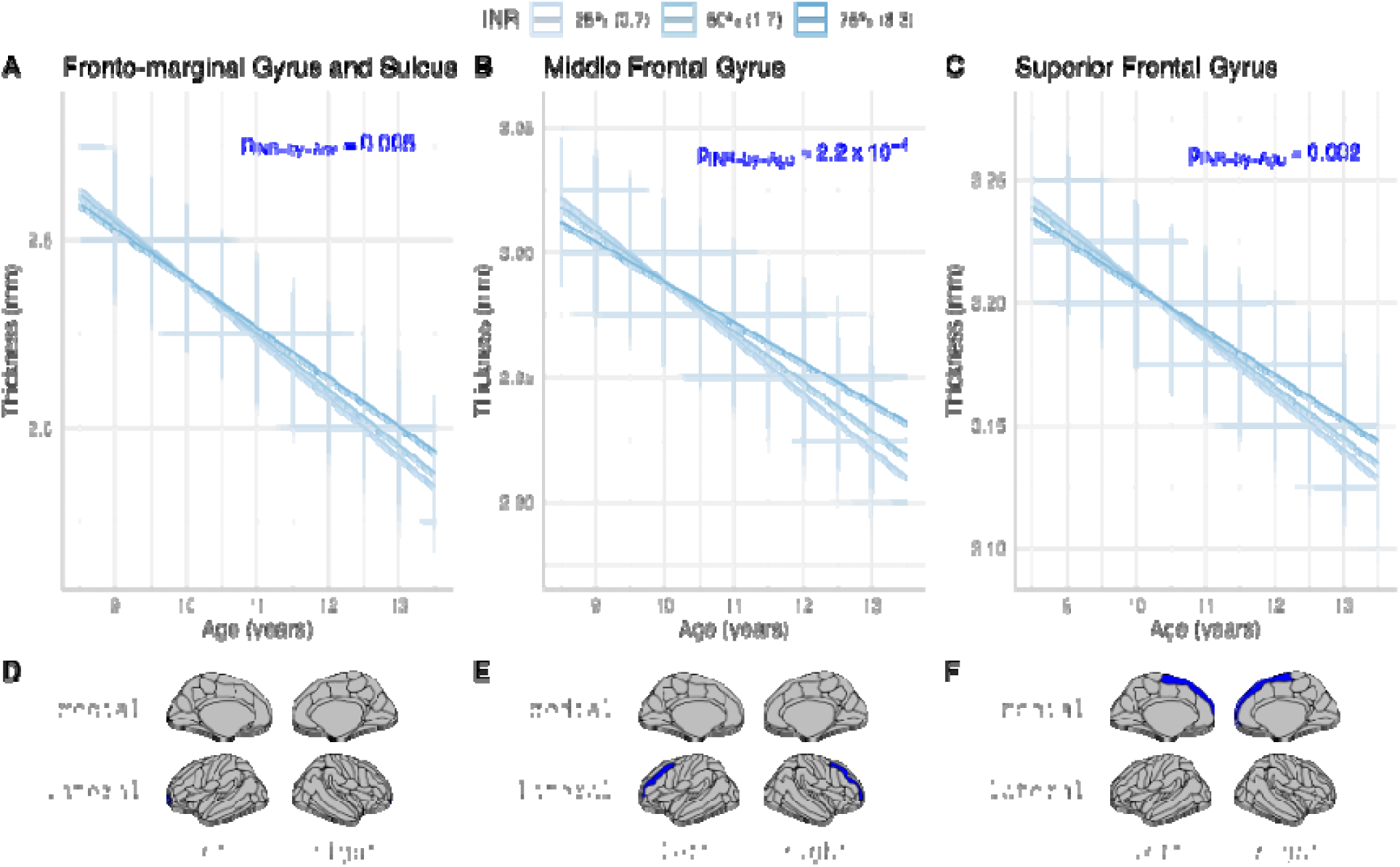
Significant effect of Age and INR on numerous sMRI metrics in the European-like Group 2 Sample not. A-C) Interactive effect of income-to-needs ratio (INR) and age on the thickness of the above structural MRI metrics in the not European-like Group 2 sample with D-F) corresponding visual of the brain region location below each graph. The INR categories are the 25th, 50th, and 75th percentile values of the distribution of INR in the not European-like Group 2 sample; this corresponds to an INR raw value of 0.7, 1.7, and 3.3, respectively. Covariates held constant (caregiver identified race/ethnicity, MRI scanner serial number, sex-at-birth, highest parents’ education, handedness). Abbreviations: p_INR-by-Age_ = p-value for interaction effect of INR and age; sMRI = structural magnetic resonance imaging.

## Discussion

This study investigated the independent and interactive effects of socioeconomic status (as measured by income-to-needs ratio; INR) and depression polygenic risk scores (D-PRS) on the behavioral and brain development of adolescents, using data from the ABCD Study. By examining these relationships across individuals with European-like and not European-like ancestry, we aimed to uncover how genetic predisposition to depression and environmental factors such as socioeconomic status may shape adolescent behavior and brain biomarkers associated with depression risk. Our study highlights distinct influences of D-PRS and INR on adolescent behavior and brain development, with genetic risk effects more prominent in European-like adolescents and socioeconomic effects in not European-like groups. These findings underscore the involvement of genetic and environmental risk factors in shaping depression-related brain biomarkers. As adolescent depression rates rise, understanding these dynamics may be crucial for developing effective, equitable interventions that address socioeconomic and racial disparities, ensuring mental health resources and policies are inclusive and applicable across diverse populations.

The D-PRS utilized in the current study was developed on an adult-based population (33) and previously replicated in children (33). Here, using the landmark ABCD Study cohort, we replicate this established D-PRS relates to greater withdrawn/depressed and positive affect behaviors in European-like ancestry youths between the ages of 9-13 years-old. Moreover, our findings indicate that this D-PRS, generated from European-like ancestry samples, shows stronger associations with depression-linked brain and behavioral markers in European-like individuals compared to those with not European-like ancestry. This lack of generalizability of D-PRS to not European-like groups was evident both in behavioral and neuroimaging results, with small effects observed in one not European-like sample but no significant effects in the other. Importantly, trend-level D-PRS associations with brain function were observed within the European-like group only. We found that higher D-PRS values correlated with increased intra-network connectivity within the DMN and FPN at higher INR levels, but this effect was diminished or absent at lower INR levels. These results suggest that while D-PRS may serve as a useful marker of depression risk, its relationship with functional brain connectivity is modified by socioeconomic context. Previous research has shown increased DMN connectivity patterns in adolescents diagnosed with MDD (10); however, many of these studies do not include socioeconomic measures, highlighting a potential bias in existing depression biomarkers towards higher-SES individuals. Our findings therefore underscore the importance of including socioeconomic status as a variable in neuroimaging research to enhance the generalizability and equity of mental health biomarkers.

For structural brain metrics, we observed associations between D-PRS and cortical thickness of the superior frontal gyrus in the European-like sample. Increased thickness in this region was associated with higher D-PRS values, consistent with findings in adult MDD studies where greater superior frontal gyrus thickness was observed in patients with MDD (38). However, in prior studies of adolescents with MDD, superior frontal thickness differences were not found (39), possibly indicating that D-PRS-associated structural changes may be more relevant as biomarkers of early-onset depression. Since past research has often used samples with higher SES backgrounds and lacked detailed socioeconomic data, findings in structural brain differences related to depression risk may also be incomplete or not broadly applicable. Considering this, the positive associations between D-PRS and superior frontal gyrus thickness we observed may reflect a specific vulnerability in European-like adolescents from higher SES backgrounds. More research is needed to assess whether this relationship persists or evolves with age and to determine if it can predict depression onset.

In contrast to the D-PRS findings, INR effects were primarily observed in not European-like individuals. Notably, we found age-by-INR interactions within the prefrontal cortex, indicating that low-SES adolescents in the not European-like samples experienced amplified effects of age on cortical metrics. This aligns with studies showing that lower household income is linked to greater negative age effects on brain development (40). The presence of more substantial INR-related effects in not European-like individuals could be reflective of broader systemic inequities that lead to SES disparities between ancestry groups, impacting neurodevelopmental outcomes. These structural changes emphasize the importance of studying socioeconomic factors independently and alongside genetic risk, as both SES and age contribute to significant developmental variations in brain structure among minoritized groups.

Our study faced several methodological challenges, including the ABCD Study’s higher average SES and uneven INR distribution across ancestry groups, which may have limited our ability to detect INR effects in lower SES European-like or higher SES not European-like participants. Additionally, low INR values among not European-like Group 1 participants likely contributed to the null D-PRS and INR associations in this group. Selection bias and retention issues, particularly among not European-like groups, further reduced sample overlap across study visits (**Supplemental Tables 19-20**), affecting generalizability. Despite these limitations, our study had many strengths. Depression rates in the ABCD Study are low at these time points (**Supplemental Table 21**), aligning with research showing peak early-onset depression diagnosis occurs between ages 20 and 30 (2). Since earlier onset is linked to greater disease burden, focusing on dimensional measures of depressive symptoms—such as withdrawn/depressed behaviors and positive affect—allows for a more nuanced assessment of early risk factors. In addition, the use of longitudinal data and diverse samples strengthens our findings further. Our findings highlight that recruiting individuals from diverse backgrounds and incorporating socioeconomic variables as covariates will help refine D-PRS applications, prevent biases rooted in higher-SES European-like samples, and support the development of inclusive mental health interventions and equitable policies.

## Data Availability

All data produced are available online at https://nda.nih.gov/abcd

## Acknowledgements

This work would not be possible without all the Adolescent Brain Cognitive Development^SM^ (ABCD) Study (https://abcdstudy.org) participants and their families; a huge thank you to all who participated, organized, and collected the data. This is a multisite, longitudinal study designed to recruit more than 10,000 children aged 9–10 and follow them over 10 years into early adulthood. The ABCD Study® is supported by the National Institutes of Health and additional federal partners under award numbers U01DA041048, U01DA050989, U01DA051016, U01DA041022, U01DA051018, U01DA051037, U01DA050987, U01DA041174, U01DA041106, U01DA041117, U01DA041028, U01DA041134, U01DA050988, U01DA051039, U01DA041156, U01DA041025, U01DA041120, U01DA051038, U01DA041148, U01DA041093, U01DA041089, U24DA041123, U24DA041147. A full list of supporters is available at https://abcdstudy.org/federal-partners.html. A listing of participating sites and a complete listing of the study investigators can be found at https://abcdstudy.org/consortium_members/. ABCD consortium investigators designed and implemented the study and/or provided data but did not necessarily participate in the analysis or writing of this report. This manuscript reflects the views of the authors and may not reflect the opinions or views of the NIH or ABCD consortium investigators. In addition to the ABCD Study, this work was also supported by funding from the National Institutes of Health: NIEHS T32ES013678 (Gauderman and McConnell, trainee: Campbell), NIEHS R01- ES032295 (Herting), and NIMH F31MH131347 (Campbell). Also, the Advanced Statistical Methods in Neuroimaging and Genetics (NINDS R25NS117281) was crucial in developing the polygenic risk score calculations, with special recognition to Dr. Andrey Shabalin. The authors acknowledge the Center for Advanced Research Computing (CARC) at the University of Southern California for providing computing resources that have contributed to the research results reported within this publication.

