## Supplemental File for "Effects of depression genetic risk and household socioeconomic status on emotional behavior and brain development in early adolescence"

### Supplemental Material for Campbell et al.

### A. Methods

#### A.1 General Exclusionary Criteria for the Complete Sample

The ABCD Study® is intended to represent a population-based, non-clinical sample (exclusion criteria included severe sensory, intellectual, medical, and neurological disorders; sample recruitment, inclusion criteria, and protocol have been published in detail (1,2). Next, for this research, subjects assessed at site 22 were removed, since this site was not assessed long term. Next, given this works mental health focus, data collected after March 1st, 2020 were removed to avoid the confounding effects from the COVID-19 pandemic (3–5). Subjects who had poor genetic data (see section B.2.3 in main manuscript), no income-to-needs values (see section B.2.4 in main manuscript), and whose caregiver incorrectly entered their sex-at-birth were also excluded.

#### A.2 Passing Scores for MRI Data

For both time points, a passing score for the T1-weighted image meant the ABCD Study variable "imgincl_t1w_include" was equal to 1, plus no abnormal findings or normal anatomical variant of no clinical significance on the MRI image (mrif_score=2 or 3) based on ABCD Study rating values (6). For the rs-fMRI sample, a passing T2-weighted image (imgincl_t2w_include=1) was also needed given the inclusion of the T2-weighted image in the rs-fMRI functional calculations (6).

#### A.3 Neuroimaging Data

##### *A.3.1 Resting-State Functional MRI Connectivity*

Functional connectivity was measured using resting-state functional magnetic resonance imaging (rs-fMRI) data. These data were collected while the participant passively viewed a cross hair image on a screen for up to 20 minutes, split up into four 5-minute sessions (7). Each rs-fMRI scan was acquired at 2.4mm isotropic (TR=800ms) with a T2*-weighted image with a multiband EPI with slice acceleration; for each scan a fieldmap was also collected for $B_0$ distortion correction (7). Functional correlations for each region of interest (ROI) were then calculated based on the Fisher transformed z-statistic averaged within and between the networks (8).

##### *A.3.2 Structural MRI*

Structural MRI (sMRI) data was measured using the $T_1$-weighted image collected at 1-mm isotropic (7) via an inverted RF-spoiled gradient echo scan sequence utilizing prospective motion correction (9). Cortical and subcortical segmentation were completed using the semi-automated FreeSurfer v. 7.1.1 brain segmentation protocol (10–19). Again, all images underwent manual quality check post-acquisition and post-segmentation with FreeSurfer by the ABCD Study.

#### A.4 Genetic Data

##### *A.4.1 Genetic Data Collection*

At the baseline visit, saliva samples were collected to assess each individual's genetic data by the ABCD Study (20). DNA is isolated from saliva and genotyped using the Affymetric NIDA Smokescreen™ Genotyping array that has over 600,000 single nucleotide polymorphisms (SNPs) (21). DNA quality control was then completed by Rutgers RUCDR on all calling signals and variant call rates. The genotype dataset was then imputed using the TOPMed imputation server (22). These data are then made available in PLINK format by the consortium for further follow-up analysis.

##### *A.4.2 Depression Polygenic Risk Score (D-PRS) Generation*

To calculate D-PRS values, the discovery sample (23) had 9.6 million SNPs with 135,458 subjects with depression and 344,901 controls. This sample was made up of seven cohorts from numerous countries (Australia, Denmark, Germany,

Iceland, Italy, Sweden, Switzerland, The Netherlands, The United Kingdom, and The United States). For details on how the PRS weights were generated please refer to the original research article (23). To calculate the D-PRS with similar parameters to the pediatric cohort replication study (24), using the ABCD Study® post-imputation files, bfctools (25) was used to filter the files with a call rate <98% and minor allele frequency (MAF) < 0.05 were excluded; a Hardy-Weinberg Equilibrium (HWE) p < 0.0001 was not excluded given that this is done for homogenous sample, and the ABCD Study is a non-homogenous group of subjects. Utilizing plink2 (26) (www.cog-genomics.org/plink/2.0/) and R, samples with extreme heterozygosity - which could indicate poor quality sample (high heterozygosity) or inbreeding (low heterozygosity) (27) - were removed (28). Next, to generate the D-PRS values, we utilized the software program PRSice-2 (29) with SNPs having an $r^2$ > 0.25 with the index SNP being removed and SNPs within 200k of the index SNP considered for clumping.

With this data, principal components (PCs) of the sample's ancestry were generated (30). These PCs were used to account for population stratification that is inherent within genome-wide association studies (GWAS). To generate the PCs, plink2 was used to generate the first 50 PCs with a pruning window size of 200 variants, a 50 variant step size, and filtering out any SNPs with a linkage disequilibrium (LD) $r^2$ > 0.25. This was conducted within the European-like sample and not European-like sample separately with the greatest number of subjects available for each group.

##### A.5 Incomes-to-Needs Ratio (INR)

Family size and household income are assessed every year starting at the baseline visit within the ABCD Study. For family size, the caregiver reported the number of individuals living at the household as an integer value; the few individuals who responded with 0 or 1 for household size were converted to NA given the invalid nature of these answers (i.e., one caregiver plus child would indicate a minimum household of 2). They also reported the household's combined total income based on several binned categories: 1 = Less than $5,000; 2 = $5,000 through $11,999; 3 = $12,000 through $15,999; 4 = $16,000 through $24,999; 5 = $25,000 through $34,999; 6 = $35,000 through $49,999; 7 = $50,000 through $74,999; 8 = $75,000 through $99,999; 9 = $100,000 through $199,999; 10 = $200,000 and greater. Binned categories were converted to a continuous variable, mean-adjusted, and divided by the federal poverty level for the corresponding year (31).

##### A.6 Race/Ethnicity Variable

When asked to the caregiver, the race/ethnicity variable is defined as: non-Hispanic white, non-Hispanic Black, non-Hispanic Asian (Asian Indian, Chinese, Filipino, Japanese, Korean, Vietnamese, other Asian), Hispanic, other (American Indian, Native American, Alaska Native, Native Hawaiian, Guamanian, Samoan, other Pacific Islander, or other race) (32).

##### A.7 Data Analysis: Statistical Model Details

For our longitudinal mixed effects models, let $Y_{ij}$ represent the outcome for each model (behavioral or neuroimaging outcomes) for participant $i$ and visit $j$. Let $T_{ij}$ denote age and $X_{ij}$ a set of time-varying adjustment variables (age and study site/MRI serial number). The quantities of interest in the Level 1 model are the subject-specific intercepts ($a_i$, representing mean of the outcome at age $T_*$) and the slopes ($b_i$, representing the longitudinal change in $Y$ per year of age). Level 2 consists of separate regression models for the subject-specific intercepts (Level 2a) and slopes (Level 2b). The regressors of primary interest in these models are $Z_i$, the D-PRS for each individual and, $W_i$, represents the average INR across three study visits, and their interaction ($W_i Z_i$). These models also included adjustments for time-invariant covariates, $X_i$ (i.e., sex-at-birth, maximum highest-level of parents' education across all time points, caregiver-identified race/ethnicity, and the first 10 genetic PCs) and a random effect of subject nested within site:

Level 1 (visit): $$Y_{ij} = a_i + b_i(T_{ij} - T_*) + \delta_1 X_{ij} + \epsilon_{ij}$$

Level 2a (intercepts): $a_i = a + \beta_1 Z_i + \beta_2 W_i + \beta_3 W_i Z_i + \delta_2 X_i + e_i$

Level 2b (slopes): $b_i = b + \theta_1 Z_i + \theta_2 W_i + \theta_3 W_i Z_i + \delta_3 X_i + f_i$

The Level 2 expressions for $a_i$ and $b_i$ are substituted into the Level 1 model and the resulting single mixed-effect linear model is used to estimate the parameters of interest. The baseline levels ($\beta$'s) and rate of change for the participants ($\theta$'s) were assessed in terms of the interactive influence of D-PRS and INR. For where the individual starts for D-PRS, INR, and their interaction, we examined $\beta_1$, $\beta_2$, and $\beta_3$, respectively; for how they change over time, we examined $\theta_1$, $\theta_2$ and $\theta_3$, respectively. Utilizing $T_*$, we could set Age to values of interest. In addition, since the neuroimaging models had numerous outcomes, these models were corrected for multiple comparisons with a False Discovery Rate of $\alpha$<0.05. Finally, given the complexity of our final models, higher order terms were dropped in a stepwise manner in favor of a parsimonious model if they were not significant. First, if INR-by-PRS-by-Age was insignificant it was removed; next, if not significant, INR-by-Age and/or PRS-by-Age were removed. Finally, INR-by-PRS was removed if not significant to test the main effects of INR and PRS. For the final parsimonious models for each outcome see **Supplemental Table 12**.

#### B. Results

##### B.1 Withdrawn/Depressed Symptoms: Depression Polygenic Risk Score Effects

For the European-like sample, a main effect of D-PRS (Incidence Risk Ratio (IRR)=1.184, p=1.1x10- 8) was found (**Figure 2A**; **Supplemental Table 13**). A non-linear relationship was seen between D-PRS and withdrawn/depressed symptoms with higher D-PRS values leading to greater symptom counts. On average, an individual with a mean D-PRS in our European-like sample, has a score of 0.74 out of a possible 16 points, meaning they have little-to-no withdrawn/depressed symptoms. If an individual has a lower D-PRS (2 standard deviations (SD) below the mean), this led to a 0.21 (29%) decrease in their symptom score to 0.53, meaning less depressed/withdrawn symptoms. A higher D-PRS (2 SD above the mean), would lead to a 0.30 (40%) increase as compared to mean D-PRS, to a score of 1.03. Though, if an individual had a D-PRS towards the upper bounds of the range (6 SD above the mean), they would have a 1.29 (175%) increase compared to the mean, having an estimated score of 2.03. To put these results into perspective, if a 10-year-old female has a mean D-PRS value - based on our model - the CBCL score would range from 0-1 points (50-52 t-score); with a score of 0-1, these symptoms are not clinically relevant102. If a 10-year-old female has a D-PRS 6 standard deviations above the mean, the CBCL score would be ~2 points (56 t-score), which would indicate that the symptoms are still not clinically relevant. Next, for the not European-like Group 1 sample, there is a small main effect of D-PRS (IRR=1.223, p=0.028) (**Figure 2A**; **Supplemental Table 13).** While this same effect was seen in the European-like sample, this effect is deprecated in European-like Group 1. For the mean D-PRS value, an adolescent in the not European-like Group 1, would have an estimated withdrawn/depressed symptom score of 0.71. As the D-PRS decreased (2 SD below the mean), the predicted score decreased by 0.23 (33%) to 0.47, while an increase (2 SD above the mean) led to an increase of 0.35 points (49%) to an estimated score of 1.06, which is not clinically relevant. Given the smaller range of D-PRS observed in this group, the larger effects seen at high D-PRS in the European-like sample were not observed (**Supplemental Figure 10**). For the not European-like Group 2, no significant effects of D-PRS were found for the withdrawn/depressed symptom score (**Figure 2A**; **Supplemental Table 13**). Given that no significant D-PRS by age interaction terms were seen for any of the groups, it suggests this genetic effect is consistent over time between the ages of 9-12 years-old (i.e., from baseline to follow-up sessions).

##### B.2 Positive Affect: Depression Polygenic Risk Score Effects

In the European-like sample, there was a main effect of D-PRS ($\beta$=-0.181, p=0.002,), with higher D-PRS leading to lower positive affect scores (**Figure 3**; **Supplemental Table 14**). Specifically, a 1 standard deviation increase in a D-PRS decreased self-report of positive affect score by 0.181 points, which is minimal given the range of the potential points

(9-27 points). In the not European Group 1 ($\beta$=-0.121, p=0.516) and Group 2 ($\beta$=-0.182, p=0.245) samples, a similar trend was seen, albeit was not significant, potentially due to the minimized variance in D-PRS values generated for youths in these groups. Given that no significant D-PRS-by-age interaction terms were seen for any of the groups, suggests this genetic effect is consistent over time between the ages of 9-12 years-old (i.e., from 6-month follow-up to 18-month follow-up sessions).

### D. Figures

**Supplemental Figure 1: General exclusionary flowchart of complete sample prior to outcome groupings**

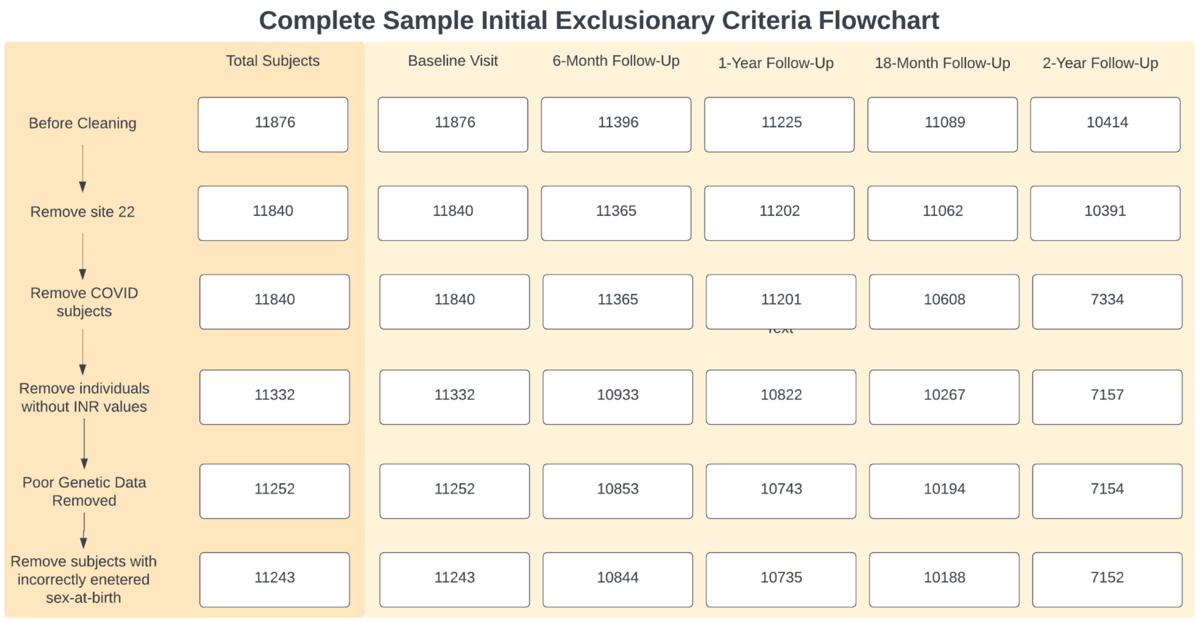

By study design, baseline visit corresponds to ages 9-10 years, 1-year follow-up visit corresponds to ages 10-11 years, 2-year follow-up visit corresponds to ages 11-12 years and are conducted in person, whereas 6-month follow-up and 18-month follow-up are conducted by phone. Abbreviations: INR = income-to-needs ratio.

**Supplemental Figure 2: Flowchart of exclusionary criteria for emotional behavior analyses**

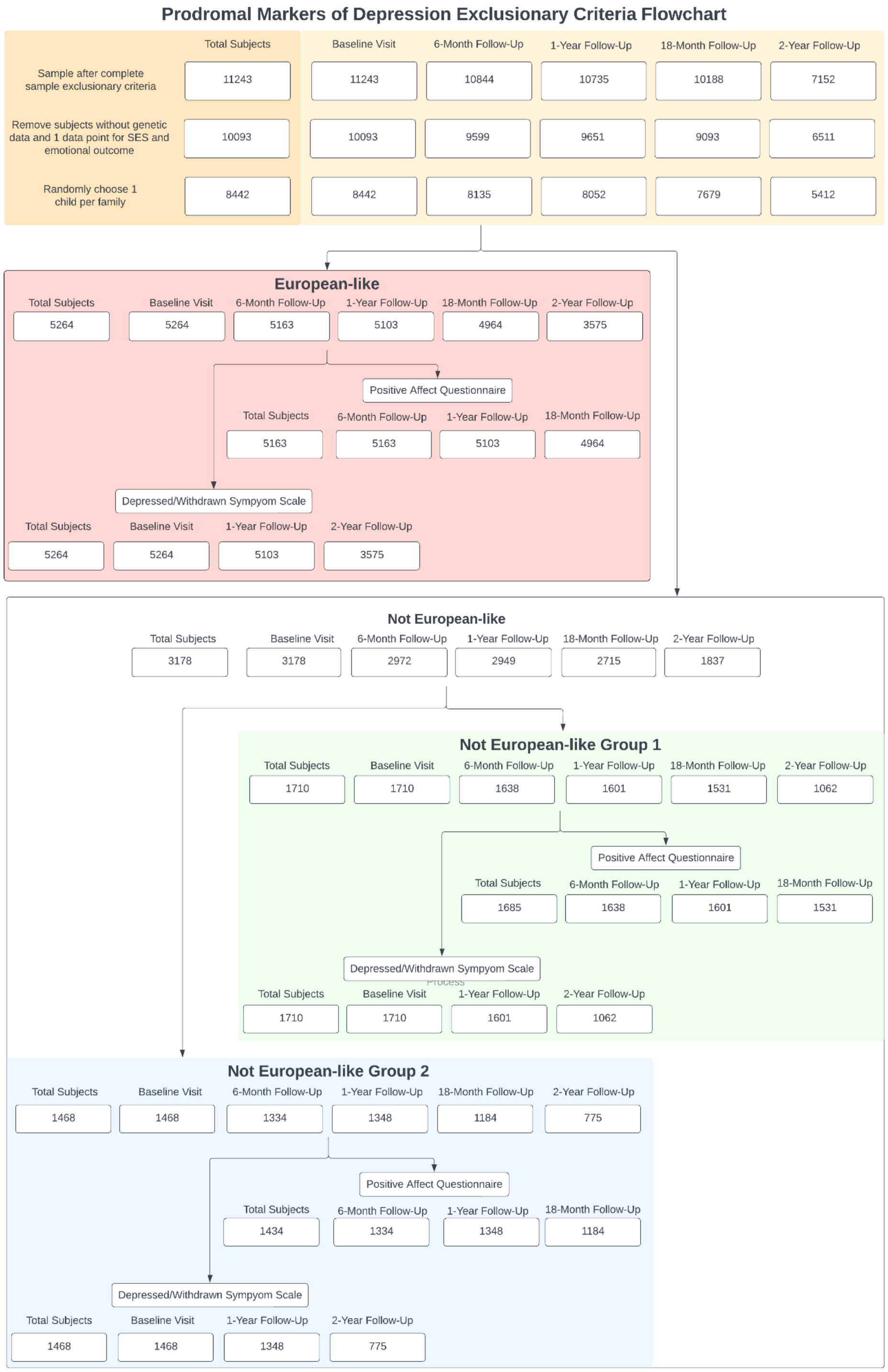

Flowchart of exclusionary criteria for Depressed/Withdrawn Symptom Scale and Positive Affect Questionnaire after exclusionary criteria is completed for the full sample (reference Supplemental Figure 1). By study design, baseline visit corresponds to ages 9-10 years, 1-year follow-up visit corresponds to ages 10-11 years, 2-year follow-up visit corresponds to ages 11-12 years and are conducted in person, whereas 6-month follow-up and 18-month follow-up are conducted by phone. Abbreviations: SES = socioeconomic status as measured by income-to-needs ratio (INR).

**Supplemental Figure 3: Flowchart of exclusionary criteria for rs-fMRI analyses**

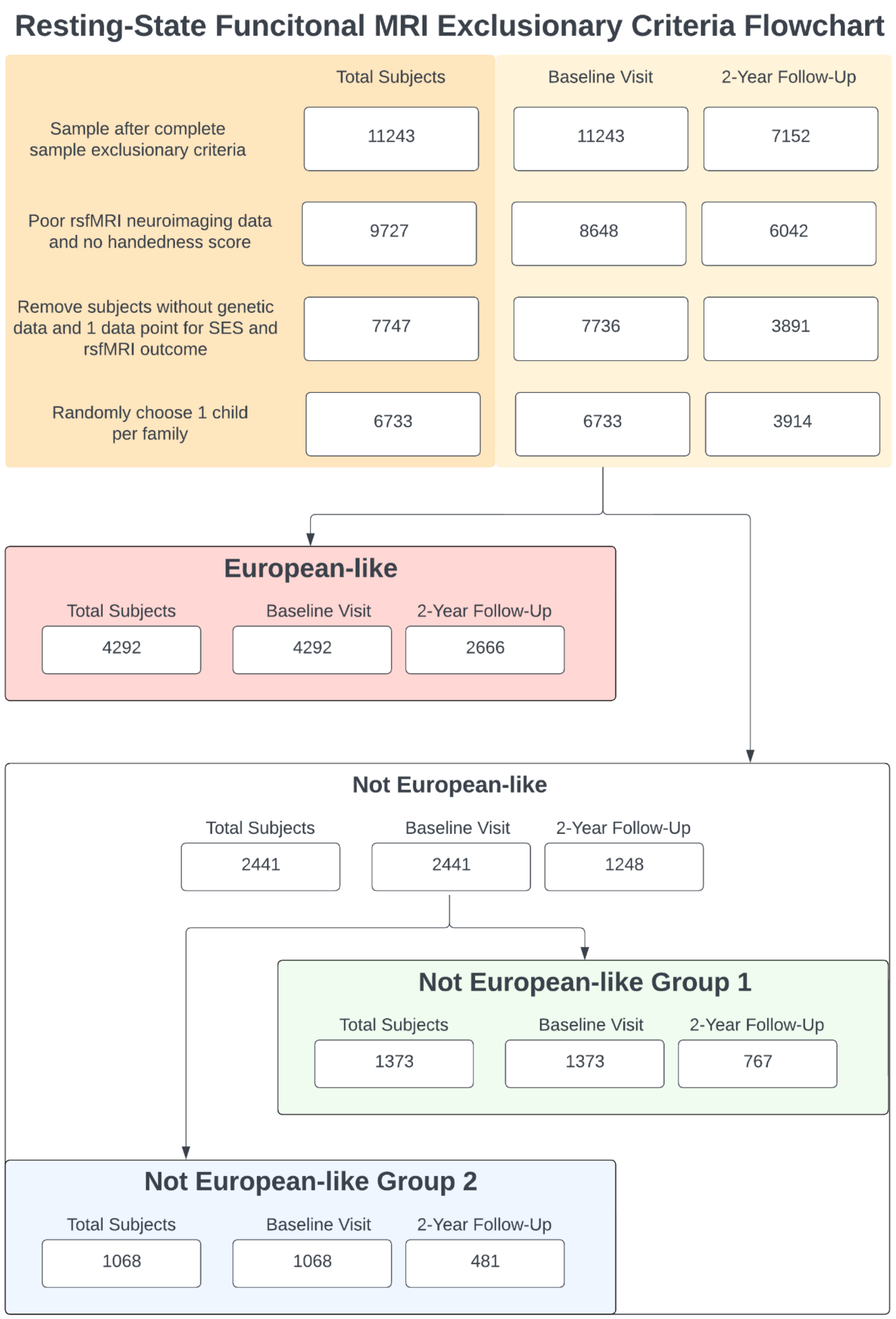

Flowchart of exclusionary criteria for resting-state functional MRI analyses after exclusionary criteria is completed for the full sample (reference Supplemental Figure 1). By study design, baseline visit corresponds to ages 9-10 years and 2-year follow-up visit corresponds to ages 11-12 years; both are conducted in-person. Abbreviations: SES = socioeconomic status; rs-fMRI = resting-state functional magnetic resonance imaging.

**Supplemental Figure 4: Flowchart of exclusionary criteria for sMRI analyses**

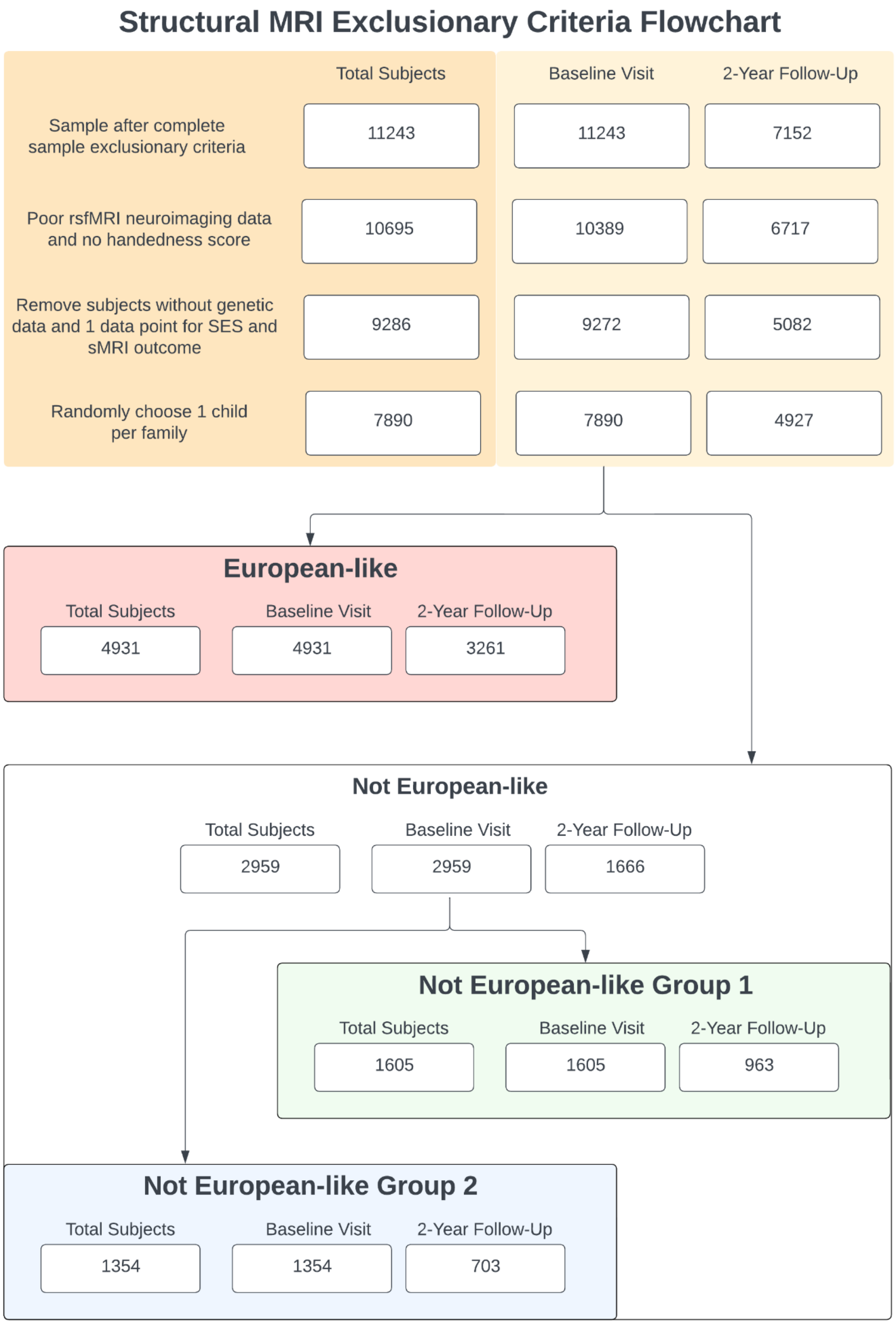

Flowchart of exclusionary criteria for structural MRI analyses after exclusionary criteria is completed for the full sample (reference Supplemental Figure 1). By study design, baseline visit corresponds to ages 9-10 years and 2-year follow-up visit corresponds to ages 11-12 years; both are conducted in-person. Abbreviations: SES = socioeconomic status; sMRI = structural magnetic resonance imaging.

**Supplemental Figure 5: Location of brain regions associated with each of the large-scale brain networks**

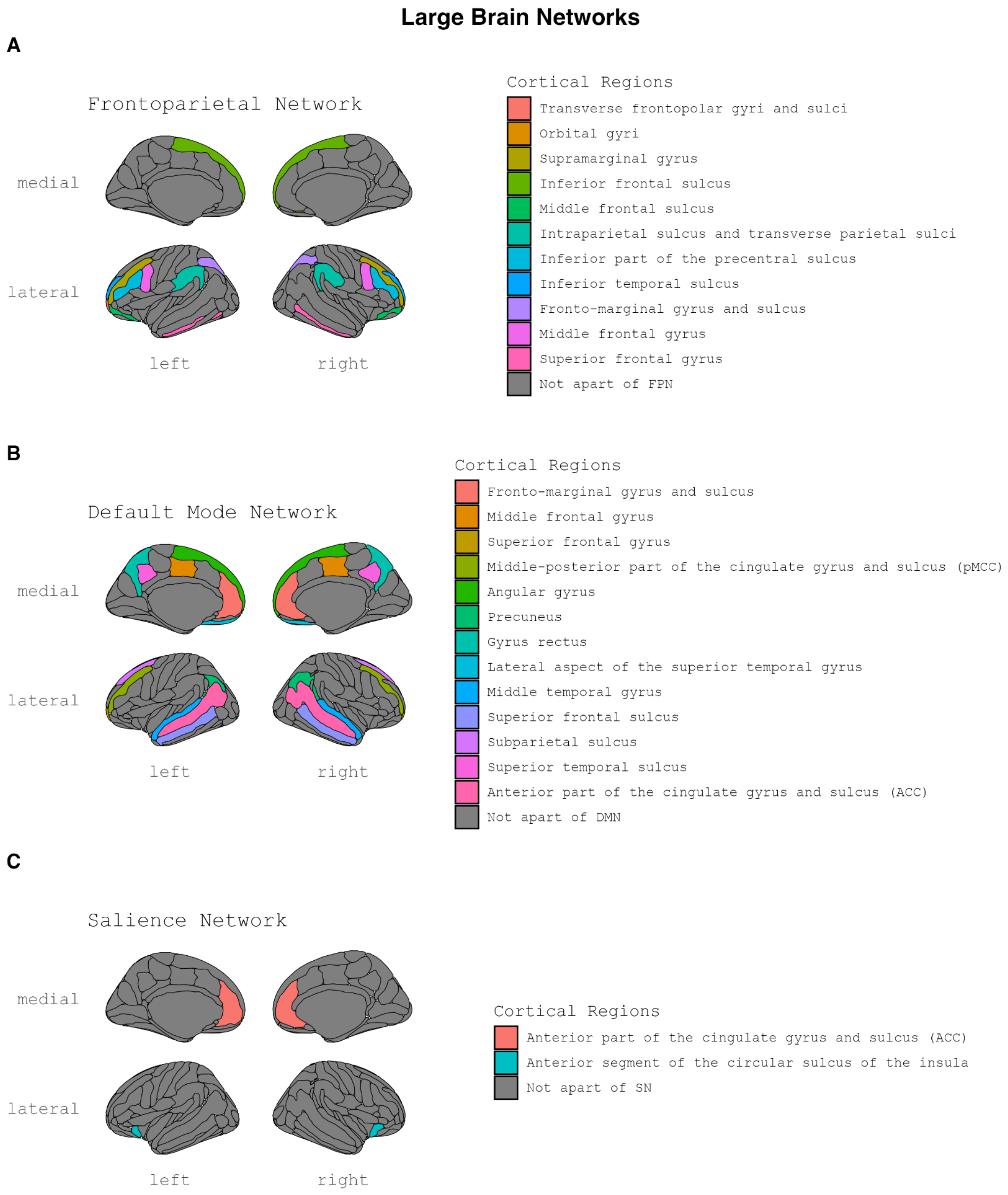

Representation of the large-scale brain networks in this research and their corresponding cortical regions; A) frontoparietal network (FPN), B) default mode network (DMN), C) salience network (SN).

**Supplemental Figure 6: Violin distributions with boxplots for withdrawn/depressed symptoms analyses**

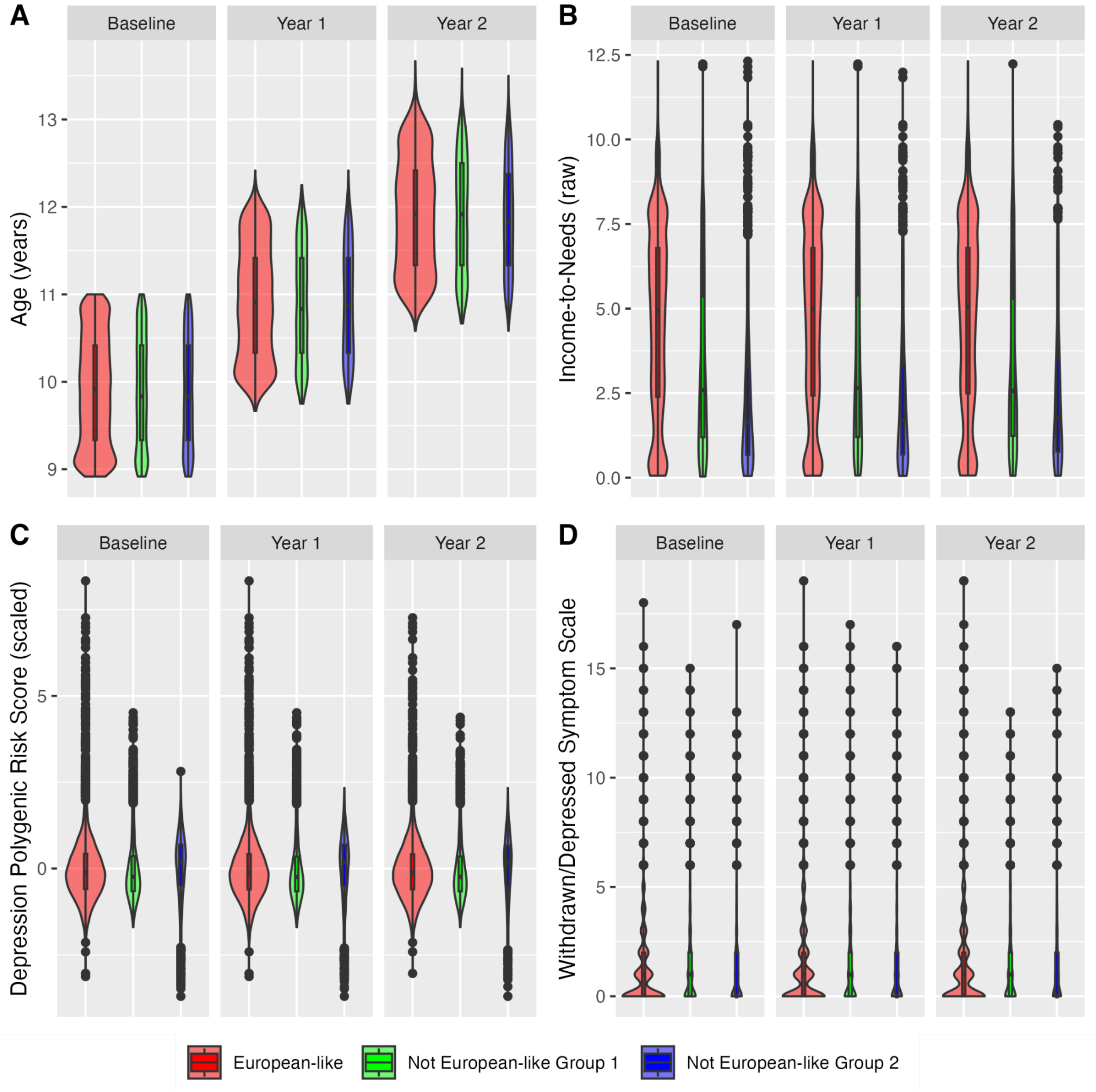

Violin distributions with boxplots of A) age, B) income-to-needs (INR), C) depression polygenic risk score (D-PRS), and D) withdrawn/depressed symptoms score reported by caregiver across all groups and by data collection year for depressed/withdrawn symptom scale analysis. By study design, baseline visit corresponds to ages 9-10 years, 1-year follow-up visit corresponds to ages 10-11 years, 2-year follow-up visit corresponds to ages 11-12 years and are conducted in-person. Note, D-PRS values are scaled within each sample.

**Supplemental Figure 7: Violin distributions with boxplots for positive affect analyses**

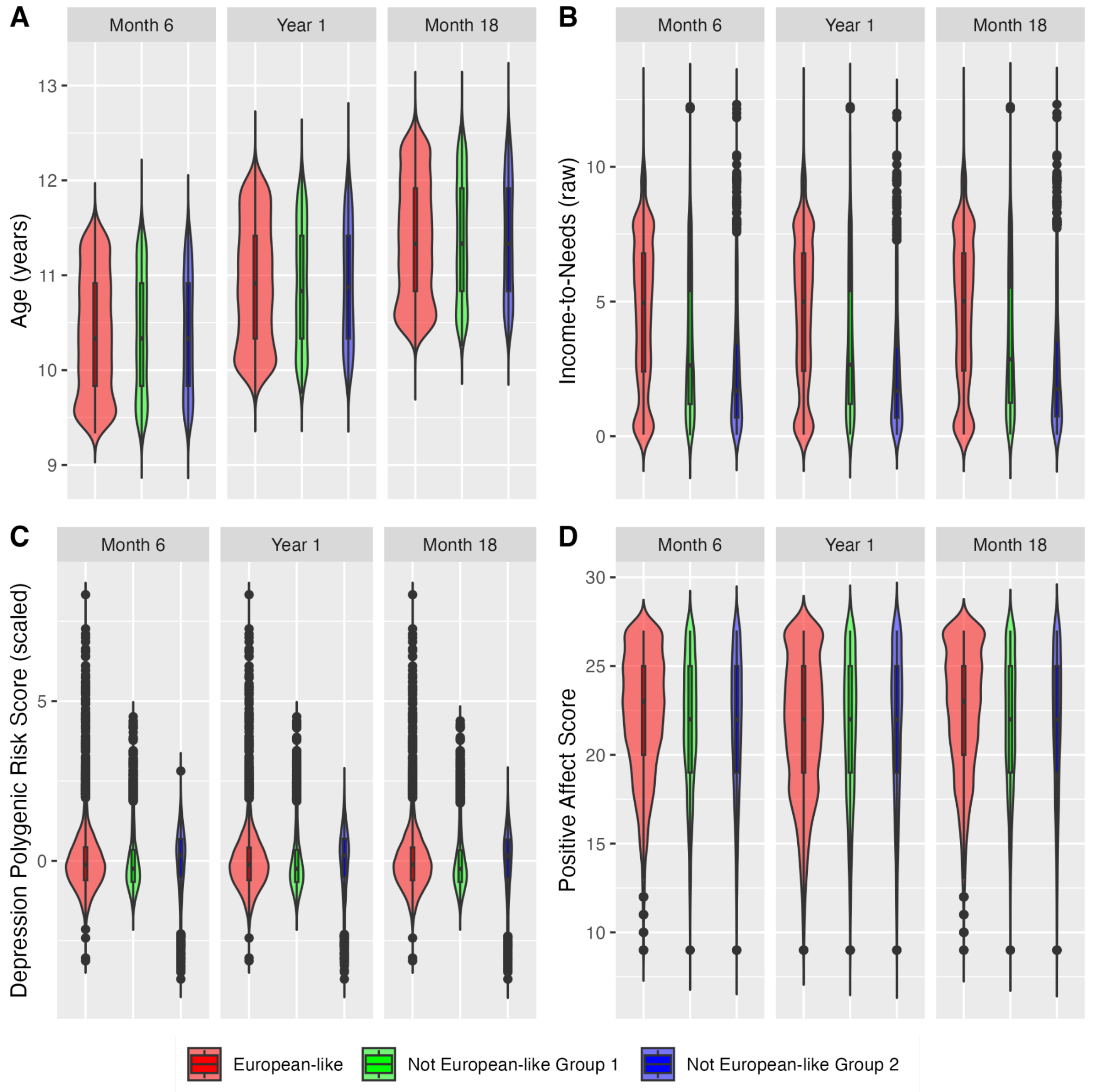

Violin distributions with boxplots of A) age, B) income-to-needs (INR), C) depression polygenic risk score (D-PRS), and D) positive affect score reported by youth across all groups and by data collection year for positive affect questionnaire analysis. By study design, baseline visit corresponds to ages 9-10 years, 1-year follow-up visit corresponds to ages 10-11 years, 2-year follow-up visit corresponds to ages 11-12 years and are conducted in person, whereas 6-month follow-up and 18-month follow-up are conducted by phone. Note, D-PRS values are scaled within each sample.

**Supplemental Figure 8: Violin distributions with boxplots for rs-fMRI analyses**

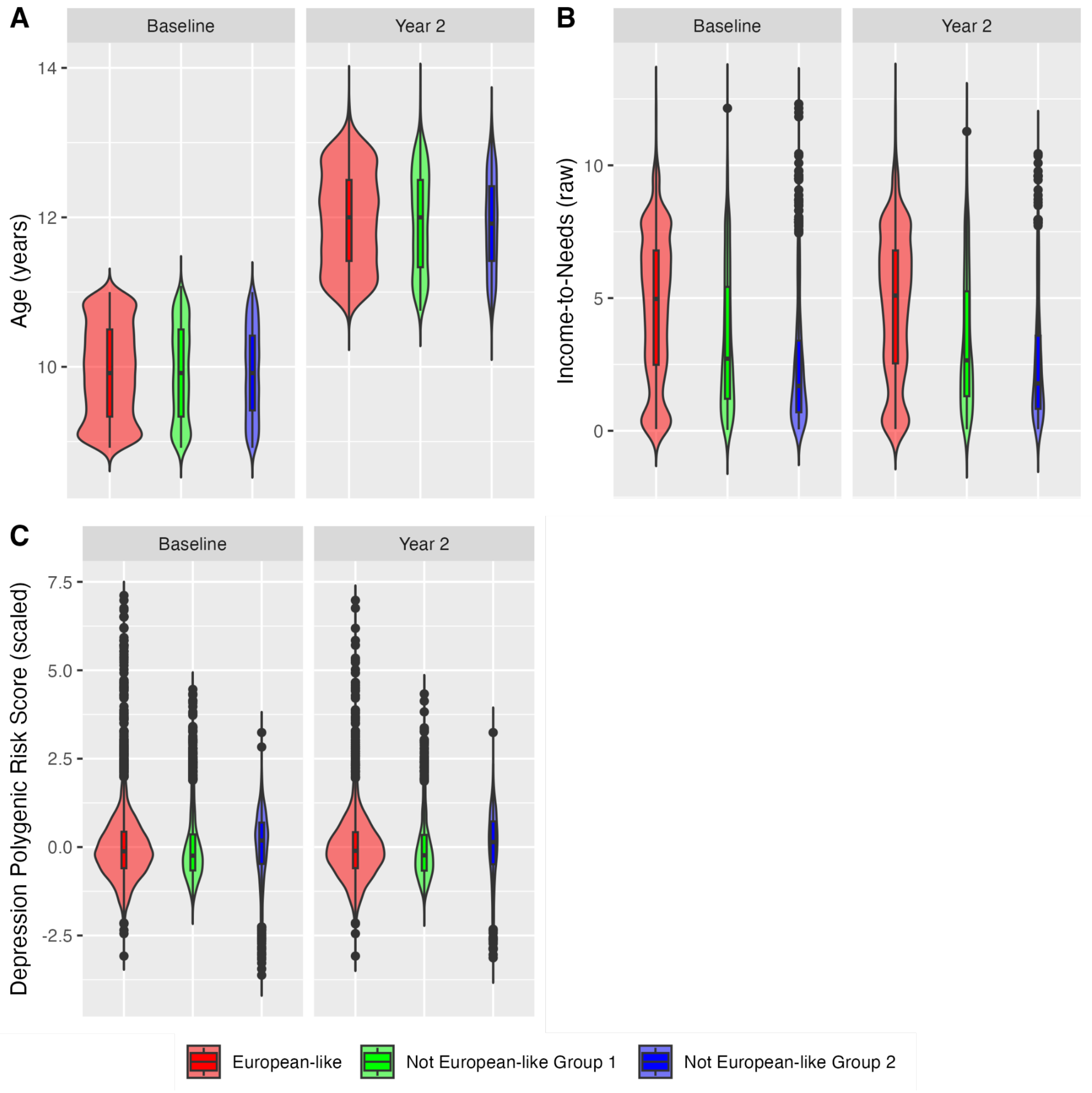

Violin distributions with boxplots of A) age, B) income-to-needs (INR), and C) depression polygenic risk score (D-PRS) across all groups and by data collection year for resting-state functional MRI analysis. By study design, baseline visit corresponds to ages 9-10 years and 2-year follow-up visit corresponds to ages 11-12 years; both are conducted in-person. Note, D-PRS values are scaled within each sample. Abbreviations: rs-fMRI = resting-state functional magnetic resonance imaging.

**Supplemental Figure 9: Violin distributions with boxplots for sMRI analyses**

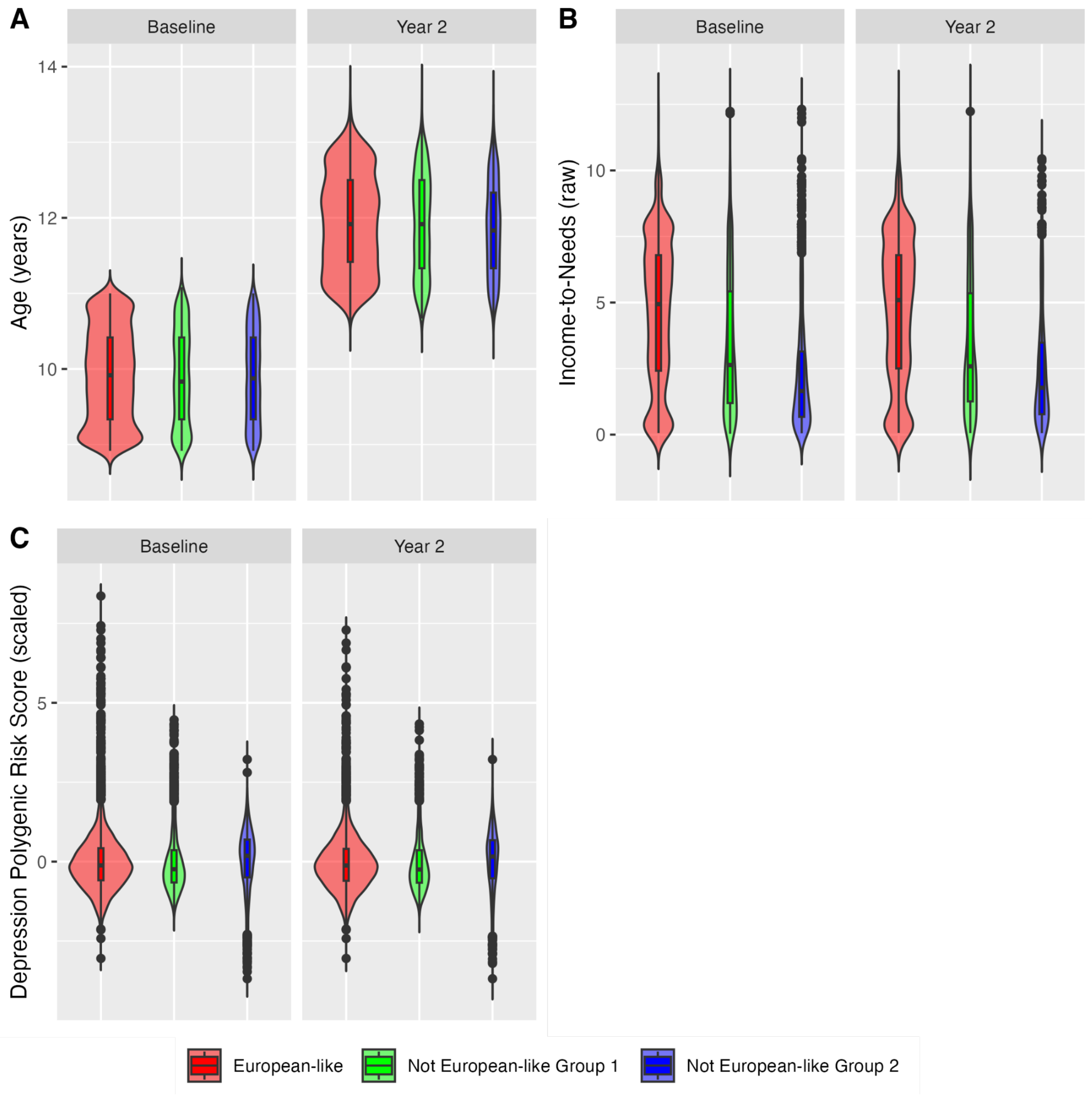

Violin distributions with boxplots of A) age, B) income-to-needs (INR), and C) depression polygenic risk score (D-PRS) across all groups and by data collection year for structural MRI analysis. By study design, baseline visit corresponds to ages 9-10 years and 2-year follow-up visit corresponds to ages 11-12 years; both are conducted in-person. Note, D-PRS values are scaled within each sample. Abbreviations: sMRI = structural magnetic resonance imaging.

**Supplemental Figure 10: Distribution of D-PRS by analysis group.**

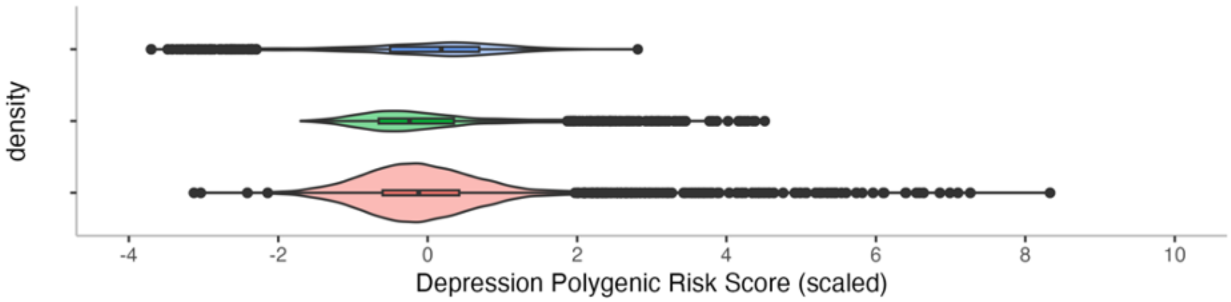

Note, CBCL withdrawn/depression symptom scale can range from 0–16 points (i.e., 8 questions). Abbreviations: $p_{D\text{-}PRS}$ = p-value for effect of D-PRS; n.s. = not significant.

**Supplemental Figure 11: Effect of INR on Positive Affect**

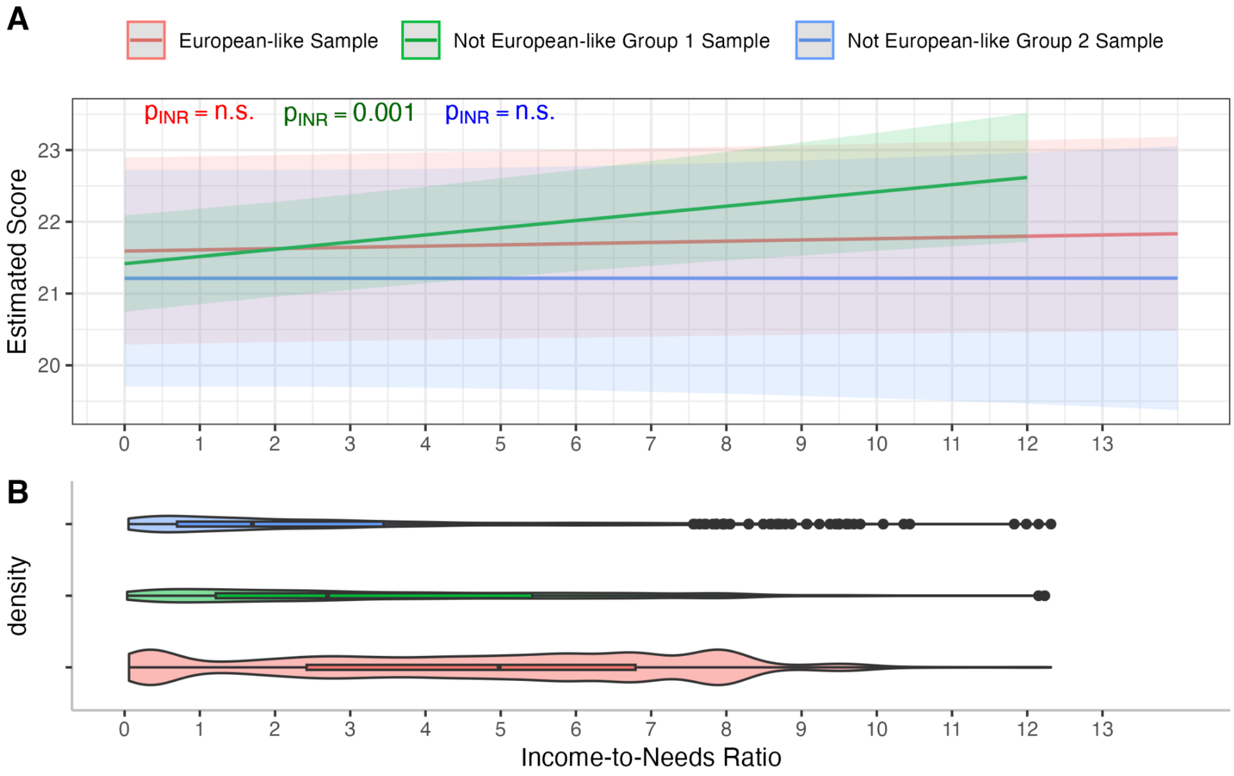

A) Effect of the income-to-needs ratio (INR) on the positive affect questionnaire in each group; B) Distribution of INR by analysis group. Note, Positive Affect scale can range from 9–27 points. Abbreviations: $p_{INR}$ = p-value for effect of INR; n.s. = not significant.

**Supplemental Figure 12: Trending effect of INR on middle temporal gyrus surface area in European-like Sample**

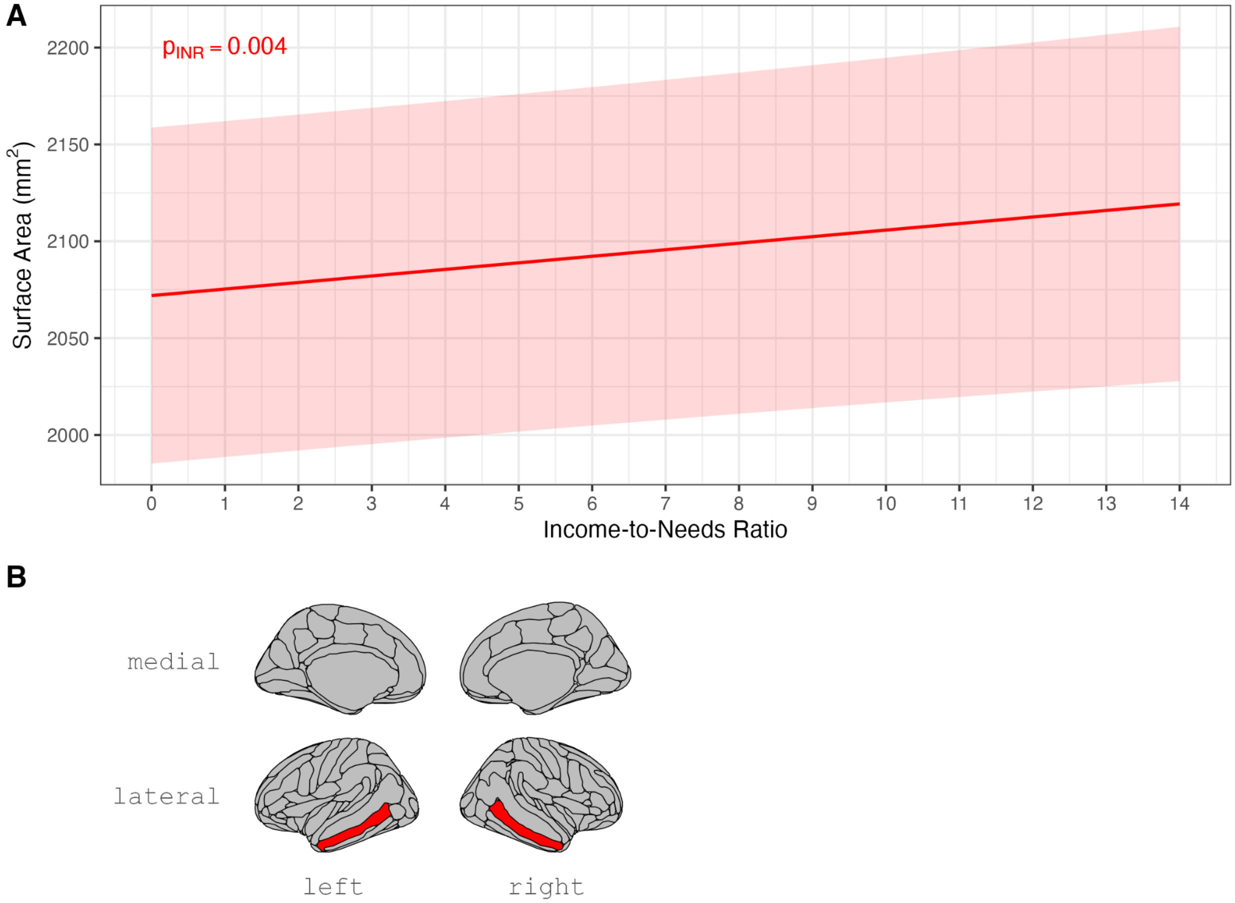

A) Trending effect of income-to-needs ratio (INR) on the surface area of the middle temporal gyrus in the European-like sample with B) corresponding visual of the middle temporal gyrus location. Age centered at 10 years; D-PRS mean-centered in the European-like sample; other covariates held constant (caregiver identified race/ethnicity, MRI scanner serial number, sex-at-birth, highest parents' education, handedness, first 10 genetic PCs). Abbreviations: mm$^2$, millimeters cubed; $p_{INR}$ = p-value for effect of INR.

**Supplemental Figure 13: Significant effect of INR on sMRI metrics in the not European-like Group 2 Sample**

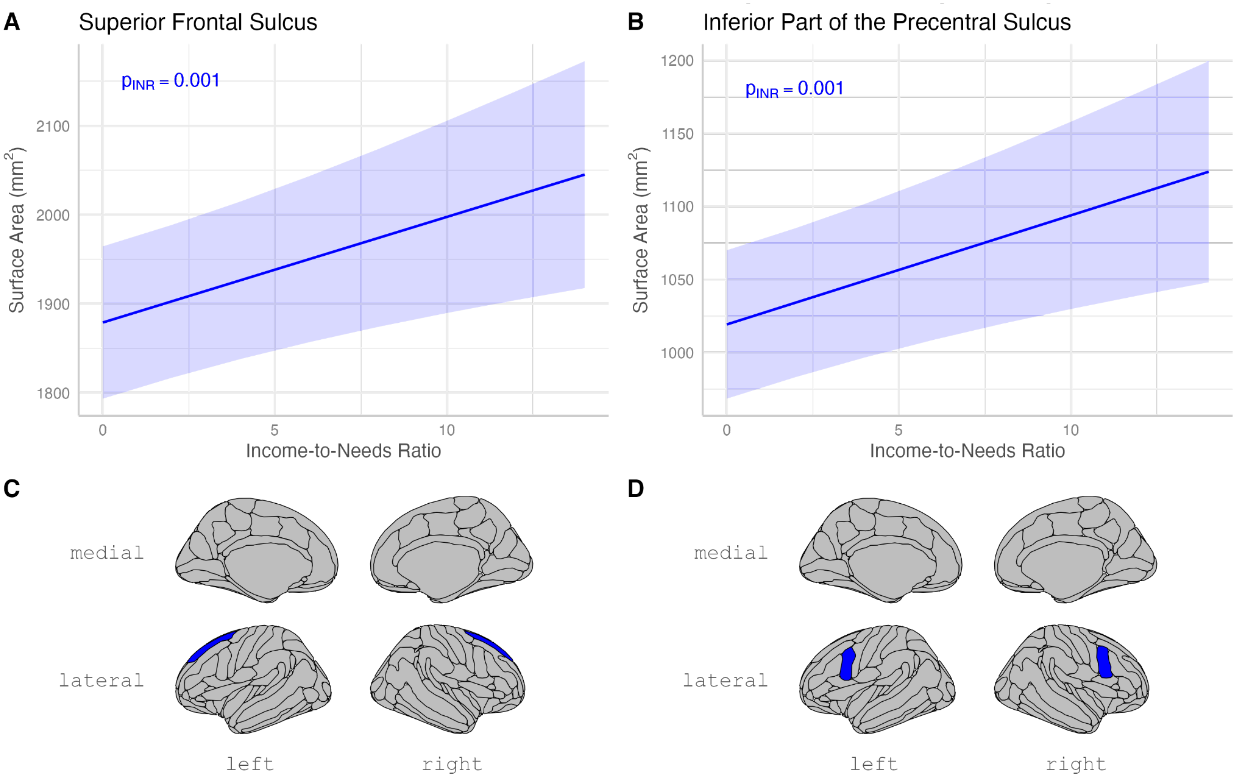

A-B) Effect of income-to-needs ratio (INR) on the surface area of the above structural MRI metrics in the not European-like Group 2 sample with C-D) corresponding visual of the brain region location below each graph. Age centered at 10 years; covariates held constant (caregiver identified race/ethnicity, MRI scanner serial number, sex-at-birth, highest parents' education, handedness). Abbreviations: $p_{INR}$ = p-value of INR effect; sMRI = structural magnetic resonance imaging.

**Supplemental Figure 14: Trending effect of INR on sMRI metrics in the not European-like Group 2 Sample**

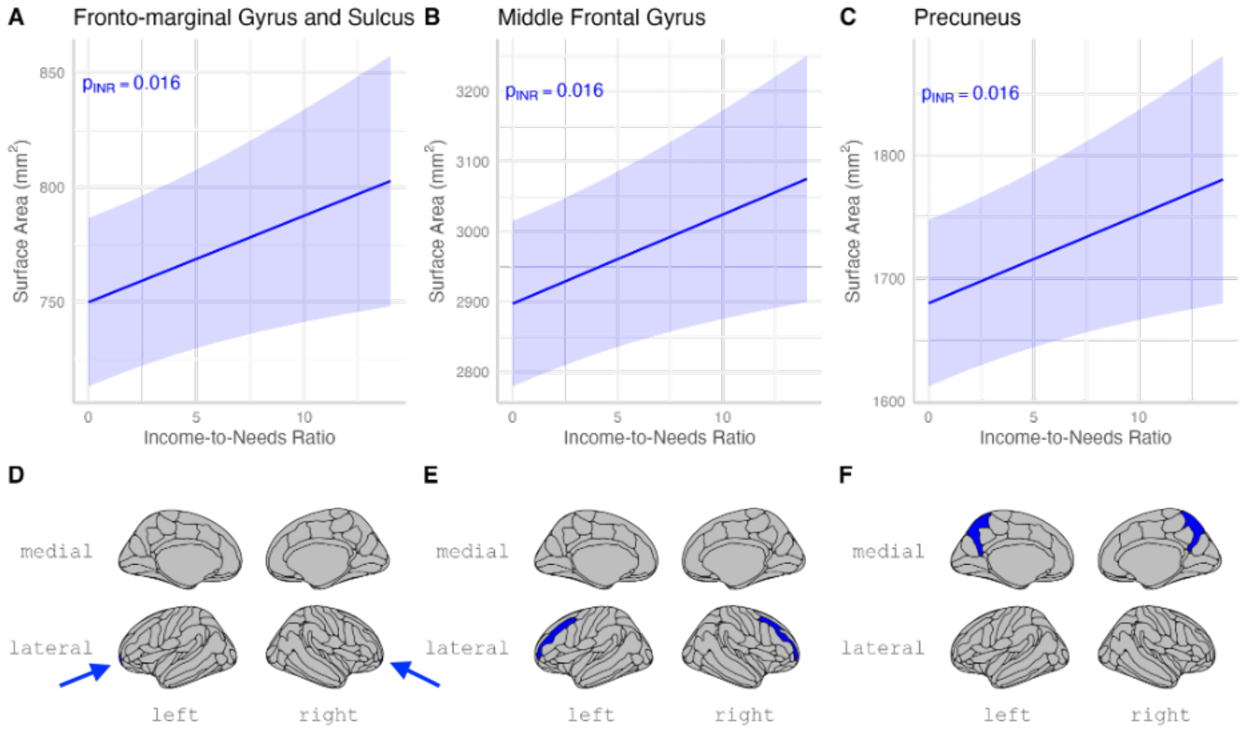

A-C) Trending effect of income-to-needs ratio (INR) on the surface area of the structural MRI metrics in the not European-like Group 2 sample with D-F) corresponding visual of the brain region location below each graph. Age centered at 10 years; PRS mean-centered in the sample; other covariates held constant (caregiver identified race/ethnicity, MRI scanner serial number, sex-at-birth, highest parents' education, handedness). Abbreviations: $p_{INR}$ = p-value of INR effect; sMRI = structural magnetic resonance imaging.

### E. Tables

#### Supplemental Table 1: Cortical region names and their corresponding functional networks

| Region | Destrieux Atlas Short Name | FPN | DMN | SN |
|---|---|---|---|---|
| Transverse frontopolar gyri and sulci | G and S transv frontopol | ☑ | | |
| Orbital gyri | G orbital | ☑ | | |
| Supramarginal gyrus | G pariet inf-Supramar | ☑ | | |
| Inferior frontal sulcus | S front inf | ☑ | | |
| Middle frontal sulcus | S front middle | ☑ | | |
| Intraparietal sulcus (interparietal sulcus) and transverse parietal sulci | S intrapariet and P trans | ☑ | | |
| Inferior part of the precentral sulcus | S precentral-inf-part | ☑ | | |
| Inferior temporal sulcus | S temporal inf | ☑ | | |
| Fronto-marginal gyrus (of Wernicke) and sulcus | G and S frontomargin | ☑ | ☑ | |
| Middle frontal gyrus (F2) | G front middle | ☑ | ☑ | |
| Superior frontal gyrus (F1) | G front sup | ☑ | ☑ | |
| Middle-posterior part of the cingulate gyrus and sulcus (pMCC) | G and S cingul-Mid-Post | | ☑ | |
| Angular gyrus | G pariet inf-Angular | | ☑ | |
| Precuneus (medial part of P1) | G precuneus | | ☑ | |
| Straight gyrus, Gyrus rectus | G rectus | | ☑ | |
| Lateral aspect of the superior temporal gyrus | G temp sup-Lateral | | ☑ | |
| Middle temporal gyrus (T2) | G temporal middle | | ☑ | |
| Superior frontal sulcus | S front sup | | ☑ | |
| Subparietal sulcus | S subparietal | | ☑ | |
| Superior temporal sulcus (parallel sulcus) | S temporal sup | | ☑ | |
| Anterior part of the cingulate gyrus and sulcus (ACC) | G and S cingul-Ant | | ☑ | ☑ |
| Anterior segment of the circular sulcus of the insula | S circular insula ant | | | ☑ |

This table represents the cortical regions that correspond to the frontoparietal network (FPN), default mode network (DMN), and the salience network (SN); the corresponding Destrieux atlas short name is also listed. Abbreviations: G = Gyrus; S = Sulcus.

**Supplemental Table 2: Withdrawn/Depressed Symptoms Analyses Demographics for Baseline Data Collection**

Comparison of Depressed/Withdrawn Sample Across Cohorts - Baseline

| | European-like (N=5264) | Not European-like Group 1 (N=1710) | Not European-like Group 2 (N=1468) | p value |
|---|---|---|---|---|
| **Age (years)** | | | | 0.745 |
| Mean (SD) | 9.902 (0.620) | 9.892 (0.621) | 9.891 (0.606) | |
| Range | 8.917 - 11.000 | 8.917 - 11.000 | 8.917 - 11.000 | |
| **INR** | | | | < 0.001 |
| Mean (SD) | 4.619 (2.772) | 3.366 (2.595) | 2.385 (2.271) | |
| Range | 0.059 - 12.315 | 0.033 - 12.233 | 0.052 - 12.315 | |
| **D-PRS (scaled)** | | | | 0.966 |
| Mean (SD) | 0.010 (1.015) | 0.014 (1.013) | 0.017 (0.993) | |
| Range | -3.128 - 8.331 | -1.701 - 4.512 | -3.700 - 2.813 | |
| **Withdrawn/Depressed Symptom Scale** | | | | 0.209 |
| N-Miss | 1 | 0 | 0 | |
| Mean (SD) | 1.284 (1.978) | 1.308 (2.068) | 1.191 (1.986) | |
| Range | 0.000 - 18.000 | 0.000 - 15.000 | 0.000 - 17.000 | |
| **Sex at Birth** | | | | 0.017 |
| Female | 2476 (47.0%) | 765 (44.7%) | 731 (49.8%) | |
| Male | 2788 (53.0%) | 945 (55.3%) | 737 (50.2%) | |
| **Highest Caregiver's Education** | | | | < 0.001 |
| N-Miss | 0 | 0 | 1 | |
| Less Than HS | 31 (0.6%) | 162 (9.5%) | 60 (4.1%) | |
| HS Diploma/GED | 213 (4.0%) | 218 (12.7%) | 328 (22.4%) | |
| Some College | 1936 (36.8%) | 764 (44.7%) | 766 (52.2%) | |
| Bachelor | 928 (17.6%) | 208 (12.2%) | 139 (9.5%) | |
| Post-Grad | 2156 (41.0%) | 358 (20.9%) | 174 (11.9%) | |
| **Caregiver-Identified Race/Ethnicity** | | | | < 0.001 |
| non-Hispanic white | 4644 (88.2%) | 54 (3.2%) | 1 (0.1%) | |
| non-Hispanic Black | 2 (0.0%) | 5 (0.3%) | 1247 (84.9%) | |
| Hispanic | 398 (7.6%) | 1206 (70.5%) | 76 (5.2%) | |
| non-Hispanic Asian | 3 (0.1%) | 57 (3.3%) | 0 (0.0%) | |
| other | 217 (4.1%) | 388 (22.7%) | 144 (9.8%) | |
| **Race/Ethnicity Categories for Modeling** | | | | < 0.001 |
| non-Hispanic white | 4644 (88.2%) | 0 (0.0%) | 0 (0.0%) | |
| Hispanic | 398 (7.6%) | 1206 (70.5%) | 0 (0.0%) | |
| other | 222 (4.2%) | 504 (29.5%) | 221 (15.1%) | |
| non-Hispanic Black | 0 (0.0%) | 0 (0.0%) | 1247 (84.9%) | |

For continuous variables, the p-value represents an ANOVA t-test with equal variances, while for categorical variables, it is a chi-squared p-value. Abbreviations: INR = income-to-needs; D-PRS = depression polygenic risk score; HS = high-school; SD = standard deviation.

**Supplemental Table 3: Withdrawn/Depressed Symptoms Analyses Demographics for 1-Year Follow-Up Data Collection**

Comparison of Depressed/Withdrawn Sample Across Cohorts - 1-Year Follow-Up

| | European-like (N=5103) | Not European-like Group 1 (N=1601) | Not European-like Group 2 (N=1348) | p value |
|---|---|---|---|---|
| **Age (years)** | | | | 0.589 |
| Mean (SD) | 10.913 (0.638) | 10.894 (0.638) | 10.906 (0.624) | |
| Range | 9.667 - 12.417 | 9.750 - 12.250 | 9.750 - 12.417 | |
| **INR** | | | | < 0.001 |
| Mean (SD) | 4.642 (2.764) | 3.389 (2.587) | 2.413 (2.272) | |
| Range | 0.059 - 12.315 | 0.064 - 12.233 | 0.052 - 11.989 | |
| **D-PRS (scaled)** | | | | 0.946 |
| Mean (SD) | -0.005 (0.995) | 0.001 (1.010) | 0.004 (0.996) | |
| Range | -3.128 - 8.331 | -1.701 - 4.512 | -3.700 - 2.338 | |
| **Withdrawn/Depressed Symptom Scale** | | | | 0.001 |
| N-Miss | 6 | 3 | 3 | |
| Mean (SD) | 1.483 (2.230) | 1.431 (2.228) | 1.234 (2.170) | |
| Range | 0.000 - 19.000 | 0.000 - 17.000 | 0.000 - 16.000 | |
| **Sex at Birth** | | | | 0.007 |
| Female | 2393 (46.9%) | 708 (44.2%) | 675 (50.1%) | |
| Male | 2710 (53.1%) | 893 (55.8%) | 673 (49.9%) | |
| **Highest Caregiver's Education** | | | | < 0.001 |
| N-Miss | 0 | 0 | 1 | |
| Less Than HS | 29 (0.6%) | 143 (8.9%) | 53 (3.9%) | |
| HS Diploma/GED | 195 (3.8%) | 203 (12.7%) | 294 (21.8%) | |
| Some College | 1854 (36.3%) | 715 (44.7%) | 702 (52.1%) | |
| Bachelor | 910 (17.8%) | 192 (12.0%) | 129 (9.6%) | |
| Post-Grad | 2115 (41.4%) | 348 (21.7%) | 169 (12.5%) | |
| **Caregiver-Identified Race/Ethnicity** | | | | < 0.001 |
| non-Hispanic white | 4511 (88.4%) | 48 (3.0%) | 1 (0.1%) | |
| non-Hispanic Black | 2 (0.0%) | 4 (0.2%) | 1143 (84.8%) | |
| Hispanic | 379 (7.4%) | 1120 (70.0%) | 73 (5.4%) | |
| non-Hispanic Asian | 3 (0.1%) | 55 (3.4%) | 0 (0.0%) | |
| other | 208 (4.1%) | 374 (23.4%) | 131 (9.7%) | |
| **Race/Ethnicity Categories for Modeling** | | | | < 0.001 |
| non-Hispanic white | 4511 (88.4%) | 0 (0.0%) | 0 (0.0%) | |
| Hispanic | 379 (7.4%) | 1120 (70.0%) | 0 (0.0%) | |
| other | 213 (4.2%) | 481 (30.0%) | 205 (15.2%) | |
| non-Hispanic Black | 0 (0.0%) | 0 (0.0%) | 1143 (84.8%) | |

For continuous variables, the p-value represents an ANOVA t-test with equal variances, while for categorical variables, it is a chi-squared p-value. Abbreviations: INR = income-to-needs; D-PRS = depression polygenic risk score; HS = high-school; SD = standard deviation.

**Supplemental Table 4: Withdrawn/Depressed Symptoms Analyses Demographics for 2-Year Follow-Up Data Collection**

Comparison of Depressed/Withdrawn Sample Across Cohorts - 2-Year Follow-Up

| | European-like (N=3575) | Not European-like Group 1 (N=1062) | Not European-like Group 2 (N=775) | p value |
|---|---|---|---|---|
| **Age (years)** | | | | 0.093 |
| Mean (SD) | 11.927 (0.639) | 11.905 (0.650) | 11.874 (0.617) | |
| Range | 10.583 - 13.667 | 10.667 - 13.583 | 10.583 - 13.500 | |
| **INR** | | | | < 0.001 |
| Mean (SD) | 4.686 (2.741) | 3.369 (2.582) | 2.521 (2.252) | |
| Range | 0.064 - 12.315 | 0.058 - 12.233 | 0.058 - 10.439 | |
| **D-PRS (scaled)** | | | | 0.968 |
| Mean (SD) | -0.011 (0.981) | -0.019 (0.960) | -0.011 (1.002) | |
| Range | -3.038 - 7.263 | -1.701 - 4.382 | -3.700 - 2.338 | |
| **Withdrawn/Depressed Symptom Scale** | | | | < 0.001 |
| N-Miss | 3 | 1 | 1 | |
| Mean (SD) | 1.556 (2.275) | 1.447 (2.225) | 1.221 (2.030) | |
| Range | 0.000 - 19.000 | 0.000 - 13.000 | 0.000 - 15.000 | |
| **Sex at Birth** | | | | 0.104 |
| Female | 1662 (46.5%) | 473 (44.5%) | 384 (49.5%) | |
| Male | 1913 (53.5%) | 589 (55.5%) | 391 (50.5%) | |
| **Highest Caregiver's Education** | | | | < 0.001 |
| N-Miss | 0 | 0 | 1 | |
| Less Than HS | 21 (0.6%) | 98 (9.2%) | 27 (3.5%) | |
| HS Diploma/GED | 123 (3.4%) | 125 (11.8%) | 154 (19.9%) | |
| Some College | 1287 (36.0%) | 480 (45.2%) | 407 (52.6%) | |
| Bachelor | 646 (18.1%) | 132 (12.4%) | 84 (10.9%) | |
| Post-Grad | 1498 (41.9%) | 227 (21.4%) | 102 (13.2%) | |
| **Caregiver-Identified Race/Ethnicity** | | | | < 0.001 |
| non-Hispanic white | 3185 (89.1%) | 33 (3.1%) | 0 (0.0%) | |
| non-Hispanic Black | 1 (0.0%) | 1 (0.1%) | 648 (83.6%) | |
| Hispanic | 249 (7.0%) | 748 (70.4%) | 50 (6.5%) | |
| non-Hispanic Asian | 1 (0.0%) | 33 (3.1%) | 0 (0.0%) | |
| other | 139 (3.9%) | 247 (23.3%) | 77 (9.9%) | |
| **Race/Ethnicity Categories for Modeling** | | | | < 0.001 |
| non-Hispanic white | 3185 (89.1%) | 0 (0.0%) | 0 (0.0%) | |
| Hispanic | 249 (7.0%) | 748 (70.4%) | 0 (0.0%) | |
| other | 141 (3.9%) | 314 (29.6%) | 127 (16.4%) | |
| non-Hispanic Black | 0 (0.0%) | 0 (0.0%) | 648 (83.6%) | |

For continuous variables, the p-value represents an ANOVA t-test with equal variances, while for categorical variables, it is a chi-squared p-value. Abbreviations: INR = income-to-needs; D-PRS = depression polygenic risk score; HS = high-school; SD = standard deviation.

**Supplemental Table 5: Positive Affect Analyses Demographics for 6-Month Follow-Up Data Collection**

Comparison of Positive Affect Sample Across Cohorts - 6-Month Follow-Up

| | European-like (N=5163) | Not European-like Group 1 (N=1638) | Not European-like Group 2 (N=1334) | p value |
|---|---|---|---|---|
| **Age (years)** | | | | 0.798 |
| Mean (SD) | 10.380 (0.627) | 10.389 (0.629) | 10.390 (0.609) | |
| Range | 9.333 - 11.667 | 9.250 - 11.833 | 9.250 - 11.667 | |
| **INR** | | | | < 0.001 |
| Mean (SD) | 4.636 (2.770) | 3.388 (2.592) | 2.464 (2.309) | |
| Range | 0.059 - 12.315 | 0.033 - 12.233 | 0.052 - 12.315 | |
| **Positive Affect Score** | | | | < 0.001 |
| N-Miss | 67 | 22 | 25 | |
| Mean (SD) | 22.184 (3.535) | 21.696 (3.641) | 21.650 (3.877) | |
| Range | 9.000 - 27.000 | 9.000 - 27.000 | 9.000 - 27.000 | |
| **D-PRS (scaled)** | | | | 0.996 |
| Mean (SD) | 0.004 (1.011) | 0.007 (1.008) | 0.005 (0.999) | |
| Range | -3.128 - 8.331 | -1.701 - 4.512 | -3.700 - 2.813 | |
| **Sex at Birth** | | | | 0.007 |
| Female | 2420 (46.9%) | 720 (44.0%) | 664 (49.8%) | |
| Male | 2743 (53.1%) | 918 (56.0%) | 670 (50.2%) | |
| **Highest Caregiver's Education** | | | | < 0.001 |
| N-Miss | 0 | 0 | 1 | |
| Less Than HS | 30 (0.6%) | 152 (9.3%) | 52 (3.9%) | |
| HS Diploma/GED | 208 (4.0%) | 208 (12.7%) | 284 (21.3%) | |
| Some College | 1883 (36.5%) | 724 (44.2%) | 699 (52.4%) | |
| Bachelor | 918 (17.8%) | 201 (12.3%) | 130 (9.8%) | |
| Post-Grad | 2124 (41.1%) | 353 (21.6%) | 168 (12.6%) | |
| **Caregiver-Identified Race/Ethnicity** | | | | < 0.001 |
| non-Hispanic white | 4557 (88.3%) | 48 (2.9%) | 1 (0.1%) | |
| non-Hispanic Black | 2 (0.0%) | 5 (0.3%) | 1132 (84.9%) | |
| Hispanic | 387 (7.5%) | 1149 (70.1%) | 67 (5.0%) | |
| non-Hispanic Asian | 3 (0.1%) | 57 (3.5%) | 0 (0.0%) | |
| other | 214 (4.1%) | 379 (23.1%) | 134 (10.0%) | |
| **Race/Ethnicity Categories for Modeling** | | | | < 0.001 |
| non-Hispanic white | 4557 (88.3%) | 0 (0.0%) | 0 (0.0%) | |
| Hispanic | 387 (7.5%) | 1149 (70.1%) | 0 (0.0%) | |
| other | 219 (4.2%) | 489 (29.9%) | 202 (15.1%) | |
| non-Hispanic Black | 0 (0.0%) | 0 (0.0%) | 1132 (84.9%) | |

For continuous variables, the p-value represents an ANOVA t-test with equal variances, while for categorical variables, it is a chi-squared p-value. Abbreviations: INR, income-to-needs; D-PRS, depression polygenic risk score; HS, high-school.

#### Supplemental Table 6: Positive Affect Analyses Demographics for 1-Year Follow-Up Data Collection

Comparison of Positive Affect Sample Across Cohorts - 1-Year Follow-Up

| | European-like (N=5103) | Not European-like Group 1 (N=1601) | Not European-like Group 2 (N=1348) | p value |
|---|---|---|---|---|
| **Age (years)** | | | | 0.589 |
| Mean (SD) | 10.913 (0.638) | 10.894 (0.638) | 10.906 (0.624) | |
| Range | 9.667 - 12.417 | 9.750 - 12.250 | 9.750 - 12.417 | |
| **INR** | | | | < 0.001 |
| Mean (SD) | 4.642 (2.764) | 3.389 (2.587) | 2.413 (2.272) | |
| Range | 0.059 - 12.315 | 0.064 - 12.233 | 0.052 - 11.989 | |
| **Positive Affect Score** | | | | 0.120 |
| N-Miss | 431 | 171 | 163 | |
| Mean (SD) | 21.531 (3.924) | 21.394 (4.029) | 21.716 (4.119) | |
| Range | 9.000 - 27.000 | 9.000 - 27.000 | 9.000 - 27.000 | |
| **D-PRS (scaled)** | | | | 0.946 |
| Mean (SD) | -0.005 (0.995) | 0.001 (1.010) | 0.004 (0.996) | |
| Range | -3.128 - 8.331 | -1.701 - 4.512 | -3.700 - 2.338 | |
| **Sex at Birth** | | | | 0.007 |
| Female | 2393 (46.9%) | 708 (44.2%) | 675 (50.1%) | |
| Male | 2710 (53.1%) | 893 (55.8%) | 673 (49.9%) | |
| **Highest Caregiver's Education** | | | | < 0.001 |
| N-Miss | 0 | 0 | 1 | |
| Less Than HS | 29 (0.6%) | 143 (8.9%) | 53 (3.9%) | |
| HS Diploma/GED | 195 (3.8%) | 203 (12.7%) | 294 (21.8%) | |
| Some College | 1854 (36.3%) | 715 (44.7%) | 702 (52.1%) | |
| Bachelor | 910 (17.8%) | 192 (12.0%) | 129 (9.6%) | |
| Post-Grad | 2115 (41.4%) | 348 (21.7%) | 169 (12.5%) | |
| **Caregiver-Identified Race/Ethnicity** | | | | < 0.001 |
| non-Hispanic white | 4511 (88.4%) | 48 (3.0%) | 1 (0.1%) | |
| non-Hispanic Black | 2 (0.0%) | 4 (0.2%) | 1143 (84.8%) | |
| Hispanic | 379 (7.4%) | 1120 (70.0%) | 73 (5.4%) | |
| non-Hispanic Asian | 3 (0.1%) | 55 (3.4%) | 0 (0.0%) | |
| other | 208 (4.1%) | 374 (23.4%) | 131 (9.7%) | |
| **Race/Ethnicity Categories for Modeling** | | | | < 0.001 |
| non-Hispanic white | 4511 (88.4%) | 0 (0.0%) | 0 (0.0%) | |
| Hispanic | 379 (7.4%) | 1120 (70.0%) | 0 (0.0%) | |
| other | 213 (4.2%) | 481 (30.0%) | 205 (15.2%) | |
| non-Hispanic Black | 0 (0.0%) | 0 (0.0%) | 1143 (84.8%) | |

For continuous variables, the p-value represents an ANOVA t-test with equal variances, while for categorical variables, it is a chi-squared p-value. Abbreviations: INR, income-to-needs; D-PRS, depression polygenic risk score; HS, high-school.

#### Supplemental Table 7: Positive Affect Analyses Demographics for 18-Month Follow-Up Data Collection

Comparison of Positive Affect Sample Across Cohorts - 18-Month Follow-Up

| | European-like (N=4964) | Not European-like Group 1 (N=1531) | Not European-like Group 2 (N=1184) | p value |
|---|---|---|---|---|
| **Age (years)** | | | | 0.980 |
| Mean (SD) | 11.379 (0.631) | 11.379 (0.635) | 11.383 (0.618) | |
| Range | 10.000 - 12.833 | 10.250 - 12.750 | 10.250 - 12.833 | |
| **INR** | | | | < 0.001 |
| Mean (SD) | 4.655 (2.770) | 3.466 (2.605) | 2.536 (2.342) | |
| Range | 0.064 - 12.315 | 0.064 - 12.233 | 0.052 - 12.315 | |
| **Positive Affect Score** | | | | < 0.001 |
| N-Miss | 43 | 26 | 18 | |
| Mean (SD) | 22.474 (3.592) | 21.888 (3.681) | 21.823 (3.973) | |
| Range | 9.000 - 27.000 | 9.000 - 27.000 | 9.000 - 27.000 | |
| **D-PRS (scaled)** | | | | 0.773 |
| Mean (SD) | -0.002 (0.992) | -0.010 (0.996) | -0.025 (1.014) | |
| Range | -3.128 - 8.331 | -1.701 - 4.382 | -3.700 - 2.338 | |
| **Sex at Birth** | | | | 0.060 |
| Female | 2317 (46.7%) | 685 (44.7%) | 584 (49.3%) | |
| Male | 2647 (53.3%) | 846 (55.3%) | 600 (50.7%) | |
| **Highest Caregiver's Education** | | | | < 0.001 |
| N-Miss | 0 | 0 | 1 | |
| Less Than HS | 26 (0.5%) | 129 (8.4%) | 42 (3.6%) | |
| HS Diploma/GED | 183 (3.7%) | 178 (11.6%) | 244 (20.6%) | |
| Some College | 1785 (36.0%) | 689 (45.0%) | 612 (51.7%) | |
| Bachelor | 900 (18.1%) | 194 (12.7%) | 127 (10.7%) | |
| Post-Grad | 2070 (41.7%) | 341 (22.3%) | 158 (13.4%) | |
| **Caregiver-Identified Race/Ethnicity** | | | | < 0.001 |
| non-Hispanic white | 4391 (88.5%) | 45 (2.9%) | 1 (0.1%) | |
| non-Hispanic Black | 1 (0.0%) | 3 (0.2%) | 994 (84.0%) | |
| Hispanic | 367 (7.4%) | 1073 (70.1%) | 68 (5.7%) | |
| non-Hispanic Asian | 3 (0.1%) | 54 (3.5%) | 0 (0.0%) | |
| other | 202 (4.1%) | 356 (23.3%) | 121 (10.2%) | |
| **Race/Ethnicity Categories for Modeling** | | | | < 0.001 |
| non-Hispanic white | 4391 (88.5%) | 0 (0.0%) | 0 (0.0%) | |
| Hispanic | 367 (7.4%) | 1073 (70.1%) | 0 (0.0%) | |
| other | 206 (4.1%) | 458 (29.9%) | 190 (16.0%) | |
| non-Hispanic Black | 0 (0.0%) | 0 (0.0%) | 994 (84.0%) | |

For continuous variables, the p-value represents an ANOVA t-test with equal variances, while for categorical variables, it is a chi-squared p-value. Abbreviations: INR, income-to-needs; D-PRS, depression polygenic risk score; HS, high-school.

**Supplemental Table 8: Resting-State Functional MRI Analyses Demographics for Baseline Data Collection**

Comparison of rsfMRI Sample Across Cohorts - Baseline

| | European-like (N=4292) | Not European-like Group 1 (N=1373) | Not European-like Group 2 (N=1068) | p value |
|---|---|---|---|---|
| **Age (years)** | | | | 0.773 |
| Mean (SD) | 9.934 (0.622) | 9.924 (0.626) | 9.922 (0.599) | |
| Range | 8.917 - 11.000 | 8.917 - 11.083 | 8.917 - 11.000 | |
| **INR** | | | | < 0.001 |
| Mean (SD) | 4.649 (2.760) | 3.425 (2.609) | 2.436 (2.302) | |
| Range | 0.064 - 12.315 | 0.033 - 12.151 | 0.052 - 12.315 | |
| **D-PRS (scaled)** | | | | 0.978 |
| Mean (SD) | 0.008 (1.016) | 0.012 (1.011) | 0.015 (0.990) | |
| Range | -3.082 - 7.115 | -1.691 - 4.460 | -3.623 - 3.240 | |
| **Sex at Birth** | | | | 0.091 |
| F | 2075 (48.3%) | 646 (47.1%) | 549 (51.4%) | |
| M | 2217 (51.7%) | 727 (52.9%) | 519 (48.6%) | |
| **Highest Caregiver's Education** | | | | < 0.001 |
| N-Miss | 0 | 0 | 1 | |
| lessthan_HS | 15 (0.3%) | 121 (8.8%) | 47 (4.4%) | |
| HSDiploma_GED | 163 (3.8%) | 182 (13.3%) | 200 (18.7%) | |
| SomeCollege | 1548 (36.1%) | 608 (44.3%) | 583 (54.6%) | |
| Bachelor | 762 (17.8%) | 174 (12.7%) | 107 (10.0%) | |
| PostGradDegree | 1804 (42.0%) | 288 (21.0%) | 130 (12.2%) | |
| **Caregiver-Identified Race/Ethnicity** | | | | < 0.001 |
| White | 3802 (88.6%) | 38 (2.8%) | 1 (0.1%) | |
| Black | 2 (0.0%) | 4 (0.3%) | 904 (84.6%) | |
| Hispanic | 319 (7.4%) | 975 (71.0%) | 60 (5.6%) | |
| Asian | 2 (0.0%) | 41 (3.0%) | 0 (0.0%) | |
| Other | 167 (3.9%) | 315 (22.9%) | 103 (9.6%) | |
| **Race/Ethnicity Categories for Modeling** | | | | < 0.001 |
| White | 3802 (88.6%) | 0 (0.0%) | 0 (0.0%) | |
| Hispanic | 319 (7.4%) | 975 (71.0%) | 0 (0.0%) | |
| Other | 171 (4.0%) | 398 (29.0%) | 164 (15.4%) | |
| Black | 0 (0.0%) | 0 (0.0%) | 904 (84.6%) | |

For continuous variables, the p-value represents an ANOVA t-test with equal variances, while for categorical variables, it is a chi-squared p-value. Abbreviations: INR = income-to-needs; D-PRS = depression polygenic risk score; HS = high-school; SD = standard deviation.

**Supplemental Table 9: Resting-State Functional MRI Analyses Demographics for 2-Year Follow-Up Data Collection**

Comparison of rsfMRI Sample Across Cohorts - 2-Year Follow-Up

| | European-like (N=2666) | Not European-like Group 1 (N=767) | Not European-like Group 2 (N=481) | p value |
|---|---|---|---|---|
| **Age (years)** | | | | 0.127 |
| Mean (SD) | 11.976 (0.645) | 11.952 (0.663) | 11.913 (0.626) | |
| Range | 10.583 - 13.667 | 10.750 - 13.583 | 10.583 - 13.250 | |
| **INR** | | | | < 0.001 |
| Mean (SD) | 4.711 (2.722) | 3.407 (2.551) | 2.577 (2.301) | |
| Range | 0.064 - 12.315 | 0.058 - 11.276 | 0.059 - 10.439 | |
| **D-PRS (scaled)** | | | | 0.875 |
| Mean (SD) | -0.010 (0.969) | -0.010 (0.973) | 0.014 (1.014) | |
| Range | -3.082 - 6.978 | -1.691 - 4.331 | -3.130 - 3.240 | |
| **Sex at Birth** | | | | 0.514 |
| F | 1272 (47.7%) | 366 (47.7%) | 243 (50.5%) | |
| M | 1394 (52.3%) | 401 (52.3%) | 238 (49.5%) | |
| **Highest Caregiver's Education** | | | | < 0.001 |
| N-Miss | 0 | 0 | 1 | |
| lessthan_HS | 10 (0.4%) | 62 (8.1%) | 16 (3.3%) | |
| HSDiploma_GED | 81 (3.0%) | 91 (11.9%) | 83 (17.3%) | |
| SomeCollege | 931 (34.9%) | 351 (45.8%) | 258 (53.8%) | |
| Bachelor | 490 (18.4%) | 98 (12.8%) | 56 (11.7%) | |
| PostGradDegree | 1154 (43.3%) | 165 (21.5%) | 67 (14.0%) | |
| **Caregiver-Identified Race/Ethnicity** | | | | < 0.001 |
| White | 2385 (89.5%) | 22 (2.9%) | 0 (0.0%) | |
| Black | 1 (0.0%) | 1 (0.1%) | 402 (83.6%) | |
| Hispanic | 185 (6.9%) | 541 (70.5%) | 32 (6.7%) | |
| Asian | 0 (0.0%) | 23 (3.0%) | 0 (0.0%) | |
| Other | 95 (3.6%) | 180 (23.5%) | 47 (9.8%) | |
| **Race/Ethnicity Categories for Modeling** | | | | < 0.001 |
| White | 2385 (89.5%) | 0 (0.0%) | 0 (0.0%) | |
| Hispanic | 185 (6.9%) | 541 (70.5%) | 0 (0.0%) | |
| Other | 96 (3.6%) | 226 (29.5%) | 79 (16.4%) | |
| Black | 0 (0.0%) | 0 (0.0%) | 402 (83.6%) | |

For continuous variables, the p-value represents an ANOVA t-test with equal variances, while for categorical variables, it is a chi-squared p-value. Abbreviations: INR = income-to-needs; D-PRS = depression polygenic risk score; HS = high-school; SD = standard deviation.

**Supplemental Table 10: Structural MRI Analyses Demographics for Baseline Data Collection**

Comparison of sMRI_cortex Sample Across Cohorts - Baseline

| | European-like (N=4931) | Not European-like Group 1 (N=1605) | Not European-like Group 2 (N=1354) | p value |
|---|---|---|---|---|
| **Age (years)** | | | | 0.485 |
| Mean (SD) | 9.912 (0.622) | 9.892 (0.623) | 9.898 (0.600) | |
| Range | 8.917 - 11.000 | 8.917 - 11.083 | 8.917 - 11.000 | |
| **INR** | | | | < 0.001 |
| Mean (SD) | 4.627 (2.767) | 3.399 (2.613) | 2.359 (2.254) | |
| Range | 0.059 - 12.315 | 0.033 - 12.233 | 0.052 - 12.315 | |
| **D-PRS (scaled)** | | | | 0.978 |
| Mean (SD) | 0.010 (1.017) | 0.013 (1.012) | 0.017 (0.991) | |
| Range | -3.049 - 8.362 | -1.705 - 4.462 | -3.687 - 3.216 | |
| **Sex at Birth** | | | | 0.033 |
| F | 2339 (47.4%) | 723 (45.0%) | 675 (49.9%) | |
| M | 2592 (52.6%) | 882 (55.0%) | 679 (50.1%) | |
| **Highest Caregiver's Education** | | | | < 0.001 |
| N-Miss | 0 | 0 | 1 | |
| lessthan_HS | 28 (0.6%) | 144 (9.0%) | 58 (4.3%) | |
| HSDiploma_GED | 195 (4.0%) | 207 (12.9%) | 298 (22.0%) | |
| SomeCollege | 1809 (36.7%) | 725 (45.2%) | 712 (52.6%) | |
| Bachelor | 868 (17.6%) | 197 (12.3%) | 129 (9.5%) | |
| PostGradDegree | 2031 (41.2%) | 332 (20.7%) | 156 (11.5%) | |
| **Caregiver-Identified Race/Ethnicity** | | | | < 0.001 |
| White | 4362 (88.5%) | 50 (3.1%) | 0 (0.0%) | |
| Black | 3 (0.1%) | 4 (0.2%) | 1147 (84.7%) | |
| Hispanic | 373 (7.6%) | 1135 (70.7%) | 73 (5.4%) | |
| Asian | 2 (0.0%) | 48 (3.0%) | 0 (0.0%) | |
| Other | 191 (3.9%) | 368 (22.9%) | 134 (9.9%) | |
| **Race/Ethnicity Categories for Modeling** | | | | < 0.001 |
| White | 4362 (88.5%) | 0 (0.0%) | 0 (0.0%) | |
| Hispanic | 373 (7.6%) | 1135 (70.7%) | 0 (0.0%) | |
| Other | 196 (4.0%) | 470 (29.3%) | 207 (15.3%) | |
| Black | 0 (0.0%) | 0 (0.0%) | 1147 (84.7%) | |

For continuous variables, the p-value represents an ANOVA t-test with equal variances, while for categorical variables, it is a chi-squared p-value. Abbreviations: INR = income-to-needs; D-PRS = depression polygenic risk score; HS = high-school; SD = standard deviation.

#### Supplemental Table 11: Structural MRI Analyses Demographics for 2-Year Follow-Up Data Collection

Comparison of sMRI_cortex Sample Across Cohorts - 2-Year Follow-Up

| | European-like (N=3261) | Not European-like Group 1 (N=963) | Not European-like Group 2 (N=703) | p value |
|---|---|---|---|---|
| **Age (years)** | | | | 0.016 |
| Mean (SD) | 11.942 (0.640) | 11.912 (0.650) | 11.869 (0.610) | |
| Range | 10.583 - 13.667 | 10.667 - 13.583 | 10.583 - 13.500 | |
| **INR** | | | | < 0.001 |
| Mean (SD) | 4.687 (2.731) | 3.398 (2.592) | 2.514 (2.250) | |
| Range | 0.064 - 12.315 | 0.058 - 12.233 | 0.058 - 10.439 | |
| **D-PRS (scaled)** | | | | 0.956 |
| Mean (SD) | -0.018 (0.967) | -0.016 (0.964) | -0.005 (1.009) | |
| Range | -3.049 - 7.290 | -1.705 - 4.333 | -3.687 - 3.216 | |
| **Sex at Birth** | | | | 0.168 |
| F | 1522 (46.7%) | 429 (44.5%) | 346 (49.2%) | |
| M | 1739 (53.3%) | 534 (55.5%) | 357 (50.8%) | |
| **Highest Caregiver's Education** | | | | < 0.001 |
| N-Miss | 0 | 0 | 1 | |
| lessthan_HS | 19 (0.6%) | 85 (8.8%) | 23 (3.3%) | |
| HSDiploma_GED | 109 (3.3%) | 112 (11.6%) | 141 (20.1%) | |
| SomeCollege | 1166 (35.8%) | 442 (45.9%) | 370 (52.7%) | |
| Bachelor | 597 (18.3%) | 119 (12.4%) | 78 (11.1%) | |
| PostGradDegree | 1370 (42.0%) | 205 (21.3%) | 90 (12.8%) | |
| **Caregiver-Identified Race/Ethnicity** | | | | < 0.001 |
| White | 2922 (89.6%) | 30 (3.1%) | 0 (0.0%) | |
| Black | 1 (0.0%) | 1 (0.1%) | 581 (82.6%) | |
| Hispanic | 222 (6.8%) | 680 (70.6%) | 48 (6.8%) | |
| Asian | 0 (0.0%) | 27 (2.8%) | 0 (0.0%) | |
| Other | 116 (3.6%) | 225 (23.4%) | 74 (10.5%) | |
| **Race/Ethnicity Categories for Modeling** | | | | < 0.001 |
| White | 2922 (89.6%) | 0 (0.0%) | 0 (0.0%) | |
| Hispanic | 222 (6.8%) | 680 (70.6%) | 0 (0.0%) | |
| Other | 117 (3.6%) | 283 (29.4%) | 122 (17.4%) | |
| Black | 0 (0.0%) | 0 (0.0%) | 581 (82.6%) | |

For continuous variables, the p-value represents an ANOVA t-test with equal variances, while for categorical variables, it is a chi-squared p-value. Abbreviations: INR = income-to-needs; D-PRS = depression polygenic risk score; HS = high-school; SD = standard deviation.

#### Supplemental Table 12: Predictors, Covariates, and Random Effects by Outcome in Each Sample

| | | European-like Sample | | | Not European-like Group 1 Sample | | | Not European-like Group 2 Sample | | |
|---|---|---|---|---|---|---|---|---|---|---|
| | *Outcome* | *Predictors* | *Covariates* | *Random Effects* | *Predictors* | *Covariates* | *Random Effects* | *Predictors* | *Covariates* | *Random Effects* |
| **Behavior Outcomes** | Withdrawn/Depressed Syndrome Scale | INR + D-PRS | age, sex-at-birth, highest caregivers education, caregiver-identified race/ethnicity, ABCD Study site, and first 10 genetics PCs | subjects nested within site | Age + INR + INR*Age + D-PRS | sex-at-birth, highest caregivers education, caregiver-identified race/ethnicity, ABCD Study site, and first 10 genetics PCs | subjects nested within site | Age + INR + INR*Age + D-PRS | sex-at-birth, highest caregivers education, caregiver-identified race/ethnicity, ABCD Study site, and first 10 genetics PCs | subjects nested within site |
| | Positive Affect | INR + D-PRS | age, sex-at-birth, highest caregivers education, caregiver-identified race/ethnicity, ABCD Study site, and the first 10 genetics PCs | subjects nested within site | INR + D-PRS | age, sex-at-birth, highest caregivers education, caregiver-identified race/ethnicity, ABCD Study site, and the first 10 genetics PCs | subjects nested within site | INR + D-PRS | age, sex-at-birth, highest caregivers education, caregiver-identified race/ethnicity, ABCD Study site, and the first 10 genetics PCs | subjects nested within site |
| | DMN Intra-Network Connectivity | INR + D-PRS<br><br>Trending effect (pFDR<0.10) seen for INR-by-PRS for this outcome, so this interaction was also examined | age, sex-at-birth, highest caregivers education, caregiver-identified race/ethnicity, handedness score, MRI motion, MRI device serial number, and the first 10 genetics PCs | subjects nested within site | INR + D-PRS | age, sex-at-birth, highest caregivers education, caregiver-identified race/ethnicity, handedness score, MRI motion, MRI device serial number, and the first 10 genetics PCs | subjects nested within site | INR | age, sex-at-birth, highest caregivers education, caregiver-identified race/ethnicity, handedness score, MRI motion, and MRI device serial number | subjects nested within site |
| | FPN Intra-Network Connectivity | INR + D-PRS<br><br>Trending effect (pFDR<0.10) seen for INR-by-PRS for this outcome, so this interaction was also examined | age, sex-at-birth, highest caregivers education, caregiver-identified race/ethnicity, handedness score, MRI motion, MRI device serial number, and the first 10 genetics PCs | subjects nested within site | INR + D-PRS | age, sex-at-birth, highest caregivers education, caregiver-identified race/ethnicity, handedness score, MRI motion, MRI device serial number, and the first 10 genetics PCs | subjects nested within site | INR | age, sex-at-birth, highest caregivers education, caregiver-identified race/ethnicity, handedness score, MRI motion, and MRI device serial number | subjects nested within site |
| | SN Intra-Network Connectivity | INR + D-PRS | age, sex-at-birth, highest caregivers education, caregiver-identified race/ethnicity, handedness score, MRI motion, MRI device serial number, and the first 10 genetics PCs | subjects nested within site | INR + D-PRS | age, sex-at-birth, highest caregivers education, caregiver-identified race/ethnicity, handedness score, MRI motion, MRI device serial number, and the first 10 genetics PCs | subjects nested within site | INR | age, sex-at-birth, highest caregivers education, caregiver-identified race/ethnicity, handedness score, MRI motion, and MRI device serial number | subjects nested within site |
| | DMN to FPN Inter-Network Connectivity | INR + D-PRS | age, sex-at-birth, highest caregivers education, caregiver-identified race/ethnicity, handedness score, MRI motion, MRI device serial number, and the first 10 genetics PCs | subjects nested within site | INR + D-PRS | age, sex-at-birth, highest caregivers education, caregiver-identified race/ethnicity, handedness score, MRI motion, MRI device serial number, and the first 10 genetics PCs | subjects nested within site | INR | age, sex-at-birth, highest caregivers education, caregiver-identified race/ethnicity, handedness score, MRI motion, and MRI device serial number | subjects nested within site |
| | DMN to SN Inter-Network Connectivity | INR + D-PRS | age, sex-at-birth, highest caregivers education, caregiver-identified race/ethnicity, handedness score, MRI motion, MRI device serial number, and the first 10 genetics PCs | subjects nested within site | INR + D-PRS | age, sex-at-birth, highest caregivers education, caregiver-identified race/ethnicity, handedness score, MRI motion, MRI device serial number, and the first 10 genetics PCs | subjects nested within site | INR | age, sex-at-birth, highest caregivers education, caregiver-identified race/ethnicity, handedness score, MRI motion, and MRI device serial number | subjects nested within site |
| | FPN to SN Inter-Network Connectivity | INR + D-PRS | age, sex-at-birth, highest caregivers education, caregiver-identified race/ethnicity, handedness score, MRI motion, MRI device serial number, and the first 10 genetics PCs | subjects nested within site | INR + D-PRS | age, sex-at-birth, highest caregivers education, caregiver-identified race/ethnicity, handedness score, MRI motion, MRI device serial number, and the first 10 genetics PCs | subjects nested within site | INR | age, sex-at-birth, highest caregivers education, caregiver-identified race/ethnicity, handedness score, MRI motion, and MRI device serial number | subjects nested within site |
| | DMN to Left Amygdala Connectivity | INR + D-PRS | age, sex-at-birth, highest caregivers education, caregiver-identified race/ethnicity, handedness score, MRI motion, MRI device serial number, and the first 10 genetics PCs | subjects nested within site | INR + D-PRS | age, sex-at-birth, highest caregivers education, caregiver-identified race/ethnicity, handedness score, MRI motion, MRI device serial number, and the first 10 genetics PCs | subjects nested within site | INR | age, sex-at-birth, highest caregivers education, caregiver-identified race/ethnicity, handedness score, MRI motion, and MRI device serial number | subjects nested within site |

| | | | | | | | | | |
|---|---|---|---|---|---|---|---|---|---|
| **Functional Connectivity Outcomes** | DMN to Right Amygdala Connectivity | INR + D-PRS | age, sex-at-birth, highest caregivers education, caregiver-identified race/ethnicity, handedness score, MRI motion, MRI device serial number, and the first 10 genetics PCs | subjects nested within site | INR + D-PRS | age, sex-at-birth, highest caregivers education, caregiver-identified race/ethnicity, handedness score, MRI motion, MRI device serial number, and the first 10 genetics PCs | subjects nested within site | INR | age, sex-at-birth, highest caregivers education, caregiver-identified race/ethnicity, handedness score, MRI motion, and MRI device serial number | subjects nested within site |
| | DMN to Left Hippocampus Connectivity | INR + D-PRS | age, sex-at-birth, highest caregivers education, caregiver-identified race/ethnicity, handedness score, MRI motion, MRI device serial number, and the first 10 genetics PCs | subjects nested within site | INR + D-PRS | age, sex-at-birth, highest caregivers education, caregiver-identified race/ethnicity, handedness score, MRI motion, MRI device serial number, and the first 10 genetics PCs | subjects nested within site | INR | age, sex-at-birth, highest caregivers education, caregiver-identified race/ethnicity, handedness score, MRI motion, and MRI device serial number | subjects nested within site |
| | DMN to Right Hippocampus Connectivity | INR + D-PRS | age, sex-at-birth, highest caregivers education, caregiver-identified race/ethnicity, handedness score, MRI motion, MRI device serial number, and the first 10 genetics PCs | subjects nested within site | INR + D-PRS | age, sex-at-birth, highest caregivers education, caregiver-identified race/ethnicity, handedness score, MRI motion, MRI device serial number, and the first 10 genetics PCs | subjects nested within site | INR | age, sex-at-birth, highest caregivers education, caregiver-identified race/ethnicity, handedness score, MRI motion, and MRI device serial number | subjects nested within site |
| | FPN to Left Amygdala Connectivity | INR + D-PRS | age, sex-at-birth, highest caregivers education, caregiver-identified race/ethnicity, handedness score, MRI motion, MRI device serial number, and the first 10 genetics PCs | subjects nested within site | INR + D-PRS | age, sex-at-birth, highest caregivers education, caregiver-identified race/ethnicity, handedness score, MRI motion, MRI device serial number, and the first 10 genetics PCs | subjects nested within site | INR | age, sex-at-birth, highest caregivers education, caregiver-identified race/ethnicity, handedness score, MRI motion, and MRI device serial number | subjects nested within site |
| | FPN to Right Amygdala Connectivity | INR + D-PRS | age, sex-at-birth, highest caregivers education, caregiver-identified race/ethnicity, handedness score, MRI motion, MRI device serial number, and the first 10 genetics PCs | subjects nested within site | INR + D-PRS | age, sex-at-birth, highest caregivers education, caregiver-identified race/ethnicity, handedness score, MRI motion, MRI device serial number, and the first 10 genetics PCs | subjects nested within site | INR | age, sex-at-birth, highest caregivers education, caregiver-identified race/ethnicity, handedness score, MRI motion, and MRI device serial number | subjects nested within site |
| | FPN to Left Hippocampus Connectivity | INR + D-PRS | age, sex-at-birth, highest caregivers education, caregiver-identified race/ethnicity, handedness score, MRI motion, MRI device serial number, and the first 10 genetics PCs | subjects nested within site | INR + D-PRS | age, sex-at-birth, highest caregivers education, caregiver-identified race/ethnicity, handedness score, MRI motion, MRI device serial number, and the first 10 genetics PCs | subjects nested within site | INR | age, sex-at-birth, highest caregivers education, caregiver-identified race/ethnicity, handedness score, MRI motion, and MRI device serial number | subjects nested within site |
| | FPN to Right Hippocampus Connectivity | INR + D-PRS | age, sex-at-birth, highest caregivers education, caregiver-identified race/ethnicity, handedness score, MRI motion, MRI device serial number, and the first 10 genetics PCs | subjects nested within site | INR + D-PRS | age, sex-at-birth, highest caregivers education, caregiver-identified race/ethnicity, handedness score, MRI motion, MRI device serial number, and the first 10 genetics PCs | subjects nested within site | INR | age, sex-at-birth, highest caregivers education, caregiver-identified race/ethnicity, handedness score, MRI motion, and MRI device serial number | subjects nested within site |
| | SN to Left Amygdala Connectivity | INR + D-PRS | age, sex-at-birth, highest caregivers education, caregiver-identified race/ethnicity, handedness score, MRI motion, MRI device serial number, and the first 10 genetics PCs | subjects nested within site | INR + D-PRS | age, sex-at-birth, highest caregivers education, caregiver-identified race/ethnicity, handedness score, MRI motion, MRI device serial number, and the first 10 genetics PCs | subjects nested within site | INR | age, sex-at-birth, highest caregivers education, caregiver-identified race/ethnicity, handedness score, MRI motion, and MRI device serial number | subjects nested within site |
| | SN to Right Amygdala Connectivity | INR + D-PRS | age, sex-at-birth, highest caregivers education, caregiver-identified race/ethnicity, handedness score, MRI motion, MRI device serial number, and the first 10 genetics PCs | subjects nested within site | INR + D-PRS | age, sex-at-birth, highest caregivers education, caregiver-identified race/ethnicity, handedness score, MRI motion, MRI device serial number, and the first 10 genetics PCs | subjects nested within site | INR | age, sex-at-birth, highest caregivers education, caregiver-identified race/ethnicity, handedness score, MRI motion, and MRI device serial number | subjects nested within site |
| | SN to Left Hippocampus Connectivity | INR + D-PRS | age, sex-at-birth, highest caregivers education, caregiver-identified race/ethnicity, handedness score, MRI motion, MRI device serial number, and the first 10 genetics PCs | subjects nested within site | INR + D-PRS | age, sex-at-birth, highest caregivers education, caregiver-identified race/ethnicity, handedness score, MRI motion, MRI device serial number, and the first 10 genetics PCs | subjects nested within site | INR | age, sex-at-birth, highest caregivers education, caregiver-identified race/ethnicity, handedness score, MRI motion, and MRI device serial number | subjects nested within site |

| | | | | | | | | | | |
|---|---|---|---|---|---|---|---|---|---|---|
| | SN to Right Hippocampus Connectivity | INR + D-PRS | age, sex-at-birth, highest caregivers education, caregiver-identified race/ethnicity, handedness score, MRI motion, MRI device serial number, and the first 10 genetics PCs | subjects nested within site | INR + D-PRS | age, sex-at-birth, highest caregivers education, caregiver-identified race/ethnicity, handedness score, MRI motion, MRI device serial number, and the first 10 genetics PCs | subjects nested within site | INR | age, sex-at-birth, highest caregivers education, caregiver-identified race/ethnicity, handedness score, MRI motion, and MRI device serial number | subjects nested within site |
| | Transverse frontopolar gyri and sulci | INR + D-PRS | age, sex-at-birth, highest caregivers education, caregiver-identified race/ethnicity, handedness score, MRI device serial number, and the first 10 genetics PCs | hemisphere nested within subject nested within site | INR + D-PRS | age, sex-at-birth, highest caregivers education, caregiver-identified race/ethnicity, handedness score, MRI device serial number, and the first 10 genetics PCs | hemisphere nested within subject nested within site | Age + INR + Ag | sex-at-birth, highest caregivers education, caregiver-identified race/ethnicity, handedness score, and MRI device serial number | hemisphere nested within subject nested within site |
| | Orbital gyri | INR + D-PRS | age, sex-at-birth, highest caregivers education, caregiver-identified race/ethnicity, handedness score, MRI device serial number, and the first 10 genetics PCs | hemisphere nested within subject nested within site | INR + D-PRS | age, sex-at-birth, highest caregivers education, caregiver-identified race/ethnicity, handedness score, MRI device serial number, and the first 10 genetics PCs | hemisphere nested within subject nested within site | INR | age, sex-at-birth, highest caregivers education, caregiver-identified race/ethnicity, handedness score, and MRI device serial number | hemisphere nested within subject nested within site |
| | Supramarginal gyrus | INR + D-PRS | age, sex-at-birth, highest caregivers education, caregiver-identified race/ethnicity, handedness score, MRI device serial number, and the first 10 genetics PCs | hemisphere nested within subject nested within site | INR + D-PRS | age, sex-at-birth, highest caregivers education, caregiver-identified race/ethnicity, handedness score, MRI device serial number, and the first 10 genetics PCs | hemisphere nested within subject nested within site | INR | age, sex-at-birth, highest caregivers education, caregiver-identified race/ethnicity, handedness score, and MRI device serial number | hemisphere nested within subject nested within site |
| | Inferior frontal sulcus | INR + D-PRS | age, sex-at-birth, highest caregivers education, caregiver-identified race/ethnicity, handedness score, MRI device serial number, and the first 10 genetics PCs | hemisphere nested within subject nested within site | INR + D-PRS | age, sex-at-birth, highest caregivers education, caregiver-identified race/ethnicity, handedness score, MRI device serial number, and the first 10 genetics PCs | hemisphere nested within subject nested within site | INR | age, sex-at-birth, highest caregivers education, caregiver-identified race/ethnicity, handedness score, and MRI device serial number | hemisphere nested within subject nested within site |
| | Middle frontal sulcus | INR + D-PRS | age, sex-at-birth, highest caregivers education, caregiver-identified race/ethnicity, handedness score, MRI device serial number, and the first 10 genetics PCs | hemisphere nested within subject nested within site | INR + D-PRS | age, sex-at-birth, highest caregivers education, caregiver-identified race/ethnicity, handedness score, MRI device serial number, and the first 10 genetics PCs | hemisphere nested within subject nested within site | INR | age, sex-at-birth, highest caregivers education, caregiver-identified race/ethnicity, handedness score, and MRI device serial number | hemisphere nested within subject nested within site |
| | Intraparietal sulcus and transverse parietal sulci | INR + D-PRS | age, sex-at-birth, highest caregivers education, caregiver-identified race/ethnicity, handedness score, MRI device serial number, and the first 10 genetics PCs | hemisphere nested within subject nested within site | INR + D-PRS | age, sex-at-birth, highest caregivers education, caregiver-identified race/ethnicity, handedness score, MRI device serial number, and the first 10 genetics PCs | hemisphere nested within subject nested within site | INR | age, sex-at-birth, highest caregivers education, caregiver-identified race/ethnicity, handedness score, and MRI device serial number | hemisphere nested within subject nested within site |
| | Inferior part of the precentral sulcus | INR + D-PRS | age, sex-at-birth, highest caregivers education, caregiver-identified race/ethnicity, handedness score, MRI device serial number, and the first 10 genetics PCs | hemisphere nested within subject nested within site | INR + D-PRS | age, sex-at-birth, highest caregivers education, caregiver-identified race/ethnicity, handedness score, MRI device serial number, and the first 10 genetics PCs | hemisphere nested within subject nested within site | INR | age, sex-at-birth, highest caregivers education, caregiver-identified race/ethnicity, handedness score, and MRI device serial number | hemisphere nested within subject nested within site |
| | Inferior temporal sulcus | INR + D-PRS | age, sex-at-birth, highest caregivers education, caregiver-identified race/ethnicity, handedness score, MRI device serial number, and the first 10 genetics PCs | hemisphere nested within subject nested within site | INR + D-PRS | age, sex-at-birth, highest caregivers education, caregiver-identified race/ethnicity, handedness score, MRI device serial number, and the first 10 genetics PCs | hemisphere nested within subject nested within site | INR | age, sex-at-birth, highest caregivers education, caregiver-identified race/ethnicity, handedness score, and MRI device serial number | hemisphere nested within subject nested within site |
| | Fronto-marginal gyrus and sulcus | INR + D-PRS | age, sex-at-birth, highest caregivers education, caregiver-identified race/ethnicity, handedness score, MRI device serial number, and the first 10 genetics PCs | hemisphere nested within subject nested within site | INR + D-PRS | age, sex-at-birth, highest caregivers education, caregiver-identified race/ethnicity, handedness score, MRI device serial number, and the first 10 genetics PCs | hemisphere nested within subject nested within site | INR | age, sex-at-birth, highest caregivers education, caregiver-identified race/ethnicity, handedness score, and MRI device serial number | hemisphere nested within subject nested within site |

| | | | | | | | | | | |
|---|---|---|---|---|---|---|---|---|---|---|
| **Structural Cortical Thickness Outcomes** | Middle frontal gyrus | INR + D-PRS | age, sex-at-birth, highest caregivers education, caregiver-identified race/ethnicity, handedness score, MRI device serial number, and the first 10 genetics PCs | hemisphere nested within subject nested within site | INR + D-PRS | age, sex-at-birth, highest caregivers education, caregiver-identified race/ethnicity, handedness score, MRI device serial number, and the first 10 genetics PCs | hemisphere nested within subject nested within site | Age + INR + Ag | sex-at-birth, highest caregivers education, caregiver-identified race/ethnicity, handedness score, and MRI device serial number | hemisphere nested within subject nested within site |
| | Superior frontal gyrus | INR + D-PRS | age, sex-at-birth, highest caregivers education, caregiver-identified race/ethnicity, handedness score, MRI device serial number, and the first 10 genetics PCs | hemisphere nested within subject nested within site | INR + D-PRS | age, sex-at-birth, highest caregivers education, caregiver-identified race/ethnicity, handedness score, MRI device serial number, and the first 10 genetics PCs | hemisphere nested within subject nested within site | Age + INR + Ag | sex-at-birth, highest caregivers education, caregiver-identified race/ethnicity, handedness score, and MRI device serial number | hemisphere nested within subject nested within site |
| | Middle-posterior part of the cingulate gyrus and sulcus | INR + D-PRS | age, sex-at-birth, highest caregivers education, caregiver-identified race/ethnicity, handedness score, MRI device serial number, and the first 10 genetics PCs | hemisphere nested within subject nested within site | INR + D-PRS | age, sex-at-birth, highest caregivers education, caregiver-identified race/ethnicity, handedness score, MRI device serial number, and the first 10 genetics PCs | hemisphere nested within subject nested within site | INR | age, sex-at-birth, highest caregivers education, caregiver-identified race/ethnicity, handedness score, and MRI device serial number | hemisphere nested within subject nested within site |
| | Angular gyrus | INR + D-PRS | age, sex-at-birth, highest caregivers education, caregiver-identified race/ethnicity, handedness score, MRI device serial number, and the first 10 genetics PCs | hemisphere nested within subject nested within site | INR + D-PRS | age, sex-at-birth, highest caregivers education, caregiver-identified race/ethnicity, handedness score, MRI device serial number, and the first 10 genetics PCs | hemisphere nested within subject nested within site | INR | age, sex-at-birth, highest caregivers education, caregiver-identified race/ethnicity, handedness score, and MRI device serial number | hemisphere nested within subject nested within site |
| | Precuneus | INR + D-PRS | age, sex-at-birth, highest caregivers education, caregiver-identified race/ethnicity, handedness score, MRI device serial number, and the first 10 genetics PCs | hemisphere nested within subject nested within site | INR + D-PRS | age, sex-at-birth, highest caregivers education, caregiver-identified race/ethnicity, handedness score, MRI device serial number, and the first 10 genetics PCs | hemisphere nested within subject nested within site | INR | age, sex-at-birth, highest caregivers education, caregiver-identified race/ethnicity, handedness score, and MRI device serial number | hemisphere nested within subject nested within site |
| | Gyrus rectus | INR + D-PRS | age, sex-at-birth, highest caregivers education, caregiver-identified race/ethnicity, handedness score, MRI device serial number, and the first 10 genetics PCs | hemisphere nested within subject nested within site | INR + D-PRS | age, sex-at-birth, highest caregivers education, caregiver-identified race/ethnicity, handedness score, MRI device serial number, and the first 10 genetics PCs | hemisphere nested within subject nested within site | INR | age, sex-at-birth, highest caregivers education, caregiver-identified race/ethnicity, handedness score, and MRI device serial number | hemisphere nested within subject nested within site |
| | Lateral aspect of the superior temporal gyrus | INR + D-PRS | age, sex-at-birth, highest caregivers education, caregiver-identified race/ethnicity, handedness score, MRI device serial number, and the first 10 genetics PCs | hemisphere nested within subject nested within site | INR + D-PRS | age, sex-at-birth, highest caregivers education, caregiver-identified race/ethnicity, handedness score, MRI device serial number, and the first 10 genetics PCs | hemisphere nested within subject nested within site | INR | age, sex-at-birth, highest caregivers education, caregiver-identified race/ethnicity, handedness score, and MRI device serial number | hemisphere nested within subject nested within site |
| | Middle temporal gyrus | INR + D-PRS | age, sex-at-birth, highest caregivers education, caregiver-identified race/ethnicity, handedness score, MRI device serial number, and the first 10 genetics PCs | hemisphere nested within subject nested within site | INR + D-PRS | age, sex-at-birth, highest caregivers education, caregiver-identified race/ethnicity, handedness score, MRI device serial number, and the first 10 genetics PCs | hemisphere nested within subject nested within site | INR | age, sex-at-birth, highest caregivers education, caregiver-identified race/ethnicity, handedness score, and MRI device serial number | hemisphere nested within subject nested within site |
| | Superior frontal sulcus | INR + D-PRS | age, sex-at-birth, highest caregivers education, caregiver-identified race/ethnicity, handedness score, MRI device serial number, and the first 10 genetics PCs | hemisphere nested within subject nested within site | INR + D-PRS | age, sex-at-birth, highest caregivers education, caregiver-identified race/ethnicity, handedness score, MRI device serial number, and the first 10 genetics PCs | hemisphere nested within subject nested within site | INR | age, sex-at-birth, highest caregivers education, caregiver-identified race/ethnicity, handedness score, and MRI device serial number | hemisphere nested within subject nested within site |
| | Subparietal sulcus | INR + D-PRS | age, sex-at-birth, highest caregivers education, caregiver-identified race/ethnicity, handedness score, MRI device serial number, and the first 10 genetics PCs | hemisphere nested within subject nested within site | INR + D-PRS | age, sex-at-birth, highest caregivers education, caregiver-identified race/ethnicity, handedness score, MRI device serial number, and the first 10 genetics PCs | hemisphere nested within subject nested within site | INR | age, sex-at-birth, highest caregivers education, caregiver-identified race/ethnicity, handedness score, and MRI device serial number | hemisphere nested within subject nested within site |

| | | | | | | | | | | |
|---|---|---|---|---|---|---|---|---|---|---|
| | Superior temporal sulcus | INR + D-PRS | age, sex-at-birth, highest caregivers education, caregiver-identified race/ethnicity, handedness score, MRI device serial number, and the first 10 genetics PCs | hemisphere nested within subject nested within site | INR + D-PRS | age, sex-at-birth, highest caregivers education, caregiver-identified race/ethnicity, handedness score, MRI device serial number, and the first 10 genetics PCs | hemisphere nested within subject nested within site | INR | age, sex-at-birth, highest caregivers education, caregiver-identified race/ethnicity, handedness score, and MRI device serial number | hemisphere nested within subject nested within site |
| | Anterior part of the cingulate gyrus and sulcus (ACC) | INR + D-PRS | age, sex-at-birth, highest caregivers education, caregiver-identified race/ethnicity, handedness score, MRI device serial number, and the first 10 genetics PCs | hemisphere nested within subject nested within site | INR + D-PRS | age, sex-at-birth, highest caregivers education, caregiver-identified race/ethnicity, handedness score, MRI device serial number, and the first 10 genetics PCs | hemisphere nested within subject nested within site | INR | age, sex-at-birth, highest caregivers education, caregiver-identified race/ethnicity, handedness score, and MRI device serial number | hemisphere nested within subject nested within site |
| | Anterior segment of the circular sulcus of the insula | INR + D-PRS | age, sex-at-birth, highest caregivers education, caregiver-identified race/ethnicity, handedness score, MRI device serial number, and the first 10 genetics PCs | hemisphere nested within subject nested within site | INR + D-PRS | age, sex-at-birth, highest caregivers education, caregiver-identified race/ethnicity, handedness score, MRI device serial number, and the first 10 genetics PCs | hemisphere nested within subject nested within site | INR | age, sex-at-birth, highest caregivers education, caregiver-identified race/ethnicity, handedness score, and MRI device serial number | hemisphere nested within subject nested within site |
| | Transverse frontopolar gyri and sulci | INR + D-PRS | age, sex-at-birth, highest caregivers education, caregiver-identified race/ethnicity, handedness score, MRI device serial number, and the first 10 genetics PCs | hemisphere nested within subject nested within site | INR + D-PRS | age, sex-at-birth, highest caregivers education, caregiver-identified race/ethnicity, handedness score, MRI device serial number, and the first 10 genetics PCs | hemisphere nested within subject nested within site | INR | age, sex-at-birth, highest caregivers education, caregiver-identified race/ethnicity, handedness score, and MRI device serial number | hemisphere nested within subject nested within site |
| | Orbital gyri | INR + D-PRS | age, sex-at-birth, highest caregivers education, caregiver-identified race/ethnicity, handedness score, MRI device serial number, and the first 10 genetics PCs | hemisphere nested within subject nested within site | INR + D-PRS | age, sex-at-birth, highest caregivers education, caregiver-identified race/ethnicity, handedness score, MRI device serial number, and the first 10 genetics PCs | hemisphere nested within subject nested within site | INR | age, sex-at-birth, highest caregivers education, caregiver-identified race/ethnicity, handedness score, and MRI device serial number | hemisphere nested within subject nested within site |
| | Supramarginal gyrus | INR + D-PRS | age, sex-at-birth, highest caregivers education, caregiver-identified race/ethnicity, handedness score, MRI device serial number, and the first 10 genetics PCs | hemisphere nested within subject nested within site | INR + D-PRS | age, sex-at-birth, highest caregivers education, caregiver-identified race/ethnicity, handedness score, MRI device serial number, and the first 10 genetics PCs | hemisphere nested within subject nested within site | INR | age, sex-at-birth, highest caregivers education, caregiver-identified race/ethnicity, handedness score, and MRI device serial number | hemisphere nested within subject nested within site |
| | Inferior frontal sulcus | INR + D-PRS | age, sex-at-birth, highest caregivers education, caregiver-identified race/ethnicity, handedness score, MRI device serial number, and the first 10 genetics PCs | hemisphere nested within subject nested within site | INR + D-PRS | age, sex-at-birth, highest caregivers education, caregiver-identified race/ethnicity, handedness score, MRI device serial number, and the first 10 genetics PCs | hemisphere nested within subject nested within site | INR | age, sex-at-birth, highest caregivers education, caregiver-identified race/ethnicity, handedness score, and MRI device serial number | hemisphere nested within subject nested within site |
| | Middle frontal sulcus | INR + D-PRS | age, sex-at-birth, highest caregivers education, caregiver-identified race/ethnicity, handedness score, MRI device serial number, and the first 10 genetics PCs | hemisphere nested within subject nested within site | INR + D-PRS | age, sex-at-birth, highest caregivers education, caregiver-identified race/ethnicity, handedness score, MRI device serial number, and the first 10 genetics PCs | hemisphere nested within subject nested within site | INR | age, sex-at-birth, highest caregivers education, caregiver-identified race/ethnicity, handedness score, and MRI device serial number | hemisphere nested within subject nested within site |
| | Intraparietal sulcus and transverse parietal sulci | INR + D-PRS | age, sex-at-birth, highest caregivers education, caregiver-identified race/ethnicity, handedness score, MRI device serial number, and the first 10 genetics PCs | hemisphere nested within subject nested within site | INR + D-PRS | age, sex-at-birth, highest caregivers education, caregiver-identified race/ethnicity, handedness score, MRI device serial number, and the first 10 genetics PCs | hemisphere nested within subject nested within site | INR | age, sex-at-birth, highest caregivers education, caregiver-identified race/ethnicity, handedness score, and MRI device serial number | hemisphere nested within subject nested within site |
| | Inferior part of the precentral sulcus | INR + D-PRS | age, sex-at-birth, highest caregivers education, caregiver-identified race/ethnicity, handedness score, MRI device serial number, and the first 10 genetics PCs | hemisphere nested within subject nested within site | INR + D-PRS | age, sex-at-birth, highest caregivers education, caregiver-identified race/ethnicity, handedness score, MRI device serial number, and the first 10 genetics PCs | hemisphere nested within subject nested within site | INR | age, sex-at-birth, highest caregivers education, caregiver-identified race/ethnicity, handedness score, and MRI device serial number | hemisphere nested within subject nested within site |

| | | | | | | | | | | |
|---|---|---|---|---|---|---|---|---|---|---|
| Structural Cortical Surface Area Outcomes | Inferior temporal sulcus | INR + D-PRS | age, sex-at-birth, highest caregivers education, caregiver-identified race/ethnicity, handedness score, MRI device serial number, and the first 10 genetics PCs | hemisphere nested within subject nested within site | INR + D-PRS | age, sex-at-birth, highest caregivers education, caregiver-identified race/ethnicity, handedness score, MRI device serial number, and the first 10 genetics PCs | hemisphere nested within subject nested within site | INR | age, sex-at-birth, highest caregivers education, caregiver-identified race/ethnicity, handedness score, and MRI device serial number | hemisphere nested within subject nested within site |
| | Fronto-marginal gyrus and sulcus | INR + D-PRS | age, sex-at-birth, highest caregivers education, caregiver-identified race/ethnicity, handedness score, MRI device serial number, and the first 10 genetics PCs | hemisphere nested within subject nested within site | INR + D-PRS | age, sex-at-birth, highest caregivers education, caregiver-identified race/ethnicity, handedness score, MRI device serial number, and the first 10 genetics PCs | hemisphere nested within subject nested within site | INR | age, sex-at-birth, highest caregivers education, caregiver-identified race/ethnicity, handedness score, and MRI device serial number | hemisphere nested within subject nested within site |
| | Middle frontal gyrus | INR + D-PRS | age, sex-at-birth, highest caregivers education, caregiver-identified race/ethnicity, handedness score, MRI device serial number, and the first 10 genetics PCs | hemisphere nested within subject nested within site | INR + D-PRS | age, sex-at-birth, highest caregivers education, caregiver-identified race/ethnicity, handedness score, MRI device serial number, and the first 10 genetics PCs | hemisphere nested within subject nested within site | INR | age, sex-at-birth, highest caregivers education, caregiver-identified race/ethnicity, handedness score, and MRI device serial number | hemisphere nested within subject nested within site |
| | Superior frontal gyrus | INR + D-PRS | age, sex-at-birth, highest caregivers education, caregiver-identified race/ethnicity, handedness score, MRI device serial number, and the first 10 genetics PCs | hemisphere nested within subject nested within site | INR + D-PRS | age, sex-at-birth, highest caregivers education, caregiver-identified race/ethnicity, handedness score, MRI device serial number, and the first 10 genetics PCs | hemisphere nested within subject nested within site | INR | age, sex-at-birth, highest caregivers education, caregiver-identified race/ethnicity, handedness score, and MRI device serial number | hemisphere nested within subject nested within site |
| | Middle-posterior part of the cingulate gyrus and sulcus | INR + D-PRS | age, sex-at-birth, highest caregivers education, caregiver-identified race/ethnicity, handedness score, MRI device serial number, and the first 10 genetics PCs | hemisphere nested within subject nested within site | INR + D-PRS | age, sex-at-birth, highest caregivers education, caregiver-identified race/ethnicity, handedness score, MRI device serial number, and the first 10 genetics PCs | hemisphere nested within subject nested within site | INR | age, sex-at-birth, highest caregivers education, caregiver-identified race/ethnicity, handedness score, and MRI device serial number | hemisphere nested within subject nested within site |
| | Angular gyrus | INR + D-PRS | age, sex-at-birth, highest caregivers education, caregiver-identified race/ethnicity, handedness score, MRI device serial number, and the first 10 genetics PCs | hemisphere nested within subject nested within site | INR + D-PRS | age, sex-at-birth, highest caregivers education, caregiver-identified race/ethnicity, handedness score, MRI device serial number, and the first 10 genetics PCs | hemisphere nested within subject nested within site | INR | age, sex-at-birth, highest caregivers education, caregiver-identified race/ethnicity, handedness score, and MRI device serial number | hemisphere nested within subject nested within site |
| | Precuneus | INR + D-PRS | age, sex-at-birth, highest caregivers education, caregiver-identified race/ethnicity, handedness score, MRI device serial number, and the first 10 genetics PCs | hemisphere nested within subject nested within site | INR + D-PRS | age, sex-at-birth, highest caregivers education, caregiver-identified race/ethnicity, handedness score, MRI device serial number, and the first 10 genetics PCs | hemisphere nested within subject nested within site | INR | age, sex-at-birth, highest caregivers education, caregiver-identified race/ethnicity, handedness score, and MRI device serial number | hemisphere nested within subject nested within site |
| | Gyrus rectus | INR + D-PRS | age, sex-at-birth, highest caregivers education, caregiver-identified race/ethnicity, handedness score, MRI device serial number, and the first 10 genetics PCs | hemisphere nested within subject nested within site | INR + D-PRS | age, sex-at-birth, highest caregivers education, caregiver-identified race/ethnicity, handedness score, MRI device serial number, and the first 10 genetics PCs | hemisphere nested within subject nested within site | INR | age, sex-at-birth, highest caregivers education, caregiver-identified race/ethnicity, handedness score, and MRI device serial number | hemisphere nested within subject nested within site |
| | Lateral aspect of the superior temporal gyrus | INR + D-PRS | age, sex-at-birth, highest caregivers education, caregiver-identified race/ethnicity, handedness score, MRI device serial number, and the first 10 genetics PCs | hemisphere nested within subject nested within site | INR + D-PRS | age, sex-at-birth, highest caregivers education, caregiver-identified race/ethnicity, handedness score, MRI device serial number, and the first 10 genetics PCs | hemisphere nested within subject nested within site | INR | age, sex-at-birth, highest caregivers education, caregiver-identified race/ethnicity, handedness score, and MRI device serial number | hemisphere nested within subject nested within site |
| | Middle temporal gyrus | INR + D-PRS | age, sex-at-birth, highest caregivers education, caregiver-identified race/ethnicity, handedness score, MRI device serial number, and the first 10 genetics PCs | hemisphere nested within subject nested within site | INR + D-PRS | age, sex-at-birth, highest caregivers education, caregiver-identified race/ethnicity, handedness score, MRI device serial number, and the first 10 genetics PCs | hemisphere nested within subject nested within site | INR | age, sex-at-birth, highest caregivers education, caregiver-identified race/ethnicity, handedness score, and MRI device serial number | hemisphere nested within subject nested within site |

| | | | | | | | | | | |
|---|---|---|---|---|---|---|---|---|---|---|
| | Superior frontal sulcus | INR + D-PRS | age, sex-at-birth, highest caregivers education, caregiver-identified race/ethnicity, handedness score, MRI device serial number, and the first 10 genetics PCs | hemisphere nested within subject nested within site | INR + D-PRS | age, sex-at-birth, highest caregivers education, caregiver-identified race/ethnicity, handedness score, MRI device serial number, and the first 10 genetics PCs | hemisphere nested within subject nested within site | INR | age, sex-at-birth, highest caregivers education, caregiver-identified race/ethnicity, handedness score, and MRI device serial number | hemisphere nested within subject nested within site |
| | Subparietal sulcus | INR + D-PRS | age, sex-at-birth, highest caregivers education, caregiver-identified race/ethnicity, handedness score, MRI device serial number, and the first 10 genetics PCs | hemisphere nested within subject nested within site | INR + D-PRS | age, sex-at-birth, highest caregivers education, caregiver-identified race/ethnicity, handedness score, MRI device serial number, and the first 10 genetics PCs | hemisphere nested within subject nested within site | INR | age, sex-at-birth, highest caregivers education, caregiver-identified race/ethnicity, handedness score, and MRI device serial number | hemisphere nested within subject nested within site |
| | Superior temporal sulcus | INR + D-PRS | age, sex-at-birth, highest caregivers education, caregiver-identified race/ethnicity, handedness score, MRI device serial number, and the first 10 genetics PCs | hemisphere nested within subject nested within site | INR + D-PRS | age, sex-at-birth, highest caregivers education, caregiver-identified race/ethnicity, handedness score, MRI device serial number, and the first 10 genetics PCs | hemisphere nested within subject nested within site | INR | age, sex-at-birth, highest caregivers education, caregiver-identified race/ethnicity, handedness score, and MRI device serial number | hemisphere nested within subject nested within site |
| | Anterior part of the cingulate gyrus and sulcus (ACC) | INR + D-PRS | age, sex-at-birth, highest caregivers education, caregiver-identified race/ethnicity, handedness score, MRI device serial number, and the first 10 genetics PCs | hemisphere nested within subject nested within site | INR + D-PRS | age, sex-at-birth, highest caregivers education, caregiver-identified race/ethnicity, handedness score, MRI device serial number, and the first 10 genetics PCs | hemisphere nested within subject nested within site | INR | age, sex-at-birth, highest caregivers education, caregiver-identified race/ethnicity, handedness score, and MRI device serial number | hemisphere nested within subject nested within site |
| | Anterior segment of the circular sulcus of the insula | INR + D-PRS | age, sex-at-birth, highest caregivers education, caregiver-identified race/ethnicity, handedness score, MRI device serial number, and the first 10 genetics PCs | hemisphere nested within subject nested within site | INR + D-PRS | age, sex-at-birth, highest caregivers education, caregiver-identified race/ethnicity, handedness score, MRI device serial number, and the first 10 genetics PCs | hemisphere nested within subject nested within site | INR | age, sex-at-birth, highest caregivers education, caregiver-identified race/ethnicity, handedness score, and MRI device serial number | hemisphere nested within subject nested within site |
| **Subcortical Volume** | Amygdala - left hemisphere | INR + D-PRS | age, sex-at-birth, highest caregivers education, caregiver-identified race/ethnicity, handedness score, MRI device serial number, the first 10 genetics PCs, and intracranial volume | subject nested within site | INR + D-PRS | age, sex-at-birth, highest caregivers education, caregiver-identified race/ethnicity, handedness score, MRI device serial number, the first 10 genetics PCs, and intracranial volume | subject nested within site | INR | age, sex-at-birth, highest caregivers education, caregiver-identified race/ethnicity, handedness score, MRI device serial number, and intracranial volume | subject nested within site |
| | Hippocampus - left hemisphere | INR + D-PRS | age, sex-at-birth, highest caregivers education, caregiver-identified race/ethnicity, handedness score, MRI device serial number, the first 10 genetics PCs, and intracranial volume | subject nested within site | INR + D-PRS | age, sex-at-birth, highest caregivers education, caregiver-identified race/ethnicity, handedness score, MRI device serial number, the first 10 genetics PCs, and intracranial volume | subject nested within site | INR | age, sex-at-birth, highest caregivers education, caregiver-identified race/ethnicity, handedness score, MRI device serial number, and intracranial volume | subject nested within site |
| | Amygdala - right hemisphere | INR + D-PRS | age, sex-at-birth, highest caregivers education, caregiver-identified race/ethnicity, handedness score, MRI device serial number, the first 10 genetics PCs, and intracranial volume | subject nested within site | INR + D-PRS | age, sex-at-birth, highest caregivers education, caregiver-identified race/ethnicity, handedness score, MRI device serial number, the first 10 genetics PCs, and intracranial volume | subject nested within site | INR | age, sex-at-birth, highest caregivers education, caregiver-identified race/ethnicity, handedness score, MRI device serial number, and intracranial volume | subject nested within site |
| | Hippocampus - right hemisphere | INR + D-PRS | age, sex-at-birth, highest caregivers education, caregiver-identified race/ethnicity, handedness score, MRI device serial number, the first 10 genetics PCs, and intracranial volume | subject nested within site | INR + D-PRS | age, sex-at-birth, highest caregivers education, caregiver-identified race/ethnicity, handedness score, MRI device serial number, the first 10 genetics PCs, and intracranial volume | subject nested within site | INR | age, sex-at-birth, highest caregivers education, caregiver-identified race/ethnicity, handedness score, MRI device serial number, and intracranial volume | subject nested within site |

This table represents the most parsimonious models analyzed by outcome, including the chosen predictors, covariate, and random effects by outcome in each sample.

**Supplemental Table 13: Withdrawn/depressed symptoms analyses model output**

| | Withdrawn/Depressed Symptom Scale | | | | | | | | |
|---|---|---|---|---|---|---|---|---|---|
| | European-like | | | Not European-like Group 1 | | | Not European-like Group 2 | | |
| | IRR | 95% CI | p-value | IRR | 95% CI | p-value | IRR | 95% CI | p-value |
| Age | 1.124 | [1.103,1.146] | **2.4x10$^{-33}$** | 0.991 | [0.936,1.049] | 0.756 | 0.975 | [0.912,1.042] | 0.449 |
| INR | 0.993 | [0.979,1.008] | 0.372 | 0.926 | [0.897,0.956] | **3.0x10$^{-06}$** | 0.977 | [0.934,1.023] | 0.325 |
| INR-by-Age | | | | 1.02 | [1.005,1.035] | **0.008** | 1.021 | [1.001,1.042] | **0.041** |
| PRS | 1.184 | [1.117,1.254] | **1.1x10$^{-08}$** | 1.223 | [1.022,1.463] | **0.028** | 1.062 | [0.905,1.245] | 0.462 |
| R2 (conditional) | | 0.63126 | | | 0.61877 | | | 0.6207 | |
| R2 (marginal) | | 0.03999 | | | 0.04979 | | | 0.0516 | |
| # of Subjects | | 5308 | | | 1722 | | | 1478 | |
| # of Outcomes | | 13932 | | | 4369 | | | 3584 | |

Model output of covariates of interest for each group examining the effect of income-to-needs (INR) and depression polygenic risk score (D-PRS) on depressed/withdrawn symptom scale. Note, models for the European-like sample did not include INR-by-Age (see Chapter 1 Methods for details). Age centered at 10 years; PRS mean-centered in each group; uncorrected p-values; bolded numbers, p<0.05 (i.e., significant). Abbreviations: IRR = incidence rate ratio; 95% CI = 95 percent confidence interval; # = number.

**Supplemental Table 14: Positive affect analyses model output**

| | Positive Affect | | | | | | | | |
|---|---|---|---|---|---|---|---|---|---|
| | European-like | | | Not European-like Group 1 | | | Not European-like Group 2 | | |
| | Coef. | 95% CI | p-value | Coef. | 95% CI | p-value | Coef. | 95% CI | p-value |
| INR | 0.017 | [-0.011,0.046] | 0.238 | 0.1 | [0.04,0.16] | **0.001** | 0 | [-0.082,0.082] | 0.997 |
| PRS | -0.181 | [-0.298,-0.065] | **0.002** | -0.121 | [-0.485,0.244] | 0.516 | -0.182 | [-0.489,0.125] | 0.245 |
| R2 (conditional) | | 0.41619 | | | 0.36282 | | | 0.34095 | |
| R2 (marginal) | | 0.03453 | | | 0.03161 | | | 0.04866 | |
| # of Subjects | | 5241 | | | 1686 | | | 1429 | |
| # of Outcomes | | 14689 | | | 4551 | | | 3658 | |

Model output of covariates of interest for each group examining the effect of income-to-needs (INR) and depression polygenic risk score (D-PRS) on the positive affect questionnaire. Age centered at 10 years; PRS mean-centered in each group; uncorrected p-values; bolded numbers, p<0.05 (i.e., significant). Abbreviations: Coef. = beta coefficient; 95% CI = 95 percent confidence interval; # = number.

#### Supplemental Table 15: Resting-state functional connectivity analyses model outputs

| | European-Like | | | Not European-Like Group 1 | | | Not European-Like Group 2 | | |
|---|---|---|---|---|---|---|---|---|---|
| | Coef. | 95% CI | p-value | Coef. | 95% CI | p-value | Coef. | 95% CI | p-value |
| **DMN Intra-Network Connectivity** | | | | | | | | | |
| INR | 0 | [-0.001,0] | 0.197 | 0 | [-0.001,0.001] | 0.937 | 0 | [-0.001,0.002] | 0.638 |
| PRS | 0 | [-0.002,0.003] | 0.658 | -0.002 | [-0.009,0.005] | 0.623 | | | |
| R2 (conditional) | | 0.55078 | | | 0.51325 | | | 0.50169 | |
| R2 (marginal) | | 0.15287 | | | 0.19157 | | | 0.11195 | |
| # of Subjects | | 4313 | | | 1378 | | | 1070 | |
| # of Outcomes | | 6570 | | | 2032 | | | 1494 | |
| **FPN Intra-Network Connectivity** | | | | | | | | | |
| INR | -0.001 | [-0.001,0] | 0.038 | 0 | [-0.001,0.002] | 0.504 | 0 | [-0.002,0.001] | 0.836 |
| PRS | 0.002 | [-0.001,0.004] | 0.162 | -0.005 | [-0.012,0.002] | 0.154 | | | |
| R2 (conditional) | | 0.56851 | | | 0.56157 | | | 0.48582 | |
| R2 (marginal) | | 0.07837 | | | 0.1256 | | | 0.09797 | |
| # of Subjects | | 4313 | | | 1378 | | | 1070 | |
| # of Outcomes | | 6570 | | | 2032 | | | 1494 | |
| **SN Intra-Network Connectivity** | | | | | | | | | |
| INR | -0.001 | [-0.002,0.001] | 0.333 | 0.001 | [-0.002,0.003] | 0.618 | -0.001 | [-0.005,0.002] | 0.558 |
| PRS | 0.002 | [-0.002,0.007] | 0.302 | 0.006 | [-0.009,0.021] | 0.423 | | | |
| R2 (conditional) | | 0.48214 | | | 0.37318 | | | 0.44904 | |
| R2 (marginal) | | 0.05058 | | | 0.0606 | | | 0.06577 | |
| # of Subjects | | 4313 | | | 1378 | | | 1070 | |
| # of Outcomes | | 6570 | | | 2032 | | | 1494 | |
| **DMN to FPN Inter-Network Connectivity** | | | | | | | | | |
| INR | 0 | [-0.001,0] | 0.64 | 0.001 | [0,0.002] | 0.206 | 0 | [-0.001,0.002] | 0.53 |
| PRS | 0.002 | [0,0.003] | 0.06 | -0.003 | [-0.009,0.003] | 0.288 | | | |
| R2 (conditional) | | 0.47267 | | | 0.48839 | | | 0.44617 | |
| R2 (marginal) | | 0.04853 | | | 0.06248 | | | 0.09142 | |
| # of Subjects | | 4313 | | | 1378 | | | 1070 | |
| # of Outcomes | | 6570 | | | 2032 | | | 1494 | |
| **DMN to SN Inter-Network Connectivity** | | | | | | | | | |
| INR | 0 | [-0.001,0] | 0.257 | -0.001 | [-0.002,0.001] | 0.388 | 0.001 | [0,0.003] | 0.142 |
| PRS | 0 | [-0.002,0.003] | 0.825 | 0.001 | [-0.007,0.009] | 0.789 | | | |
| R2 (conditional) | | 0.44134 | | | 0.48331 | | | 0.41303 | |
| R2 (marginal) | | 0.02931 | | | 0.04547 | | | 0.04071 | |
| # of Subjects | | 4313 | | | 1378 | | | 1070 | |
| # of Outcomes | | 6570 | | | 2032 | | | 1494 | |
| **FPN to SN Inter-Network Connectivity** | | | | | | | | | |
| INR | 0 | [0,0.001] | 0.648 | 0.001 | [0,0.002] | 0.117 | -0.001 | [-0.003,0.001] | 0.234 |
| PRS | 0.001 | [-0.001,0.004] | 0.348 | -0.01 | [-0.018,-0.003] | 0.009 | | | |
| R2 (conditional) | | 0.51955 | | | 0.50649 | | | 0.48132 | |
| R2 (marginal) | | 0.06909 | | | 0.09229 | | | 0.07635 | |
| # of Subjects | | 4313 | | | 1378 | | | 1070 | |
| # of Outcomes | | 6570 | | | 2032 | | | 1494 | |
| **DMN to Left Amygdala Connectivity** | | | | | | | | | |
| INR | 0 | [0,0.001] | 0.561 | -0.001 | [-0.002,0.001] | 0.373 | -0.001 | [-0.003,0.001] | 0.168 |
| PRS | 0.001 | [-0.002,0.004] | 0.388 | 0 | [-0.01,0.009] | 0.922 | | | |
| R2 (conditional) | | 0.16639 | | | 0.13981 | | | 0.0909 | |
| R2 (marginal) | | 0.02379 | | | 0.03553 | | | 0.06162 | |
| # of Subjects | | 4313 | | | 1378 | | | 1070 | |
| # of Outcomes | | 6570 | | | 2032 | | | 1494 | |
| **DMN to Right Amygdala Connectivity** | | | | | | | | | |
| INR | 0 | [-0.001,0.001] | 0.58 | 0 | [-0.002,0.002] | 0.894 | -0.001 | [-0.004,0.001] | 0.383 |
| PRS | 0.001 | [-0.002,0.004] | 0.597 | 0.005 | [-0.006,0.015] | 0.36 | | | |
| R2 (conditional) | | 0.22769 | | | 0.2989 | | | 0.29038 | |
| R2 (marginal) | | 0.01873 | | | 0.05243 | | | 0.0468 | |
| # of Subjects | | 4310 | | | 1378 | | | 1070 | |
| # of Outcomes | | 6561 | | | 2032 | | | 1494 | |
| **DMN to Left Hippocampus Connectivity** | | | | | | | | | |
| INR | 0 | [0,0.001] | 0.269 | 0.001 | [-0.001,0.002] | 0.285 | 0 | [-0.002,0.001] | 0.697 |
| PRS | 0 | [-0.003,0.002] | 0.711 | 0.006 | [-0.002,0.014] | 0.129 | | | |
| R2 (conditional) | | 0.20597 | | | 0.15645 | | | 0.18587 | |
| R2 (marginal) | | 0.05352 | | | 0.07733 | | | 0.07068 | |
| # of Subjects | | 4312 | | | 1378 | | | 1070 | |
| # of Outcomes | | 6569 | | | 2032 | | | 1494 | |

#### Supplemental Table 15 continued

| | European-Like | | | Not European-Like Group 1 | | | Not European-Like Group 2 | | |
|---|---|---|---|---|---|---|---|---|---|
| | Coef. | 95% CI | p-value | Coef. | 95% CI | p-value | Coef. | 95% CI | p-value |
| **DMN to Right Hippocampus Connectivity** | | | | | | | | | |
| INR | 0.001 | [0,0.001] | 0.253 | 0 | [-0.002,0.002] | 0.715 | 0 | [-0.002,0.003] | 0.744 |
| PRS | -0.003 | [-0.007,0] | 0.07 | -0.006 | [-0.017,0.006] | 0.343 | | | |
| R2 (conditional) | | 0.20369 | | | 0.16606 | | | 0.06077 | |
| R2 (marginal) | | 0.03587 | | | 0.05704 | | | 0.04353 | |
| # of Subjects | | 4313 | | | 1378 | | | 1070 | |
| # of Outcomes | | 6570 | | | 2032 | | | 1494 | |
| **FPN to Left Amygdala Connectivity** | | | | | | | | | |
| INR | 0 | [-0.001,0.001] | 0.996 | 0 | [-0.002,0.001] | 0.735 | 0.002 | [0,0.004] | 0.03 |
| PRS | 0 | [-0.002,0.003] | 0.975 | 0.001 | [-0.007,0.009] | 0.785 | | | |
| R2 (conditional) | | 0.24196 | | | 0.2363 | | | 0.12074 | |
| R2 (marginal) | | 0.061 | | | 0.07113 | | | 0.04229 | |
| # of Subjects | | 4312 | | | 1378 | | | 1070 | |
| # of Outcomes | | 6569 | | | 2032 | | | 1494 | |
| **FPN to Right Amygdala Connectivity** | | | | | | | | | |
| INR | 0 | [-0.001,0.001] | 0.864 | 0 | [-0.002,0.002] | 0.918 | 0.001 | [-0.002,0.003] | 0.509 |
| PRS | 0.003 | [0,0.007] | 0.066 | -0.011 | [-0.022,0] | 0.044 | | | |
| R2 (conditional) | | 0.17629 | | | 0.06733 | | | 0.08208 | |
| R2 (marginal) | | 0.04283 | | | 0.05044 | | | 0.05609 | |
| # of Subjects | | 4313 | | | 1378 | | | 1070 | |
| # of Outcomes | | 6570 | | | 2032 | | | 1494 | |
| **FPN to Left Hippocampus Connectivity** | | | | | | | | | |
| INR | 0 | [-0.001,0.001] | 0.768 | 0.001 | [-0.001,0.003] | 0.516 | 0.001 | [-0.002,0.003] | 0.733 |
| PRS | 0 | [-0.004,0.004] | 0.826 | 0.009 | [-0.004,0.022] | 0.154 | | | |
| R2 (conditional) | | 0.24544 | | | 0.26793 | | | 0.31043 | |
| R2 (marginal) | | 0.07932 | | | 0.08841 | | | 0.07746 | |
| # of Subjects | | 4313 | | | 1378 | | | 1070 | |
| # of Outcomes | | 6570 | | | 2032 | | | 1494 | |
| **FPN to Right Hippocampus Connectivity** | | | | | | | | | |
| INR | 0 | [0,0] | 0.9 | 0.001 | [0,0.001] | 0.106 | 0 | [-0.001,0.001] | 0.811 |
| PRS | -0.001 | [-0.003,0] | 0.075 | -0.003 | [-0.008,0.002] | 0.211 | | | |
| R2 (conditional) | | 0.29713 | | | 0.1996 | | | 0.17285 | |
| R2 (marginal) | | 0.1199 | | | 0.09576 | | | 0.10276 | |
| # of Subjects | | 4313 | | | 1378 | | | 1069 | |
| # of Outcomes | | 6568 | | | 2032 | | | 1490 | |
| **SN to Left Amygdala Connectivity** | | | | | | | | | |
| INR | -0.001 | [-0.002,0.001] | 0.32 | -0.002 | [-0.004,0.001] | 0.194 | 0 | [-0.004,0.003] | 0.883 |
| PRS | -0.001 | [-0.006,0.003] | 0.578 | -0.004 | [-0.019,0.012] | 0.64 | | | |
| R2 (conditional) | | 0.36567 | | | 0.36189 | | | 0.26237 | |
| R2 (marginal) | | 0.03854 | | | 0.04713 | | | 0.03895 | |
| # of Subjects | | 4313 | | | 1378 | | | 1070 | |
| # of Outcomes | | 6570 | | | 2032 | | | 1494 | |
| **SN to Right Amygdala Connectivity** | | | | | | | | | |
| INR | 0.001 | [0,0.001] | 0.088 | 0.001 | [-0.001,0.002] | 0.486 | 0.001 | [-0.001,0.003] | 0.485 |
| PRS | 0.001 | [-0.002,0.003] | 0.663 | 0.005 | [-0.004,0.013] | 0.305 | | | |
| R2 (conditional) | | 0.30205 | | | 0.31414 | | | 0.23354 | |
| R2 (marginal) | | 0.06977 | | | 0.07773 | | | 0.06056 | |
| # of Subjects | | 4312 | | | 1378 | | | 1070 | |
| # of Outcomes | | 6569 | | | 2032 | | | 1494 | |
| **SN to Left Hippocampus Connectivity** | | | | | | | | | |
| INR | 0 | [-0.001,0.001] | 0.694 | 0.001 | [-0.001,0.002] | 0.467 | -0.001 | [-0.004,0.001] | 0.279 |
| PRS | -0.002 | [-0.005,0.001] | 0.122 | -0.006 | [-0.016,0.004] | 0.247 | | | |
| R2 (conditional) | | 0.30948 | | | 0.26183 | | | 0.2187 | |
| R2 (marginal) | | 0.09053 | | | 0.08454 | | | 0.06358 | |
| # of Subjects | | 4311 | | | 1378 | | | 1070 | |
| # of Outcomes | | 6568 | | | 2032 | | | 1494 | |
| **SN to Right Hippocampus Connectivity** | | | | | | | | | |
| INR | 0 | [-0.001,0.001] | 0.619 | -0.001 | [-0.003,0.001] | 0.332 | -0.001 | [-0.004,0.002] | 0.628 |
| PRS | -0.003 | [-0.007,0.002] | 0.22 | 0.002 | [-0.01,0.015] | 0.711 | | | |
| R2 (conditional) | | 0.28718 | | | 0.29405 | | | 0.21838 | |
| R2 (marginal) | | 0.03967 | | | 0.05595 | | | 0.04151 | |
| # of Subjects | | 4313 | | | 1378 | | | 1070 | |
| # of Outcomes | | 6570 | | | 2032 | | | 1494 | |

Model output for covariates of interest in model examining effect of income-to-needs (INR) and depression polygenic risk score (D-PRS) on cortical structural MRI outcomes. Note, models for not European-like Group 2 included INR, but did not include D-PRS (see Chapter 2 Methods). Age centered at 10 years; PRS mean-centered in each group; uncorrected p-values; left and right refers to hemisphere. No findings were trending ($p_{FDR}<0.10$) or were significant ($p_{FDR}<0.05$). Abbreviations: DMN = Default Mode Network; FPN = Frontoparietal Network; SN = Salience Network; Coef. = beta coefficient; 95% CI = 95 percentage confidence interval; # = number.

**Supplemental Table 16: Trending resting-state functional connectivity analyses interaction model outputs**

| | European-Like | | |
|---|---|---|---|
| | Coef. | 95% CI | p-value |
| **DMN Intra-Network Connectivity** | | | |
| INR | 0 | [-0.001,0] | 0.215 |
| PRS | -0.003 | [-0.006,0] | 0.069 |
| INR-by-PRS | 0.001 | [0,0.001] | **0.006** |
| R2 (conditional) | | 0.55075 | |
| R2 (marginal) | | 0.15413 | |
| # of Subjects | | 4313 | |
| # of Outcomes | | 6570 | |
| **FPN Intra-Network Connectivity** | | | |
| INR | -0.001 | [-0.001,0] | 0.043 |
| PRS | -0.002 | [-0.006,0.001] | 0.216 |
| INR-by-PRS | 0.001 | [0,0.001] | **0.005** |
| R2 (conditional) | | 0.56868 | |
| R2 (marginal) | | 0.07979 | |
| # of Subjects | | 4313 | |
| # of Outcomes | | 6570 | |

Model output for covariates of interest in for trending results examining the interactive effect of income-to-needs (INR) and depression polygenic risk score (D-PRS) on resting-state functional MRI outcomes. Age centered at 10 years; PRS mean-centered in each group; uncorrected p-values; bolded numbers, $p_{FDR}<0.1$ (i.e., trend-level). No findings passed FDR correction at $p_{FDR}<0.05$ (i.e., significant). Abbreviations: DMN = Default Mode Network; FPN = Frontoparietal Network; Coef. = beta coefficient; 95% CI = 95 percent confidence interval; # = number.

#### Supplemental Table 17: Cortical analyses model outputs

| | European-Like | | | Not European-Like Group 1 | | | Not European-Like Group 2 | | |
|---|---|---|---|---|---|---|---|---|---|
| | Coef. | 95% CI | p-value | Coef. | 95% CI | p-value | Coef. | 95% CI | p-value |
| **Fronto-marginal gyrus (of Wernicke) and sulcus Thickness** | | | | | | | | | |
| Age | -0.022 | [-0.023,-0.021] | **$6.8x10^{-191}$** | -0.027 | [-0.03,-0.025] | **$6.9x10^{-84}$** | -0.033 | [-0.038,-0.028] | **$2.8x10^{-36}$** |
| INR | 0 | [-0.001,0.001] | 0.771 | 0 | [-0.002,0.002] | 0.986 | 0 | [-0.003,0.003] | 0.979 |
| PRS | -0.003 | [-0.008,0.001] | 0.132 | -0.008 | [-0.022,0.006] | 0.279 | | | |
| INR-by-Age | | | | | | | 0.002 | [0.001,0.004] | **0.006** |
| R2 (conditional) | | 0.74677 | | | 0.74439 | | | 0.72459 | |
| R2 (marginal) | | 0.0504 | | | 0.07321 | | | 0.07587 | |
| # of Subjects | | 9916 | | | 3220 | | | 2718 | |
| # of Outcomes | | 15518 | | | 4882 | | | 3966 | |
| **Fronto-marginal gyrus (of Wernicke) and sulcus Surface Area** | | | | | | | | | |
| Age | 2.305 | [1.612,2.998] | **$7.6x10^{-11}$** | 1.778 | [0.474,3.083] | **0.008** | 0.334 | [-1.282,1.951] | 0.685 |
| INR | 0.026 | [-1.111,1.162] | 0.965 | -0.867 | [-3.192,1.458] | 0.465 | 3.781 | [0.718,6.845] | **0.016** |
| PRS | 2.002 | [-2.624,6.628] | 0.396 | -0.793 | [-14.871,13.286] | 0.912 | | | |
| INR-by-Age | | | | | | | | | |
| R2 (conditional) | | 0.94849 | | | 0.94491 | | | 0.93802 | |
| R2 (marginal) | | 0.06081 | | | 0.06104 | | | 0.07045 | |
| # of Subjects | | 9890 | | | 3210 | | | 2713 | |
| # of Outcomes | | 15408 | | | 4852 | | | 3940 | |
| **Transverse frontopolar gyri and sulci Thickness** | | | | | | | | | |
| Age | -0.027 | [-0.028,-0.025] | **$5.7x10^{-206}$** | -0.03 | [-0.033,-0.027] | **$4.5x10^{-65}$** | -0.027 | [-0.031,-0.023] | **$3.8x10^{-41}$** |
| INR | -0.001 | [-0.002,0.001] | 0.378 | 0.002 | [-0.001,0.004] | 0.243 | 0 | [-0.004,0.004] | 0.97 |
| PRS | 0.002 | [-0.003,0.008] | 0.377 | -0.007 | [-0.024,0.01] | 0.419 | | | |
| INR-by-Age | | | | | | | | | |
| R2 (conditional) | | 0.76499 | | | 0.73044 | | | 0.75245 | |
| R2 (marginal) | | 0.06293 | | | 0.06919 | | | 0.05612 | |
| # of Subjects | | 9916 | | | 3220 | | | 2718 | |
| # of Outcomes | | 15518 | | | 4882 | | | 3966 | |
| **Transverse frontopolar gyri and sulci Surface Area** | | | | | | | | | |
| Age | -0.288 | [-1.072,0.496] | 0.472 | 1.58 | [0.052,3.108] | **0.043** | -3.333 | [-5.085,-1.581] | **$2.0x10^{-04}$** |
| INR | -0.104 | [-1.679,1.47] | 0.897 | -1.012 | [-4.117,2.093] | 0.523 | 3.597 | [-0.374,7.568] | 0.076 |
| PRS | -3.27 | [-9.686,3.147] | 0.318 | -2.473 | [-21.287,16.342] | 0.797 | | | |
| INR-by-Age | | | | | | | | | |
| R2 (conditional) | | 0.96456 | | | 0.9569 | | | 0.95575 | |
| R2 (marginal) | | 0.02894 | | | 0.03505 | | | 0.03944 | |
| # of Subjects | | 9898 | | | 3208 | | | 2715 | |
| # of Outcomes | | 15440 | | | 4849 | | | 3943 | |
| **Anterior part of the cingulate gyrus and sulcus (ACC) Thickness** | | | | | | | | | |
| Age | -0.022 | [-0.023,-0.021] | **$0.0x10^{+00}$** | -0.024 | [-0.026,-0.022] | **$1.5x10^{-126}$** | -0.026 | [-0.029,-0.024] | **$7.1x10^{-83}$** |
| INR | 0.001 | [0,0.001] | 0.152 | 0 | [-0.001,0.002] | 0.616 | -0.002 | [-0.005,0] | 0.09 |
| PRS | 0.002 | [-0.001,0.006] | 0.256 | 0.003 | [-0.007,0.014] | 0.549 | | | |
| INR-by-Age | | | | | | | | | |
| R2 (conditional) | | 0.80534 | | | 0.80002 | | | 0.7741 | |
| R2 (marginal) | | 0.0728 | | | 0.07531 | | | 0.08011 | |
| # of Subjects | | 9916 | | | 3220 | | | 2718 | |
| # of Outcomes | | 15518 | | | 4882 | | | 3966 | |
| **Anterior part of the cingulate gyrus and sulcus (ACC) Surface Area** | | | | | | | | | |
| Age | 10.482 | [9.291,11.674] | **$5.2x10^{-65}$** | 11.389 | [9.216,13.561] | **$4.4x10^{-24}$** | 6.054 | [3.145,8.963] | **$4.8x10^{-05}$** |
| INR | -0.481 | [-3.151,2.189] | 0.724 | 1.469 | [-3.808,6.745] | 0.585 | 1.621 | [-5.345,8.586] | 0.648 |
| PRS | 2.348 | [-8.526,13.223] | 0.672 | -8.299 | [-40.335,23.736] | 0.612 | | | |
| INR-by-Age | | | | | | | | | |
| R2 (conditional) | | 0.9725 | | | 0.97136 | | | 0.96106 | |
| R2 (marginal) | | 0.06764 | | | 0.08396 | | | 0.06831 | |
| # of Subjects | | 9916 | | | 3220 | | | 2718 | |
| # of Outcomes | | 15518 | | | 4882 | | | 3966 | |
| **Middle-posterior part of the cingulate gyrus and sulcus (pMCC) Thickness** | | | | | | | | | |
| Age | -0.018 | [-0.019,-0.018] | **$0.0x10^{+00}$** | -0.019 | [-0.021,-0.017] | **$3.4x10^{-104}$** | -0.02 | [-0.022,-0.018] | **$1.3x10^{-79}$** |
| INR | 0 | [-0.001,0.001] | 0.81 | -0.002 | [-0.003,0] | 0.065 | 0 | [-0.002,0.002] | 0.914 |
| PRS | -0.001 | [-0.004,0.002] | 0.463 | 0.006 | [-0.003,0.016] | 0.197 | | | |
| INR-by-Age | | | | | | | | | |
| R2 (conditional) | | 0.81518 | | | 0.80595 | | | 0.81138 | |
| R2 (marginal) | | 0.06264 | | | 0.06825 | | | 0.07259 | |
| # of Subjects | | 9916 | | | 3220 | | | 2718 | |
| # of Outcomes | | 15518 | | | 4882 | | | 3966 | |
| **Middle-posterior part of the cingulate gyrus and sulcus (pMCC) Surface Area** | | | | | | | | | |
| Age | 2.751 | [2.118,3.384] | **$2.1x10^{-17}$** | 2.333 | [1.155,3.511] | **$1.1x10^{-04}$** | -0.579 | [-2.186,1.028] | 0.48 |
| INR | 0.651 | [-0.458,1.76] | 0.25 | 0.187 | [-2.112,2.487] | 0.873 | 2.753 | [-0.122,5.628] | 0.061 |
| PRS | -2.815 | [-7.333,1.704] | 0.222 | -8.094 | [-22.025,5.837] | 0.255 | | | |
| INR-by-Age | | | | | | | | | |
| R2 (conditional) | | 0.95547 | | | 0.95594 | | | 0.92986 | |
| R2 (marginal) | | 0.07713 | | | 0.09361 | | | 0.07032 | |
| # of Subjects | | 9896 | | | 3213 | | | 2708 | |
| # of Outcomes | | 15450 | | | 4858 | | | 3940 | |
| **Middle frontal gyrus Thickness** | | | | | | | | | |
| Age | -0.015 | [-0.017,-0.014] | **$5.2x10^{-102}$** | -0.018 | [-0.02,-0.016] | **$1.2x10^{-45}$** | -0.024 | [-0.028,-0.02] | **$1.6x10^{-26}$** |
| INR | -0.001 | [-0.002,0] | 0.29 | 0.002 | [0,0.004] | 0.138 | 0 | [-0.003,0.003] | 0.952 |
| PRS | 0.003 | [-0.001,0.007] | 0.15 | 0.005 | [-0.007,0.017] | 0.423 | | | |
| INR-by-Age | | | | | | | 0.002 | [0.001,0.004] | **$2.2x10^{-04}$** |
| R2 (conditional) | | 0.72064 | | | 0.72775 | | | 0.71124 | |
| R2 (marginal) | | 0.09261 | | | 0.07165 | | | 0.0977 | |
| # of Subjects | | 9916 | | | 3220 | | | 2718 | |
| # of Outcomes | | 15518 | | | 4882 | | | 3966 | |

#### Supplemental Table 17 continued

| | European-Like | | | Not European-Like Group 1 | | | Not European-Like Group 2 | | |
|---|---|---|---|---|---|---|---|---|---|
| | Coef. | 95% CI | p-value | Coef. | 95% CI | p-value | Coef. | 95% CI | p-value |
| **Middle frontal gyrus Surface Area** | | | | | | | | | |
| Age | -0.851 | [-3.765,2.063] | 0.567 | -3.602 | [-8.908,1.705] | 0.184 | -12.879 | [-19.278,-6.48] | **$8.4x10^{-05}$** |
| INR | 1.159 | [-2.548,4.865] | 0.54 | 1.94 | [-5.553,9.432] | 0.612 | 12.709 | [2.879,22.54] | **0.011** |
| PRS | -11.224 | [-26.314,3.866] | 0.145 | -31.274 | [-76.735,14.186] | 0.178 | | | |
| INR-by-Age | | | | | | | | | |
| R2 (conditional) | | 0.91738 | | | 0.91762 | | | 0.90899 | |
| R2 (marginal) | | 0.11727 | | | 0.13754 | | | 0.13374 | |
| # of Subjects | | 9916 | | | 3220 | | | 2718 | |
| # of Outcomes | | 15518 | | | 4882 | | | 3966 | |
| **Superior frontal gyrus Thickness** | | | | | | | | | |
| Age | -0.018 | [-0.019,-0.016] | **$2.7x10^{-203}$** | -0.017 | [-0.019,-0.015] | **$1.6x10^{-58}$** | -0.024 | [-0.028,-0.02] | **$5.6x10^{-33}$** |
| INR | 0 | [-0.001,0.001] | 0.777 | 0 | [-0.002,0.002] | 0.742 | -0.001 | [-0.003,0.002] | 0.707 |
| PRS | 0.008 | [0.004,0.012] | **$1.6x10^{-04}$** | 0.006 | [-0.006,0.018] | 0.316 | | | |
| INR-by-Age | | | | | | | 0.002 | [0.001,0.003] | **0.002** |
| R2 (conditional) | | 0.81786 | | | 0.8245 | | | 0.79073 | |
| R2 (marginal) | | 0.08533 | | | 0.06899 | | | 0.08132 | |
| # of Subjects | | 9916 | | | 3220 | | | 2718 | |
| # of Outcomes | | 15518 | | | 4882 | | | 3966 | |
| **Superior frontal gyrus Surface Area** | | | | | | | | | |
| Age | 14.827 | [11.793,17.86] | **$1.4x10^{-21}$** | 11.143 | [5.876,16.411] | **$3.5x10^{-05}$** | -2.518 | [-9.752,4.717] | 0.495 |
| INR | -0.435 | [-5.396,4.525] | 0.863 | 3.08 | [-6.887,13.046] | 0.545 | 7.724 | [-5.099,20.547] | 0.238 |
| PRS | -28.997 | [-49.197,-8.796] | 0.005 | -5.98 | [-66.48,54.52] | 0.846 | | | |
| INR-by-Age | | | | | | | | | |
| R2 (conditional) | | 0.95262 | | | 0.95623 | | | 0.93218 | |
| R2 (marginal) | | 0.15246 | | | 0.15917 | | | 0.12794 | |
| # of Subjects | | 9916 | | | 3220 | | | 2718 | |
| # of Outcomes | | 15518 | | | 4882 | | | 3966 | |
| **Orbital gyri Thickness** | | | | | | | | | |
| Age | -0.023 | [-0.024,-0.021] | **$2.5x10^{-289}$** | -0.025 | [-0.027,-0.022] | **$4.8x10^{-93}$** | -0.028 | [-0.031,-0.025] | **$3.2x10^{-66}$** |
| INR | 0 | [-0.001,0.001] | 0.459 | 0.001 | [-0.001,0.003] | 0.318 | 0.002 | [-0.001,0.004] | 0.267 |
| PRS | 0.005 | [0.001,0.009] | 0.016 | -0.002 | [-0.014,0.009] | 0.68 | | | |
| INR-by-Age | | | | | | | | | |
| R2 (conditional) | | 0.77176 | | | 0.74691 | | | 0.68104 | |
| R2 (marginal) | | 0.05787 | | | 0.08541 | | | 0.07472 | |
| # of Subjects | | 9916 | | | 3220 | | | 2718 | |
| # of Outcomes | | 15518 | | | 4882 | | | 3966 | |
| **Orbital gyri Surface Area** | | | | | | | | | |
| Age | 5.973 | [4.822,7.123] | **$4.0x10^{-24}$** | 6.415 | [4.288,8.541] | **$4.0x10^{-09}$** | -0.388 | [-3.102,2.326] | 0.779 |
| INR | 0.222 | [-1.522,1.966] | 0.803 | 2.273 | [-1.201,5.747] | 0.2 | 3.982 | [-0.527,8.491] | 0.084 |
| PRS | -2.055 | [-9.157,5.046] | 0.571 | -1.626 | [-22.711,19.459] | 0.88 | | | |
| INR-by-Age | | | | | | | | | |
| R2 (conditional) | | 0.94321 | | | 0.93963 | | | 0.92235 | |
| R2 (marginal) | | 0.12972 | | | 0.14338 | | | 0.12846 | |
| # of Subjects | | 9916 | | | 3220 | | | 2718 | |
| # of Outcomes | | 15518 | | | 4882 | | | 3966 | |
| **Angular gyrus Thickness** | | | | | | | | | |
| Age | -0.022 | [-0.024,-0.021] | **$3.5x10^{-184}$** | -0.018 | [-0.021,-0.015] | **$6.5x10^{-32}$** | -0.025 | [-0.028,-0.022] | **$1.4x10^{-48}$** |
| INR | -0.001 | [-0.002,0] | 0.217 | 0.001 | [-0.002,0.003] | 0.507 | 0.002 | [-0.001,0.005] | 0.227 |
| PRS | -0.003 | [-0.007,0.001] | 0.165 | -0.003 | [-0.017,0.011] | 0.654 | | | |
| INR-by-Age | | | | | | | | | |
| R2 (conditional) | | 0.72583 | | | 0.68544 | | | 0.70855 | |
| R2 (marginal) | | 0.1192 | | | 0.12732 | | | 0.11713 | |
| # of Subjects | | 9916 | | | 3220 | | | 2718 | |
| # of Outcomes | | 15518 | | | 4882 | | | 3966 | |
| **Angular gyrus Surface Area** | | | | | | | | | |
| Age | -10.765 | [-12.739,-8.79] | **$2.0x10^{-26}$** | -17.703 | [-21.607,-13.798] | **$1.4x10^{-18}$** | -9.889 | [-14.15,-5.628] | **$5.8x10^{-06}$** |
| INR | 1.397 | [-1.395,4.189] | 0.327 | 6.589 | [0.968,12.21] | 0.022 | -5.123 | [-12.125,1.878] | 0.152 |
| PRS | -9.491 | [-20.861,1.879] | 0.102 | -0.722 | [-34.831,33.387] | 0.967 | | | |
| INR-by-Age | | | | | | | | | |
| R2 (conditional) | | 0.93117 | | | 0.91825 | | | 0.91798 | |
| R2 (marginal) | | 0.08483 | | | 0.10846 | | | 0.10025 | |
| # of Subjects | | 9916 | | | 3220 | | | 2718 | |
| # of Outcomes | | 15518 | | | 4882 | | | 3966 | |
| **Supramarginal gyrus Thickness** | | | | | | | | | |
| Age | -0.014 | [-0.016,-0.012] | **$1.3x10^{-69}$** | -0.011 | [-0.014,-0.008] | **$4.4x10^{-13}$** | -0.019 | [-0.022,-0.016] | **$4.0x10^{-31}$** |
| INR | -0.001 | [-0.002,0] | 0.056 | 0.003 | [0,0.005] | 0.035 | 0.003 | [0,0.006] | 0.071 |
| PRS | 0.001 | [-0.003,0.006] | 0.569 | 0.009 | [-0.006,0.023] | 0.255 | | | |
| INR-by-Age | | | | | | | | | |
| R2 (conditional) | | 0.75353 | | | 0.72074 | | | 0.75165 | |
| R2 (marginal) | | 0.18043 | | | 0.15611 | | | 0.13629 | |
| # of Subjects | | 9916 | | | 3220 | | | 2718 | |
| # of Outcomes | | 15518 | | | 4882 | | | 3966 | |
| **Supramarginal gyrus Surface Area** | | | | | | | | | |
| Age | -9.7 | [-11.582,-7.819] | **$8.1x10^{-24}$** | -12.255 | [-15.699,-8.81] | **$4.3x10^{-12}$** | -7.417 | [-11.477,-3.357] | **$3.5x10^{-04}$** |
| INR | 0.388 | [-2.698,3.474] | 0.806 | 5.393 | [-0.683,11.47] | 0.082 | 6.452 | [-0.986,13.889] | 0.089 |
| PRS | -0.432 | [-12.999,12.135] | 0.946 | -15.045 | [-51.926,21.837] | 0.424 | | | |
| INR-by-Age | | | | | | | | | |
| R2 (conditional) | | 0.95082 | | | 0.94602 | | | 0.93471 | |
| R2 (marginal) | | 0.11486 | | | 0.10229 | | | 0.10002 | |
| # of Subjects | | 9916 | | | 3220 | | | 2718 | |
| # of Outcomes | | 15518 | | | 4882 | | | 3965 | |

#### Supplemental Table 17 continued

| | European-Like | | | Not European-Like Group 1 | | | Not European-Like Group 2 | | |
|---|---|---|---|---|---|---|---|---|---|
| | Coef. | 95% CI | p-value | Coef. | 95% CI | p-value | Coef. | 95% CI | p-value |
| **Precuneus Thickness** | | | | | | | | | |
| Age | -0.026 | [-0.027,-0.026] | **$0.0\text{x}10^{+00}$** | -0.026 | [-0.028,-0.025] | **$1.1\text{x}10^{-168}$** | -0.032 | [-0.034,-0.03] | **$5.2\text{x}10^{-128}$** |
| INR | -0.001 | [-0.002,0] | 0.202 | 0 | [-0.001,0.002] | 0.609 | 0.001 | [-0.002,0.004] | 0.471 |
| PRS | 0.001 | [-0.003,0.005] | 0.705 | 0.011 | [-0.001,0.022] | 0.074 | | | |
| INR-by-Age | | | | | | | | | |
| R2 (conditional) | | 0.87236 | | | 0.85898 | | | 0.82588 | |
| R2 (marginal) | | 0.08059 | | | 0.08628 | | | 0.08064 | |
| # of Subjects | | 9916 | | | 3220 | | | 2718 | |
| # of Outcomes | | 15518 | | | 4882 | | | 3966 | |
| **Precuneus Surface Area** | | | | | | | | | |
| Age | -11.184 | [-12.428,-9.94] | **$9.1\text{x}10^{-68}$** | -12.876 | [-15.308,-10.443] | **$1.6\text{x}10^{-24}$** | -10.067 | [-13.29,-6.845] | **$1.2\text{x}10^{-09}$** |
| INR | -1.821 | [-4.085,0.442] | 0.115 | 1.771 | [-2.819,6.361] | 0.45 | 7.196 | [1.549,12.844] | **0.013** |
| PRS | -3.155 | [-12.372,6.062] | 0.502 | -31.234 | [-59.097,-3.372] | 0.028 | | | |
| INR-by-Age | | | | | | | | | |
| R2 (conditional) | | 0.95895 | | | 0.95325 | | | 0.92824 | |
| R2 (marginal) | | 0.0875 | | | 0.10688 | | | 0.09905 | |
| # of Subjects | | 9916 | | | 3220 | | | 2718 | |
| # of Outcomes | | 15518 | | | 4882 | | | 3966 | |
| **Gyrus rectus Thickness** | | | | | | | | | |
| Age | -0.021 | [-0.023,-0.02] | **$1.4\text{x}10^{-138}$** | -0.02 | [-0.023,-0.017] | **$7.7\text{x}10^{-39}$** | -0.023 | [-0.027,-0.019] | **$1.5\text{x}10^{-31}$** |
| INR | -0.001 | [-0.002,0] | 0.185 | 0 | [-0.002,0.002] | 0.876 | 0 | [-0.003,0.004] | 0.789 |
| PRS | 0.001 | [-0.004,0.005] | 0.79 | 0.001 | [-0.013,0.015] | 0.911 | | | |
| INR-by-Age | | | | | | | | | |
| R2 (conditional) | | 0.65938 | | | 0.65467 | | | 0.61032 | |
| R2 (marginal) | | 0.05382 | | | 0.06914 | | | 0.06227 | |
| # of Subjects | | 9916 | | | 3220 | | | 2718 | |
| # of Outcomes | | 15518 | | | 4882 | | | 3966 | |
| **Gyrus rectus Surface Area** | | | | | | | | | |
| Age | -0.418 | [-1.114,0.278] | 0.239 | -0.888 | [-2.083,0.306] | 0.145 | -2.427 | [-3.939,-0.914] | **0.002** |
| INR | -0.095 | [-0.745,0.555] | 0.774 | 1.173 | [-0.149,2.494] | 0.082 | 0.623 | [-1.114,2.36] | 0.482 |
| PRS | -0.707 | [-3.352,1.938] | 0.6 | -6.519 | [-14.534,1.496] | 0.111 | | | |
| INR-by-Age | | | | | | | | | |
| R2 (conditional) | | 0.84907 | | | 0.86602 | | | 0.82548 | |
| R2 (marginal) | | 0.14778 | | | 0.15716 | | | 0.10754 | |
| # of Subjects | | 9892 | | | 3215 | | | 2712 | |
| # of Outcomes | | 15433 | | | 4861 | | | 3949 | |
| **Lateral aspect of the superior temporal gyrus Thickness** | | | | | | | | | |
| Age | -0.01 | [-0.012,-0.009] | **$6.3\text{x}10^{-36}$** | -0.011 | [-0.014,-0.007] | **$3.9\text{x}10^{-10}$** | -0.013 | [-0.016,-0.009] | **$1.2\text{x}10^{-11}$** |
| INR | -0.001 | [-0.002,0.001] | 0.443 | 0.001 | [-0.003,0.004] | 0.752 | -0.002 | [-0.006,0.002] | 0.3 |
| PRS | 0.001 | [-0.005,0.007] | 0.681 | -0.01 | [-0.029,0.008] | 0.28 | | | |
| INR-by-Age | | | | | | | | | |
| R2 (conditional) | | 0.81878 | | | 0.79893 | | | 0.78988 | |
| R2 (marginal) | | 0.08135 | | | 0.14063 | | | 0.07643 | |
| # of Subjects | | 9916 | | | 3220 | | | 2718 | |
| # of Outcomes | | 15518 | | | 4882 | | | 3966 | |
| **Lateral aspect of the superior temporal gyrus Surface Area** | | | | | | | | | |
| Age | -2.336 | [-3.274,-1.398] | **$1.1\text{x}10^{-06}$** | -3.675 | [-5.601,-1.749] | **$1.9\text{x}10^{-04}$** | -5.163 | [-7.209,-3.116] | **$8.6\text{x}10^{-07}$** |
| INR | 0.811 | [-0.598,2.22] | 0.259 | 0.876 | [-2.034,3.787] | 0.555 | 1.98 | [-1.875,5.835] | 0.314 |
| PRS | -5.621 | [-11.359,0.118] | 0.055 | -4.791 | [-22.454,12.871] | 0.595 | | | |
| INR-by-Age | | | | | | | | | |
| R2 (conditional) | | 0.94397 | | | 0.93016 | | | 0.93973 | |
| R2 (marginal) | | 0.15619 | | | 0.15717 | | | 0.12052 | |
| # of Subjects | | 9915 | | | 3220 | | | 2718 | |
| # of Outcomes | | 15517 | | | 4882 | | | 3966 | |
| **Middle temporal gyrus Thickness** | | | | | | | | | |
| Age | -0.013 | [-0.015,-0.012] | **$1.3\text{x}10^{-61}$** | -0.01 | [-0.013,-0.006] | **$1.2\text{x}10^{-07}$** | -0.014 | [-0.017,-0.01] | **$6.2\text{x}10^{-15}$** |
| INR | 0 | [-0.001,0.002] | 0.44 | 0.001 | [-0.002,0.004] | 0.435 | -0.001 | [-0.004,0.002] | 0.566 |
| PRS | 0.002 | [-0.003,0.007] | 0.494 | -0.006 | [-0.024,0.011] | 0.471 | | | |
| INR-by-Age | | | | | | | | | |
| R2 (conditional) | | 0.7878 | | | 0.73125 | | | 0.76961 | |
| R2 (marginal) | | 0.11389 | | | 0.19661 | | | 0.11673 | |
| # of Subjects | | 9916 | | | 3220 | | | 2718 | |
| # of Outcomes | | 15518 | | | 4882 | | | 3966 | |
| **Middle temporal gyrus Surface Area** | | | | | | | | | |
| Age | 2.058 | [0.687,3.429] | **0.003** | -4.281 | [-7.253,-1.309] | **0.005** | -5.095 | [-8.059,-2.131] | **0.001** |
| INR | 3.381 | [1.084,5.677] | **0.004** | 4.152 | [-0.437,8.741] | 0.076 | 3.166 | [-2.807,9.139] | 0.299 |
| PRS | -3.311 | [-12.664,6.041] | 0.488 | -12.167 | [-40.016,15.682] | 0.392 | | | |
| INR-by-Age | | | | | | | | | |
| R2 (conditional) | | 0.95448 | | | 0.93437 | | | 0.94771 | |
| R2 (marginal) | | 0.14446 | | | 0.17163 | | | 0.12242 | |
| # of Subjects | | 9916 | | | 3220 | | | 2718 | |
| # of Outcomes | | 15518 | | | 4882 | | | 3966 | |
| **Anterior segment of the circular sulcus of the insula Thickness** | | | | | | | | | |
| Age | -0.015 | [-0.016,-0.013] | **$1.1\text{x}10^{-67}$** | -0.02 | [-0.023,-0.017] | **$8.4\text{x}10^{-40}$** | -0.022 | [-0.026,-0.018] | **$4.3\text{x}10^{-31}$** |
| INR | 0.001 | [0,0.002] | 0.132 | 0.001 | [-0.002,0.003] | 0.573 | 0.002 | [-0.002,0.005] | 0.294 |
| PRS | -0.002 | [-0.007,0.003] | 0.342 | -0.003 | [-0.019,0.012] | 0.679 | | | |
| INR-by-Age | | | | | | | | | |
| R2 (conditional) | | 0.72763 | | | 0.73155 | | | 0.70488 | |
| R2 (marginal) | | 0.03553 | | | 0.04798 | | | 0.04938 | |
| # of Subjects | | 9916 | | | 3220 | | | 2718 | |
| # of Outcomes | | 15518 | | | 4882 | | | 3966 | |

#### Supplemental Table 17 continued

| | European-Like | | | Not European-Like Group 1 | | | Not European-Like Group 2 | | |
|---|---|---|---|---|---|---|---|---|---|
| | Coef. | 95% CI | p-value | Coef. | 95% CI | p-value | Coef. | 95% CI | p-value |
| **Anterior segment of the circular sulcus of the insula Surface Area** | | | | | | | | | |
| Age | 2.821 | [2.493,3.15] | **3.0x10$^{-62}$** | 2.852 | [2.246,3.457] | **7.2x10$^{-20}$** | 2.202 | [1.517,2.888] | **4.1x10$^{-10}$** |
| INR | 0.082 | [-0.455,0.62] | 0.763 | 0.191 | [-0.866,1.247] | 0.724 | -0.403 | [-1.733,0.927] | 0.553 |
| PRS | 0.236 | [-1.952,2.423] | 0.833 | 0.992 | [-5.421,7.404] | 0.762 | | | |
| INR-by-Age | | | | | | | | | |
| R2 (conditional) | | 0.94977 | | | 0.9444 | | | 0.9401 | |
| R2 (marginal) | | 0.09833 | | | 0.09354 | | | 0.07052 | |
| # of Subjects | | 9913 | | | 3217 | | | 2716 | |
| # of Outcomes | | 15503 | | | 4874 | | | 3964 | |
| **Inferior frontal sulcus Thickness** | | | | | | | | | |
| Age | -0.016 | [-0.017,-0.015] | **8.1x10$^{-219}$** | -0.018 | [-0.02,-0.016] | **3.0x10$^{-82}$** | -0.019 | [-0.021,-0.016] | **1.9x10$^{-54}$** |
| INR | 0 | [-0.001,0.001] | 0.71 | 0 | [-0.002,0.001] | 0.75 | -0.002 | [-0.004,0.001] | 0.161 |
| PRS | 0.001 | [-0.002,0.004] | 0.499 | -0.004 | [-0.013,0.006] | 0.449 | | | |
| INR-by-Age | | | | | | | | | |
| R2 (conditional) | | 0.76991 | | | 0.7662 | | | 0.73273 | |
| R2 (marginal) | | 0.08261 | | | 0.08427 | | | 0.09853 | |
| # of Subjects | | 9916 | | | 3220 | | | 2718 | |
| # of Outcomes | | 15518 | | | 4882 | | | 3966 | |
| **Inferior frontal sulcus Surface Area** | | | | | | | | | |
| Age | 0.662 | [-0.651,1.975] | 0.323 | -0.55 | [-3.244,2.144] | 0.689 | -7.076 | [-10.684,-3.468] | **1.3x10$^{-04}$** |
| INR | 0.402 | [-1.745,2.549] | 0.714 | 4.924 | [0.42,9.429] | 0.032 | 4.226 | [-1.502,9.953] | 0.148 |
| PRS | -2.246 | [-10.991,6.499] | 0.615 | -9.08 | [-36.418,18.257] | 0.515 | | | |
| INR-by-Age | | | | | | | | | |
| R2 (conditional) | | 0.94941 | | | 0.93966 | | | 0.91267 | |
| R2 (marginal) | | 0.09498 | | | 0.10165 | | | 0.10902 | |
| # of Subjects | | 9916 | | | 3220 | | | 2718 | |
| # of Outcomes | | 15518 | | | 4882 | | | 3966 | |
| **Middle frontal sulcus Thickness** | | | | | | | | | |
| Age | -0.018 | [-0.019,-0.017] | **1.4x10$^{-185}$** | -0.021 | [-0.023,-0.019] | **1.6x10$^{-75}$** | -0.021 | [-0.023,-0.018] | **1.0x10$^{-44}$** |
| INR | 0 | [0,0.001] | 0.375 | 0 | [-0.002,0.002] | 0.784 | 0 | [-0.003,0.002] | 0.801 |
| PRS | 0 | [-0.004,0.003] | 0.826 | 0.004 | [-0.007,0.015] | 0.454 | | | |
| INR-by-Age | | | | | | | | | |
| R2 (conditional) | | 0.7096 | | | 0.71667 | | | 0.65859 | |
| R2 (marginal) | | 0.08827 | | | 0.08363 | | | 0.09719 | |
| # of Subjects | | 9916 | | | 3220 | | | 2718 | |
| # of Outcomes | | 15518 | | | 4882 | | | 3966 | |
| **Middle frontal sulcus Surface Area** | | | | | | | | | |
| Age | 3.755 | [1.949,5.562] | **4.7x10$^{-05}$** | 5.077 | [1.896,8.258] | **0.002** | -3.84 | [-8.128,0.447] | 0.079 |
| INR | -0.746 | [-3.978,2.486] | 0.651 | -4.572 | [-11.051,1.906] | 0.167 | 8.035 | [-0.593,16.663] | 0.068 |
| PRS | -6.195 | [-19.358,6.967] | 0.356 | -19.46 | [-58.79,19.87] | 0.332 | | | |
| INR-by-Age | | | | | | | | | |
| R2 (conditional) | | 0.95549 | | | 0.95779 | | | 0.94369 | |
| R2 (marginal) | | 0.04276 | | | 0.0546 | | | 0.05541 | |
| # of Subjects | | 9907 | | | 3218 | | | 2717 | |
| # of Outcomes | | 15500 | | | 4876 | | | 3961 | |
| **Superior frontal sulcus Thickness** | | | | | | | | | |
| Age | -0.01 | [-0.011,-0.009] | **1.0x10$^{-83}$** | -0.01 | [-0.011,-0.008] | **1.3x10$^{-26}$** | -0.011 | [-0.014,-0.009] | **4.8x10$^{-19}$** |
| INR | 0.001 | [0,0.002] | 0.027 | 0 | [-0.002,0.001] | 0.782 | 0 | [-0.002,0.003] | 0.766 |
| PRS | 0.004 | [0.001,0.007] | 0.018 | 0.005 | [-0.005,0.014] | 0.338 | | | |
| INR-by-Age | | | | | | | | | |
| R2 (conditional) | | 0.75259 | | | 0.76468 | | | 0.70454 | |
| R2 (marginal) | | 0.08114 | | | 0.0587 | | | 0.1145 | |
| # of Subjects | | 9916 | | | 3220 | | | 2718 | |
| # of Outcomes | | 15518 | | | 4882 | | | 3966 | |
| **Superior frontal sulcus Surface Area** | | | | | | | | | |
| Age | 8.323 | [6.214,10.432] | **1.2x10$^{-14}$** | 9.357 | [5.66,13.053] | **7.6x10$^{-07}$** | -2.391 | [-7.407,2.625] | 0.35 |
| INR | 1.331 | [-1.286,3.947] | 0.319 | 2.478 | [-2.756,7.712] | 0.353 | 11.865 | [4.729,19.001] | **0.001** |
| PRS | -9.353 | [-20.005,1.299] | 0.085 | -26.73 | [-58.489,5.028] | 0.099 | | | |
| INR-by-Age | | | | | | | | | |
| R2 (conditional) | | 0.91218 | | | 0.91686 | | | 0.88747 | |
| R2 (marginal) | | 0.10887 | | | 0.12438 | | | 0.0903 | |
| # of Subjects | | 9916 | | | 3220 | | | 2718 | |
| # of Outcomes | | 15518 | | | 4882 | | | 3966 | |
| **Intraparietal sulcus and transverse parietal sulci Thickness** | | | | | | | | | |
| Age | -0.021 | [-0.022,-0.02] | **0.0x10$^{+00}$** | -0.021 | [-0.023,-0.02] | **3.9x10$^{-140}$** | -0.025 | [-0.027,-0.023] | **3.1x10$^{-113}$** |
| INR | 0 | [0,0.001] | 0.266 | 0.001 | [0,0.002] | 0.178 | 0.001 | [-0.001,0.003] | 0.43 |
| PRS | 0 | [-0.002,0.003] | 0.782 | -0.001 | [-0.01,0.008] | 0.83 | | | |
| INR-by-Age | | | | | | | | | |
| R2 (conditional) | | 0.77787 | | | 0.78566 | | | 0.75723 | |
| R2 (marginal) | | 0.09761 | | | 0.09613 | | | 0.09352 | |
| # of Subjects | | 9916 | | | 3220 | | | 2718 | |
| # of Outcomes | | 15518 | | | 4882 | | | 3966 | |
| **Intraparietal sulcus and transverse parietal sulci Surface Area** | | | | | | | | | |
| Age | -9.149 | [-11.196,-7.101] | **2.5x10$^{-18}$** | -14.62 | [-18.278,-10.962] | **8.0x10$^{-15}$** | -9.305 | [-14.215,-4.395] | **2.1x10$^{-04}$** |
| INR | 0.055 | [-2.877,2.987] | 0.971 | 0.023 | [-6.052,6.098] | 0.994 | 3.302 | [-3.901,10.505] | 0.369 |
| PRS | -6.008 | [-17.947,5.931] | 0.324 | -11.217 | [-48.088,25.654] | 0.551 | | | |
| INR-by-Age | | | | | | | | | |
| R2 (conditional) | | 0.93205 | | | 0.93883 | | | 0.89291 | |
| R2 (marginal) | | 0.07319 | | | 0.10196 | | | 0.07561 | |
| # of Subjects | | 9916 | | | 3220 | | | 2718 | |
| # of Outcomes | | 15518 | | | 4882 | | | 3966 | |

#### Supplemental Table 17 continued

| | European-Like | | | Not European-Like Group 1 | | | Not European-Like Group 2 | | |
|---|---|---|---|---|---|---|---|---|---|
| | Coef. | 95% CI | p-value | Coef. | 95% CI | p-value | Coef. | 95% CI | p-value |
| **Inferior part of the precentral sulcus Thickness** | | | | | | | | | |
| Age | -0.007 | [-0.008,-0.006] | **1.8x10$^{-43}$** | -0.007 | [-0.008,-0.005] | **9.7x10$^{-12}$** | -0.008 | [-0.01,-0.005] | **4.3x10$^{-08}$** |
| INR | 0.001 | [0,0.002] | 0.058 | -0.001 | [-0.003,0.001] | 0.272 | -0.001 | [-0.004,0.001] | 0.394 |
| PRS | 0.002 | [-0.002,0.005] | 0.294 | 0.008 | [-0.004,0.019] | 0.185 | | | |
| INR-by-Age | | | | | | | | | |
| R2 (conditional) | | 0.79334 | | | 0.80396 | | | 0.72559 | |
| R2 (marginal) | | 0.02602 | | | 0.03546 | | | 0.02795 | |
| # of Subjects | | 9916 | | | 3220 | | | 2718 | |
| # of Outcomes | | 15518 | | | 4882 | | | 3966 | |
| **Inferior part of the precentral sulcus Surface Area** | | | | | | | | | |
| Age | 1.661 | [0.565,2.757] | **0.003** | 1.225 | [-0.965,3.415] | 0.273 | 0.156 | [-2.354,2.666] | 0.903 |
| INR | 0.658 | [-0.995,2.311] | 0.435 | 2.896 | [-0.416,6.207] | 0.087 | 7.461 | [3.227,11.694] | **0.001** |
| PRS | -0.354 | [-7.08,6.372] | 0.918 | 20.36 | [0.243,40.477] | 0.047 | | | |
| INR-by-Age | | | | | | | | | |
| R2 (conditional) | | 0.93906 | | | 0.92577 | | | 0.92195 | |
| R2 (marginal) | | 0.07566 | | | 0.09976 | | | 0.09146 | |
| # of Subjects | | 9904 | | | 3215 | | | 2713 | |
| # of Outcomes | | 15491 | | | 4870 | | | 3956 | |
| **Subparietal sulcus Thickness** | | | | | | | | | |
| Age | -0.023 | [-0.024,-0.022] | **0.0x10$^{+00}$** | -0.025 | [-0.027,-0.023] | **5.5x10$^{-129}$** | -0.028 | [-0.031,-0.026] | **3.8x10$^{-95}$** |
| INR | 0 | [-0.001,0.001] | 0.891 | 0 | [-0.002,0.002] | 0.93 | 0.001 | [-0.002,0.003] | 0.654 |
| PRS | 0 | [-0.003,0.003] | 0.958 | -0.006 | [-0.016,0.005] | 0.299 | | | |
| INR-by-Age | | | | | | | | | |
| R2 (conditional) | | 0.79485 | | | 0.78021 | | | 0.75128 | |
| R2 (marginal) | | 0.09969 | | | 0.09795 | | | 0.08332 | |
| # of Subjects | | 9916 | | | 3220 | | | 2718 | |
| # of Outcomes | | 15518 | | | 4882 | | | 3966 | |
| **Subparietal sulcus Surface Area** | | | | | | | | | |
| Age | -2.643 | [-3.476,-1.811] | **5.1x10$^{-10}$** | -4.207 | [-5.767,-2.646] | **1.4x10$^{-07}$** | -2.216 | [-4.377,-0.055] | **0.045** |
| INR | -0.36 | [-1.738,1.019] | 0.609 | 0.94 | [-1.857,3.738] | 0.51 | 3.677 | [0.086,7.268] | 0.045 |
| PRS | 0.267 | [-5.348,5.882] | 0.926 | -4.942 | [-21.918,12.034] | 0.568 | | | |
| INR-by-Age | | | | | | | | | |
| R2 (conditional) | | 0.94979 | | | 0.94728 | | | 0.91828 | |
| R2 (marginal) | | 0.0699 | | | 0.08975 | | | 0.07284 | |
| # of Subjects | | 9895 | | | 3208 | | | 2708 | |
| # of Outcomes | | 15438 | | | 4854 | | | 3935 | |
| **Inferior temporal sulcus Thickness** | | | | | | | | | |
| Age | -0.015 | [-0.016,-0.014] | **1.0x10$^{-186}$** | -0.015 | [-0.017,-0.013] | **2.1x10$^{-53}$** | -0.013 | [-0.016,-0.011] | **1.6x10$^{-29}$** |
| INR | 0 | [-0.001,0] | 0.356 | 0 | [-0.002,0.002] | 0.901 | 0 | [-0.003,0.002] | 0.905 |
| PRS | -0.001 | [-0.005,0.002] | 0.469 | 0.004 | [-0.007,0.015] | 0.498 | | | |
| INR-by-Age | | | | | | | | | |
| R2 (conditional) | | 0.82465 | | | 0.81563 | | | 0.81677 | |
| R2 (marginal) | | 0.06364 | | | 0.11717 | | | 0.06162 | |
| # of Subjects | | 9916 | | | 3220 | | | 2718 | |
| # of Outcomes | | 15518 | | | 4882 | | | 3966 | |
| **Inferior temporal sulcus Surface Area** | | | | | | | | | |
| Age | 2.398 | [1.314,3.483] | **1.5x10$^{-05}$** | 3.294 | [1.225,5.364] | **0.002** | -0.007 | [-2.391,2.377] | 0.995 |
| INR | 0.742 | [-0.807,2.291] | 0.348 | 1.019 | [-2.209,4.247] | 0.536 | 1.974 | [-2.32,6.269] | 0.368 |
| PRS | -4.158 | [-10.461,2.146] | 0.196 | -9.908 | [-29.493,9.677] | 0.321 | | | |
| INR-by-Age | | | | | | | | | |
| R2 (conditional) | | 0.93358 | | | 0.93192 | | | 0.93182 | |
| R2 (marginal) | | 0.09695 | | | 0.12137 | | | 0.08999 | |
| # of Subjects | | 9902 | | | 3211 | | | 2713 | |
| # of Outcomes | | 15481 | | | 4867 | | | 3955 | |
| **Superior temporal sulcus Thickness** | | | | | | | | | |
| Age | -0.02 | [-0.02,-0.019] | **0.0x10$^{+00}$** | -0.021 | [-0.022,-0.02] | **5.9x10$^{-184}$** | -0.021 | [-0.023,-0.02] | **5.6x10$^{-117}$** |
| INR | 0.001 | [0,0.001] | 0.1 | 0 | [-0.002,0.001] | 0.531 | 0 | [-0.002,0.002] | 0.648 |
| PRS | 0 | [-0.003,0.003] | 0.966 | 0 | [-0.009,0.009] | 0.989 | | | |
| INR-by-Age | | | | | | | | | |
| R2 (conditional) | | 0.87026 | | | 0.87401 | | | 0.83906 | |
| R2 (marginal) | | 0.08723 | | | 0.12551 | | | 0.08774 | |
| # of Subjects | | 9916 | | | 3220 | | | 2718 | |
| # of Outcomes | | 15518 | | | 4882 | | | 3966 | |
| **Superior temporal sulcus Surface Area** | | | | | | | | | |
| Age | -7.666 | [-9.972,-5.36] | **7.9x10$^{-11}$** | -14.945 | [-19.732,-10.158] | **1.2x10$^{-09}$** | -18.914 | [-23.706,-14.122] | **2.0x10$^{-14}$** |
| INR | 3.145 | [-1.325,7.615] | 0.168 | 3.413 | [-5.488,12.314] | 0.452 | 2.45 | [-8.804,13.705] | 0.67 |
| PRS | -14.723 | [-32.928,3.482] | 0.113 | -5.128 | [-59.157,48.901] | 0.852 | | | |
| INR-by-Age | | | | | | | | | |
| R2 (conditional) | | 0.96687 | | | 0.95576 | | | 0.96298 | |
| R2 (marginal) | | 0.16257 | | | 0.18007 | | | 0.14831 | |
| # of Subjects | | 9916 | | | 3220 | | | 2718 | |
| # of Outcomes | | 15518 | | | 4882 | | | 3966 | |

Model output for covariates of interest in each group examining effect of income-to-needs (INR) and depression polygenic risk score (D-PRS) on structural MRI outcomes. Note, models for not European-like Group 2 included INR and Age, but did not include D-PRS (see Chapter 2 Methods text). Age centered at 10 years; PRS mean-centered in each group; bolded numbers, $p_{FDR}<0.1$ (i.e., trend-level); red numbers passed FDR correction at $p_{FDR}<0.05$ (i.e., significant). Abbreviations: Coef. = beta coefficient; 95% CI = 95 percent confidence interval; # = number.

**Supplemental Table 18: Subcortical analyses model outputs**

| | European-Like | | | Not European-Like Group 1 | | | Not European-Like Group 2 | | |
|---|---|---|---|---|---|---|---|---|---|
| | Coef. | 95% CI | p-value | Coef. | 95% CI | p-value | Coef. | 95% CI | p-value |
| **Amygdala - Left Hemisphere** | | | | | | | | | |
| INR | 0.625 | [-1.201,2.451] | 0.502 | -2.688 | [-6.213,0.838] | 0.135 | 2.459 | [-2.251,7.17] | 0.306 |
| PRS | -5.715 | [-13.139,1.71] | 0.131 | -23.354 | [-44.689,-2.02] | **0.032** | | | |
| R2 (conditional) | | 0.85636 | | | 0.84158 | | | 0.83018 | |
| R2 (marginal) | | 0.39749 | | | 0.42462 | | | 0.33029 | |
| # of Subjects | | 4958 | | | 1610 | | | 1359 | |
| # of Outcomes | | 7759 | | | 2441 | | | 1983 | |
| **Amygdala - Right Hemisphere** | | | | | | | | | |
| INR | 0.554 | [-1.149,2.256] | 0.524 | -1.735 | [-5.036,1.567] | 0.303 | 0.386 | [-3.985,4.757] | 0.863 |
| PRS | -6.472 | [-13.397,0.454] | 0.067 | 2.105 | [-17.891,22.102] | 0.837 | | | |
| R2 (conditional) | | 0.87549 | | | 0.88147 | | | 0.85846 | |
| R2 (marginal) | | 0.38356 | | | 0.41961 | | | 0.37206 | |
| # of Subjects | | 4958 | | | 1610 | | | 1359 | |
| # of Outcomes | | 7759 | | | 2441 | | | 1983 | |
| **Hippocampus - Left Hemisphere** | | | | | | | | | |
| INR | 1.589 | [-1.433,4.612] | 0.303 | -2.152 | [-8.195,3.891] | 0.485 | 1.682 | [-6.725,10.089] | 0.695 |
| PRS | -4.088 | [-16.389,8.213] | 0.515 | -4.143 | [-40.78,32.493] | 0.825 | | | |
| R2 (conditional) | | 0.92904 | | | 0.94143 | | | 0.94363 | |
| R2 (marginal) | | 0.3923 | | | 0.41708 | | | 0.36464 | |
| # of Subjects | | 4958 | | | 1610 | | | 1359 | |
| # of Outcomes | | 7759 | | | 2441 | | | 1983 | |
| **Hippocampus - Right Hemisphere** | | | | | | | | | |
| INR | 1.011 | [-2.243,4.265] | 0.543 | 0.626 | [-5.818,7.07] | 0.849 | 0.625 | [-8.391,9.64] | 0.892 |
| PRS | 1.41 | [-11.838,14.658] | 0.835 | -2.714 | [-41.792,36.364] | 0.892 | | | |
| R2 (conditional) | | 0.9484 | | | 0.95562 | | | 0.94404 | |
| R2 (marginal) | | 0.35636 | | | 0.3865 | | | 0.3324 | |
| # of Subjects | | 4958 | | | 1610 | | | 1359 | |
| # of Outcomes | | 7759 | | | 2441 | | | 1983 | |

Model output for covariates of interest in main effects model examining effect of income-to-needs (INR) and depression polygenic risk score (D-PRS) on subcortical structural MRI outcomes. Note, models for not European-like Group 2 included INR, but did not include D-PRS (see Chapter 2 Methods text). Age centered at 10 years; PRS mean-centered in each group; uncorrected p-values; bolded numbers are $p_{FDR}<0.1$ (i.e., trend-level). No findings passed FDR correction at $p_{FDR}<0.05$ (i.e., significant). Abbreviations: DMN = Default Mode Network; FPN = Frontoparietal Network; SN = Salience Network; Coef. = beta coefficient; 95% CI = 95 percent confidence interval; # = number.

**Supplemental Table 19: Jaccard similarity matrix for behavioral analyses**

#### Withdrawn/Depressed Analyses Sample

***European-Like Sample***

| | Baseline | 1-Year Follow-Up | 2-Year Follow-Up |
|---|---|---|---|
| Baseline | 1 | 0.97 | 0.68 |
| 1-Year Follow-Up | | 1 | 0.69 |
| 2-Year Follow-Up | | | 1 |

***Not European-Like Group 1 Sample***

| | Baseline | 1-Year Follow-Up | 2-Year Follow-Up |
|---|---|---|---|
| Baseline | 1 | 0.94 | 0.62 |
| 1-Year Follow-Up | | 1 | 0.63 |
| 2-Year Follow-Up | | | 1 |

***Not European-Like Group 2 Sample***

| | Baseline | 1-Year Follow-Up | 2-Year Follow-Up |
|---|---|---|---|
| Baseline | 1 | 0.92 | 0.53 |
| 1-Year Follow-Up | | 1 | 0.54 |
| 2-Year Follow-Up | | | 1 |

#### Positive Affect Analyses Sample

***European-Like Sample***

| | 6-Month Follow-Up | 1-Year Follow-Up | 18-Month Follow-Up |
|---|---|---|---|
| Baseline | 1 | 0.97 | 0.94 |
| 1-Year Follow-Up | | 1 | 0.95 |
| 2-Year Follow-Up | | | 1 |

***Not European-Like Group 1 Sample***

| | 6-Month Follow-Up | 1-Year Follow-Up | 18-Month Follow-Up |
|---|---|---|---|
| Baseline | 1 | 0.93 | 0.89 |
| 1-Year Follow-Up | | 1 | 0.91 |
| 2-Year Follow-Up | | | 1 |

***Not European-Like Group 2 Sample***

| | 6-Month Follow-Up | 1-Year Follow-Up | 18-Month Follow-Up |
|---|---|---|---|
| Baseline | 1 | 0.88 | 0.79 |
| 1-Year Follow-Up | | 1 | 0.83 |
| 2-Year Follow-Up | | | 1 |

Jaccard similarity matrix across waves of data for the Withdrawn/Depressed Symptom analyses and the Positive Affect analyses within each sample: European-like, not European-like Group 1, and not European-like Group 2. A higher Jaccard index indicates greater overlap between the subjects for the compared waves of data; a Jaccard index ranges from [0,1].

**Supplemental Table 20: Jaccard similarity matrix for neuroimaging analyses**

**Resting-State Functional MRI Analyses Sample**

*European-Like Sample*

| | Baseline | 2-Year Follow-Up |
|---|---|---|
| Baseline | 1 | 0.62 |
| 2-Year Follow-Up | | 1 |

*Not European-Like Group 1 Sample*

| | Baseline | 2-Year Follow-Up |
|---|---|---|
| Baseline | 1 | 0.56 |
| 2-Year Follow-Up | | 1 |

***Not European-Like Group 2 Sample***

| | Baseline | 2-Year Follow-Up |
|---|---|---|
| Baseline | 1 | 0.45 |
| 2-Year Follow-Up | | 1 |

**Structural MRI Analyses Sample**

***European-Like Sample***

| | Baseline | 2-Year Follow-Up |
|---|---|---|
| Baseline | 1 | 0.66 |
| 2-Year Follow-Up | | 1 |

***Not European-Like Group 1 Sample***

| | Baseline | 2-Year Follow-Up |
|---|---|---|
| Baseline | 1 | 0.6 |
| 2-Year Follow-Up | | 1 |

***Not European-Like Group 2 Sample***

| | Baseline | 2-Year Follow-Up |
|---|---|---|
| Baseline | 1 | 0.52 |
| 2-Year Follow-Up | | 1 |

Jaccard similarity matrix across waves of data for the resting-state functional MRI analyses and the structural MRI analyses within each sample: European-like, not European-like Group 1, and not European-like Group 2. A higher Jaccard index indicates greater overlap between the subjects for the compared waves of data; a Jaccard index ranges from [0,1]. Abbreviations: MRI = magnetic resonance imaging.

#### Supplemental Table 21: Rates of depression in the ABCD Study

| | Baseline Visit (N=5208) | | 2-Year Follow-Up Visit (N=3536) | |
|---|---|---|---|---|
| **KSADS Diagnoses** | *Not Present (%)* | *Present (%)* | *Not Present (%)* | *Present (%)* |
| MDD - Current in Partial Remission | 5202 (99.88%) | 6 (0.12%) | 3522 (99.60%) | 14 (0.40%) |
| MDD - Past | 5089 (97.72%) | 119 (2.28%) | 3425 (96.86%) | 111 (3.14%) |
| MDD - Present | 5201 (99.87%) | 7 (0.13%) | 3525 (99.69%) | 11 (0.31%) |
| MDD - Past & Present | 5082 (97.58%) | 126 (2.42%) | 3410 (96.44%) | 126 (3.56%) |
| Unspecified Depressive Disorder - Current | 5202 (99.88%) | 6 (0.12%) | 3536 (100.00%) | 0 (0.00%) |
| Unspecified Depressive Disorder - Past | 5029 (96.56%) | 179 (3.44%) | 3390 (95.87%) | 146 (4.13%) |
| Unspecified Depressive Disorder - Past & Present | 5024 (96.47%) | 184 (3.53%) | 3390 (95.87%) | 146 (4.13%) |
| MDD & Unspecified Depressive Disorder - Past & Present | 4899 (94.07%) | 309 (5.93%) | 3265 (92.34%) | 271 (7.66%) |

Rates of Major Depressive Disorder (MDD) and/or Unspecified Depressive Disorder utilizing the Kiddie Schedule for Affective Disorders and Schizophrenia for School-Aged Children (K-SADS) caregiver report at baseline visit and 2-year follow-up visit; this data is only presented for the European-like sample used to assess the relationship between depressive polygenic risk scores and income-to-needs rations on depression prodromes. Abbreviations: N = sample size.
